# Genomic, phenomic, and geographic associations of leukocyte telomere length in the United States

**DOI:** 10.1101/2024.11.02.24316529

**Authors:** Tetsushi Nakao, Satoshi Koyama, Buu Truong, Md Mesbah Uddin, Anika Misra, Aniruddh P. Patel, Aarushi Bhatnagar, Victoria Viscosi, Caitlyn Vlasschaert, Alexander G. Bick, Christopher P. Nelson, Veryan Codd, Nilesh J. Samani, Whitney Hornsby, Patrick T. Ellinor, Pradeep Natarajan

## Abstract

Leukocyte telomere length (LTL) is associated with multiple conditions, including cardiovascular diseases and neoplasms, yet their differential associations across diverse individuals are largely unknown. We estimated LTL from blood-derived whole genome sequences in the *All of Us* Research Program (n=242,494) with diverse backgrounds across the United States. LTL was associated with lifestyle, socioeconomic status, biomarkers, cardiometabolic diseases, and neoplasms with heterogeneity across genetic ancestries and sexes. Geographical analysis revealed that significantly longer LTL clustered in the West Coast and Central Midwest, while significantly shorter LTL clustered in the Southeast in the United States, accounting for age, sex, and genetic ancestry. Genome-wide association study and meta-analysis with the UK Biobank (n=679,972) found 234 non-overlapping loci, of which 36 were novel. We identified 4 novel loci unique to non-European-like populations and one specific to females. Rare variant analysis uncovered 7 novel genes, providing new functional insights. Our study highlighted previously underappreciated contextual heterogeneities of phenomic and genomic associations with LTL.

---

Leukocyte telomere length (LTL) has been widely regarded as a marker of biological aging^1,2^. Telomeres, composed of repetitive TTAGGG sequences and associated proteins, span 3 to 15 kb at the ends of chromosomes and shorten during cellular division in cells with imperfect telomerase activity. This gradual shortening leads to reduced telomere length as individuals age. Indeed, studies have demonstrated that shorter LTL is associated with a variety of age-related diseases, such as cardiovascular diseases^3,4^, while longer LTL is associated with healthier lifestyle^5–9^ and less stressful living conditions^10–13^.

Beyond a marker, genetic studies indicate that shorter telomere length might cause increased risk for coronary artery disease (CAD) and interstitial lung disease, and decreased risk for some cancers^9,14^. Cell proliferative conditions, including neoplasms, have varied associations with telomere length. Mendelian short telomere syndromes are enriched for malignant neoplasms^15^, while Mendelian longer telomere is associated with increased risks of various types of neoplasms^16,17^. Given that both longer and shorter telomeres are markers or potential causes of diseases, granular studies in telomere length are warranted to effectively utilize telomere information for propelling population health.

Although telomere length and its attrition rate can vary across different tissues, LTL has served as a representative marker due to relative ease of accessibility and significant correlation of telomere lengths across tissues^18^. Notably, more subtle inter-individual variation in LTL has been linked to increased risks of solid tumors arising from multiple organs^4^. LTL has facilitated large-scale cohort studies of telomere length, yielding important general insights into the relationship between telomere length and diseases/conditions^4,9,19–21^.

There has been known heterogeneity in LTL itself and in associations between LTL and health-related traits, including longer LTL but faster attrition rate in African than European^8,22,23^. However, the lack of a large-scale cohort encompassing multiple genetic ancestries with genomic assay on the same platform has hindered detailed investigations of such heterogeneities. Furthermore, the conventional measurement of telomere length by quantitative polymerase chain reaction (qPCR) using a short range of genome as the control is affected by variations at the control genome, which can be significantly different across genetic ancestries. This is evidenced by the previous genome-wide association study (GWAS) in the UK Biobank (UKB) with qPCR, which found that an African-like population (AFR)-specific variant significantly affected the result^4^.

More recent methods to estimate telomere length from whole genome sequence (WGS) data leverage the entire genome as the control and, thus, are expected to be more robust to differences in particular variants. So far, such aforementioned bias has not yet been reported using these methods^24,25^.

Here, we estimated LTL from 242,494 blood-derived WGS data in the *All of Us* Research Program (AoU) accompanied by rich phenotypes in the diverse U.S. population. We described the demographic and geolocational heterogeneity of LTL and its heterogeneous associations with health-related traits across genetic ancestries and sexes. We further performed GWAS for common variants and rare variant association analysis (RVAS), followed by meta-analysis with UKB. We further described genetic ancestry- and sex-specific genomic predispositions to LTL (Fig. 1).

**Fig. 1:**
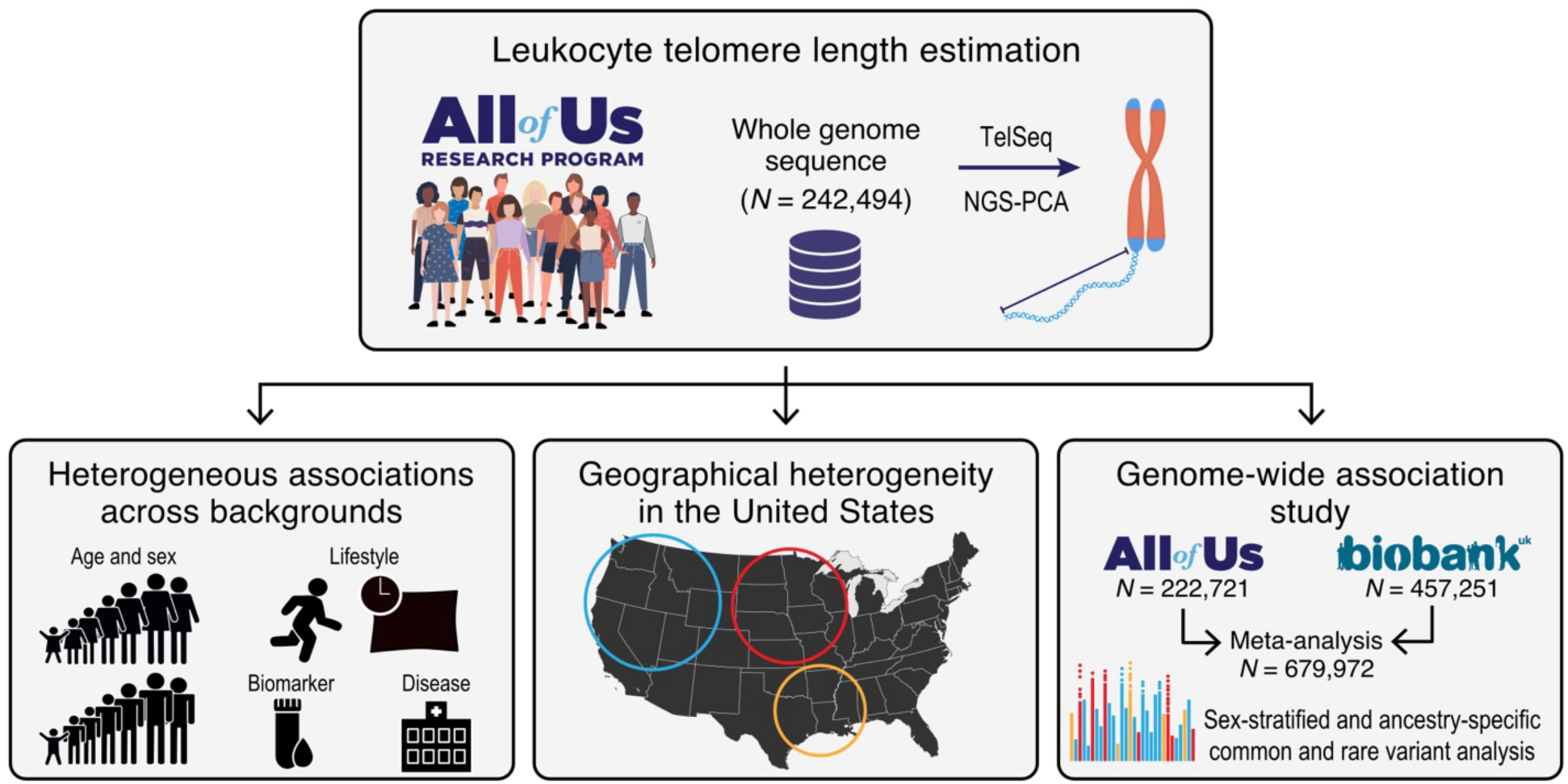
Overview of this study. Leukocyte telomere length (LTL) was estimated from 242,494 blood-derived whole genome sequence data from diverse participants of the National Institute of Health *All of Us* Research Program (AoU) using TelSeq^26^ and NGS-PCA^24^. The associations with demographics, lifestyle, biomarkers, phecodes, and their heterogeneity across genetic ancestries and sexes were investigated. We also examined the geographical heterogeneity of LTL across the United States. We performed genome-wide association studies for common and rare variants in AoU, followed by meta-analyses with UK Biobank, to prioritize novel loci and genes. We further inspected heterogeneities in genetic studies across genetic ancestries and sexes.

## Results

### Estimating leukocyte telomere length (LTL) in the *All of Us* Research Program (AoU)

We estimated LTL from 242,494 high-quality blood-derived WGS in AoU with covariate information, including age, genetically inferred sex (Supplementary Fig. 1), of whom mean age (standard deviation) was 51.65 (16.9), 147,779 (60.9%) were female, and 131,633 (54.3%) were genetically inferred as European-like (EUR). Briefly, we estimated LTL using modified TelSeq^26^, then adjusted for sequencing heterogeneity by regressing it out against principal components (PCs) derived from sequencing depth using mosdepth^27^ and NGS-PCA^24^ (Method, Extended Data Fig. 1).

The PC-regressed-out LTL was strongly correlated with age, sex, and genetic ancestry (Fig. 2a), aligning with previous reports^9,22,23^. Age was more influential in genetically inferred African-like population (AFR, β = -0.0340, *P* = 2.1×10^-846^) than EUR (β = -0.0327, *P* = 3.5×10^1184^, *P*_Ancestry_heterogeneity_ = 9.5×10^8^), aligning with previous reports that African individuals have higher age-related LTL attrition rates than European individuals^22,23^ (Fig. 2b). Heterogeneity across genetic ancestries for the effect of age on LTL was more significant in males (*P*_Ancestry_heterogeneity_ = 1.3×10^12^) than in females (*P*_Ancestry_heterogeneity_ = 0.017).

**Fig. 2:**
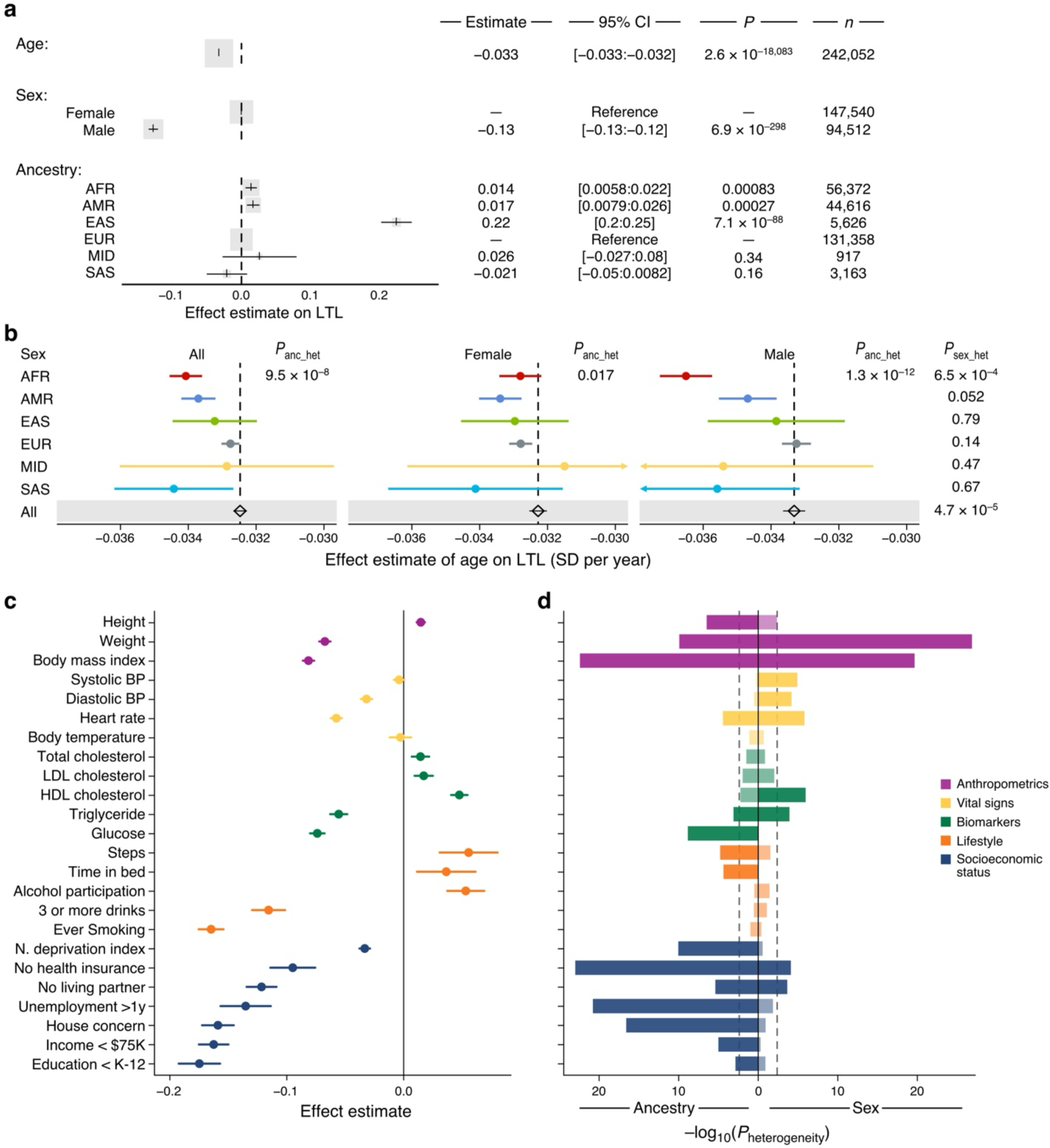
Associations between leukocyte telomere length and demographics/health-related traits in the *All of Us* Research Program. **a**, Linear regression analysis of leukocyte telomere length (LTL) in the *All of Us* Research Program, examining the effects of age, genetically determined sex, and genetic ancestry on LTL, with further adjustment for sequencing site. Effect estimates for each variable are presented with 95% confidence intervals. **b**, Analysis of the heterogeneity in the effect of age on LTL, stratified by genetic ancestry and sex. Effect estimates are presented with 95% confidence intervals. The linear regression model was adjusted for sex, the first 10 genetic principal components (PCs), and sequencing site. Heterogeneity *P*-values for genetic ancestry (*P*_anc_het_) and sex (*P*_sex_het_) were derived by Cochran’s Q test. **c**, The effects of LTL on health-related traits were tested using linear regression model (continuous trait) and logistic regression model (binary trait) adjusted for age, sex, the first 10 genetic PCs, and sequencing site. Effect estimates for each variable are presented with 95% confidence intervals. **d**, The heterogeneities of associations between LTL and traits across genetic ancestries and sexes were tested using Cochran’s Q test. Bars with high transparency indicate non-significance, while opaque bars indicate significance at α < 0.05 with Bonferroni correction (P < 0.0021). AFR: African-like population, AMR: Admixed American-like population, BP: blood pressure, CI: confidence interval, EAS: East Asian-like population, EUR: European-like population, LDL: low-density lipoprotein, HDL: high-density lipoprotein, MID: Middle Eastern-like population, N. deprivation index: neighborhood deprivation index, SAS: South Asian-like population, SD: standard deviation.

### LTL was associated with health-related traits heterogeneously across genetic ancestries and sexes in AoU

We examined whether LTL was observationally associated with health-related traits in AoU, adjusting for age, sex, genetic PCs, and sequencing site. We focused on 5 categories previously reported to be associated with LTL in various cohorts: anthropometrics^4,28^, vital signs^4,29,30^, biomarkers^4,31^, lifestyle^8,9^, and socioeconomic status^9,20^. AoU replicated many of these associations, except the systolic blood pressure (SBP), which was not significant (β = -0.00383, *P* = 0.094), while higher SBP was reported to be associated with shorter LTL in UKB and other cohorts^4,30^ (Fig. 2c, Supplementary Table 1). Of note, alcohol participation ever in life was associated with longer LTL in AoU (β = 0.0532, *P* = 4.0×10^11^); however, the amount of alcohol was associated with shorter LTL (3 or more drinks per day on average: β = -0.116, *P* = 3.5×10^-58^) as reported previously^8,32^.

Next, we tested heterogeneity of these associations across genetic ancestries and sexes. Fifteen out of 24 tested traits showed significant heterogeneity by genetic ancestry after Bonferroni correction (*P* < 0.0021), and 9 phenotypes showed significant heterogeneity by sex (*P* < 0.0021, Fig. 2d). All seven tested socioeconomic status showed heterogeneous associations with LTL by genetic ancestry. Two socioeconomic statuses, “*without living partner”* (β_Female_ = -0.104, β_Male_ = -0.154; *P*_Sex_heterogeneity_ = 2.2×10^4^) and “*stable house concern”* (β_Female_ = -0.134, β_Male_ = -0.189; *P*_Sex_heterogeneity_ = 1.3×10^4^) showed heterogeneous associations with LTL by sex. While SBP was not associated with LTL overall, higher SBP was significantly associated with shorter LTL in females (β = -0.0116, *P* = 5.1×10^5^) but not in males (β = 0.00947, *P* = 0.014, *P*_Sex_heterogeneity_ = 8.4×10^4^). Sex heterogeneities in the LTL-SBP association were most pronounced in EUR (*P*_Sex_heterogeneity_ = 8.7×10^4^) among genetic ancestries.

We computed polygenic scores for LTL (LTL-PGS) in EUR of AoU participants using GWAS summary statistics from UKB (Method). LTL-PGS were significantly associated with anthropometrics, heart rate, smoking, alcohol-related behavior, and socioeconomic status, showing concordant directions with measured LTL (Extended Data Fig. 2, Supplementary Table 1). Residualized LTL by LTL-PGS (LTL-residual) yielded similar results to those of measured LTL with attenuated effect estimates, which was in line with a previous report^21^.

### LTL was associated with diseases and conditions heterogeneously across genetic ancestries and sexes in AoU

We conducted a phenome-wide association study to identify the associations between LTL and a broad spectrum of diseases and conditions. We tested the associations between LTL and phecodes^33,34^ using the logistic regression model adjusting for age, sex, first 10 genetic PCs, and sequencing site in AoU. After Bonferroni correction, increased risk of 54 phecodes out of 1,754 tested was significantly (*P* < 2.9×10^5^) associated with longer LTL, while increased risk of 572 phecodes was associated with shorter LTL (Fig. 3a, Supplementary Table 2).

**Fig. 3:**
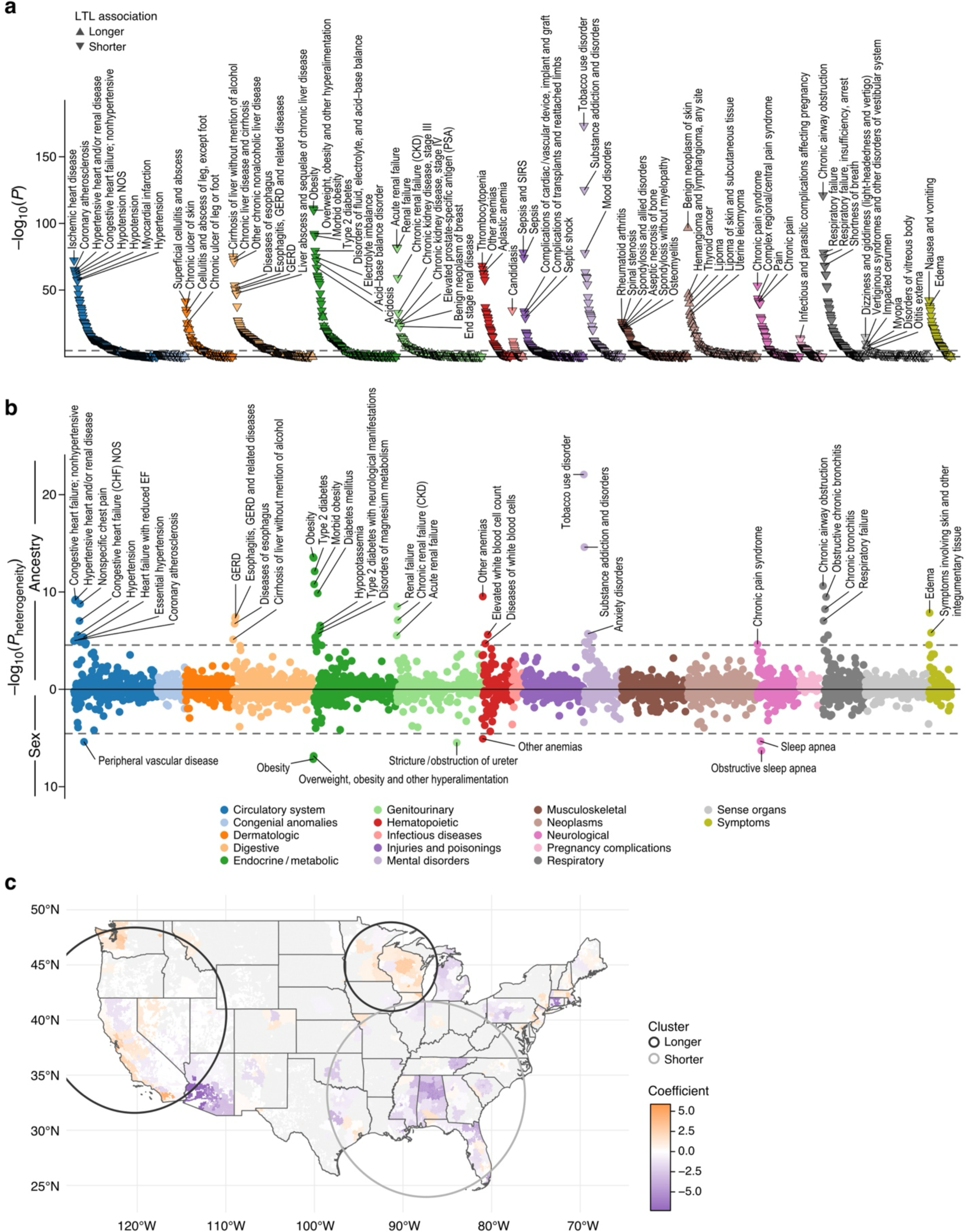
Phenome-wide association study and geographic analysis of leukocyte telomere length in the *All of Us* Research Program. **a**, Associations between phecodes and leukocyte telomere length (LTL) were tested by logistic regression model adjusting for age, sex, sequencing site, and first 10 genetic principal components (PCs). **b**, Heterogeneity of associations between phecode and LTL across genetic ancestries (upward) and sexes (downward) assessed by Cochran’s Q test. **c**, Geographical heterogeneity of LTL among participants across the United States was assessed by linear regression model adjusted with age, sex, first 10 genetic PCs, and sequencing site. Significant clusters were detected by SatScan^84^.

Given the unique relationships between LTL and neoplasms, we inspected the details of cell proliferative conditions, including neoplasms, dysplasia, and cysts (Supplementary Table 2), by assessing the number of significant associations with shorter and longer LTL. Neoplasms have been associated with longer LTL due to the allowance of more cell division (telomere length is the cause of the disease), but tumor cells have short telomeres due to accelerated cell cycles^35,36^ (telomere length is the consequence of the disease). Thus, we distinguished between non-hematologic and hematologic, considering that we measured telomere length in hematologic cells. Among the 49 phecodes for non-hematologic cell proliferative conditions significantly associated with LTL, 85.7% (42/49) of phecodes were associated with longer LTL. In comparison, all the phecodes (11/11) for hematologic cell proliferative conditions with significant LTL-association were associated with shorter LTL. On the other hand, among 566 phecodes for non-cell proliferative conditions significantly associated with LTL, 97.8% (554/566) were associated with shorter LTL.

Next, we tested if the associations were heterogeneous across genetic ancestries and sexes by Cochran’s Q test. Fifty-two phecodes were associated with LTL significantly heterogeneously across genetic ancestries (*P*_Ancestry_heterogeneity_ < 2.9×10^5^), including heart failure, hypertension, gastroesophageal reflux disease, diabetes, renal failure, tobacco use disorders, and respiratory failure. Seven phecodes were associated with LTL significantly heterogeneously between sexes (*P*_Sex_heterogeneity_ < 2.9×10^5^), including peripheral vascular disease and obstructive sleep apnea (Fig. 3b, Supplementary Fig. 2). Obesity was heterogeneous both across genetic ancestries and sexes, in line with the findings in weight and body mass index (BMI, Fig. 2d).

We tested the association of LTL-PGS with phecodes in EUR. We found that 28 phecodes were significantly associated with LTL-PGS (*P* < 2.9×10^5^, Extended Data Fig. 3a, Supplementary Table 2). Twenty-two phecodes significantly associated with longer LTL-PGS were all cell proliferative conditions, including both non-hematologic and hematologic neoplasms. On the contrary, 6 phecodes associated with shorter LTL-PGS were all non-cell proliferative conditions. LTL-residual showed similar association patterns with LTL (Extended Data Fig. 3b, Supplementary Table 2).

We specifically curated (Method) and tested the associations for the following conditions: non-hematologic neoplasms and hematologic neoplasms; clonal hematopoiesis of indeterminate potential (CHIP), a precancerous condition defined by the presence of expanded leukemogenic mutation that we previously showed to be associated with LTL^37^; and CAD. Non-hematologic neoplasms were associated with longer LTL [odds ratio (OR) = 1.07, *P* = 5.2×10^7^] with additional adjustment for total cholesterol, smoking, blood pressure, BMI, and blood glucose (n = 82,371). In comparison, hematologic neoplasms were associated with shorter LTL (OR = 0.8, *P* = 1.8×10^14^). Both non-hematologic and hematologic neoplasms were associated with longer LTL-PGS in EUR (non-hematologic: OR = 1.03, *P* = 0.01; hematologic: OR = 1.08, *P* = 3.8×10^-3^; n = 52,685). CHIP was associated with shorter LTL (β = -0.24, *P* = 4.1×10^14^), as we and others reported before in other datasets^25,37,38^, with heterogeneity across driver genes (Extended Data Fig. 4). Greater variant allele frequency (AF) of CHIP was robustly associated with shorter LTL (β = -0.014, *P* = 1.9×10^11^). Increased risk of CAD was associated with both measured shorter LTL (OR = 0.85, *P* = 6.1×10^-27^) and shorter LTL-PGS (OR = 0.95, *P* = 1.3×10^3^ in EUR) with the same additional adjustment.

### Geographically heterogeneous LTL among U.S. participants

Next, we examined whether LTL varied geographically across the U.S. We calculated the effect estimate of participants’ 3-digit ZIP code for LTL, adjusting with age, sex, the first 10 genetic PCs, and sequencing site. Several large cities and surrounding areas showed higher coefficients than others (Fig. 3c). Cluster analysis showed that the West Coast area and Central Midwest area had significantly higher coefficients (β = 0.040, *P* = 0.002, and β = 0.047, *P* = 0.027, respectively), while the Southeast area clustered significantly lower coefficients (β = -0.073, *P* = 0.001). The same trend held in state-level analysis (Extended Data Fig. 5a). Additional adjustment for BMI, smoking, and neighborhood deprivation index still resulted in similar heterogeneity (Extended Data Fig. 5b-d). EUR-only analysis, LTL-PGS, and LTL-residual in EUR also yielded similar trends (Extended Data Fig. 5e-g).

### Genome-wide association study (GWAS) for common variants in AoU identified 11 novel loci

We performed GWAS for LTL using common variants (minor AF > 0.1%, Extended Data Fig. 6a-e) with genomic control by linkage disequilibrium score (LDSC) regression intercept^39^ (Supplementary Table 3). We observed non-zero heritability for LTL [0.240 (*P* = 1.1×10^6^) in AFR, 0.132 (*P* = 3.3×10^6^) in Admixed-American-like population (AMR), and 0.133 (*P* = 1.0×10^4^) in EUR, Supplementary Table 4]. We found 37 genome-wide significant loci (*P* < 5×10^8^) in AFR, 26 in AMR, 5 in East-Asian-like population (EAS), and 78 in EUR. Among them, 11 were novel (4 in AFR, 1 in AMR, 6 in EUR, Supplementary Table 5). One of the novel loci was found in EUR with the lead variant (chr3:142485886:C:T, AF = 0.177, β = 0.032, *P* = 9.5×10^10^) at an intron of *ATR* in 3q23 (Extended Data Fig. 6f). ATR and ATM (found in previous GWAS locus at 11q22.3 and gene-based test^4,25^) are two primary DNA damage response transducers that are crucial in telomere maintenance^40^. *HBB* locus, a significant locus in the previous UKB GWAS^4^ suspected to be an artifact because of the variants near the qPCR control sequence used for LTL measurements, was not significant in any genetic ancestry in AoU.

### GWAS Meta-analysis with the UK Biobank (UKB) identified 22 additional novel loci

GWAS in UKB was revised using the previously measured LTL by qPCR^4^ (Supplementary Fig. 3a-d) for comparison and meta-analysis with AoU. The genetic correlation between AoU and UKB calculated by LDSC^39^ was 0.958 (*P* = 2.0×10^-332^) in EUR. Effect estimates for AoU (β_AoU_) and UKB (β_UKB_) for lead variants found in AoU were highly correlated (*R*^2^ = 0.91), and effect estimate of β_UKB_ for β_AoU_ in linear regression model was 1.19 (*P* = 1.7×10^83^, Supplementary Fig. 4).

We meta-analyzed AoU GWAS with UKB GWAS by genetic ancestries (Extended Data Fig. 7a-d). We found 38 genome-wide significant loci in AFR, 6 in EAS, 207 in EUR, and 2 in South Asian-like population (SAS), of which 3 in AFR, 1 in EAS, and 20 in EUR were novel (Supplementary Table 5). There was no evidence of inflation, with λ_GC_ being 1.0835 for AFR, 1.0048 for EAS, and 1.0076 for SAS. Although in EUR, we observed moderately elevated λ_GC_ (1.32), the attenuation ratio by LDSC of 0.045 suggested the contribution of the polygenic nature of LTL (Supplementary Table 6). We performed PoPS^41^ to prioritize potentially causal genes with functional plausibility. For example, *GFI1B*, a master transcription factor that is necessary for maintaining hematopoietic stem cell quiescence^42^, was prioritized by PoPS at 9q34.13 locus (Extended Data Fig. 8a). The lead variant chr9:133018743:C:T (nearest gene *GTF3C5*, AF_EUR_ = 0.186, β_EUR_ = -0.015, *P*_EUR_ = 2.5×10^10^) is in linkage disequilibrium (LD) with chr9:133004155:C:T (rs524137, *R*^2^_EUR_ = 0.52), which is a functionally validated causal variant in GWAS for myeloproliferative neoplasms^43^. Chr9:133004155:C:T accelerates the cell cycles of hematopoietic stem cells via GFI1B suppression, aligning to the association with shorter LTL. Another example at novel locus is *RMI2*, a known component of BLM-TOP3A-RMI complex that suppress alternative lengthening of telomere (ALT)^44^, which was prioritized at 16p13.13 locus (lead variant: chr16:11842460:C:T, AF_EUR_ = 0.639, β_EUR_ = -0.011, *P*_EUR_ = 2.0×10^8^, Extended Data Fig. 8b).

Genes identified by agreement on both distance to lead variants and PoPS scores at the locus were reported to have the highest precision and recall^41^. *CKS2*, *PTEN*, and *EXOSC7* were such examples in novel loci. *CKS2* at 9q22.2 (lead variant: chr9:89311173:C:T, AF_EUR_ = 0.0902, β_EUR_ = -0.021, *P*_EUR_ = 1.5×10^10^, Extended Data Fig. 8c) is essential for cell cycle interacting cyclin-dependent kinases^45^. *PTEN* at 10q23.31 (lead variant: chr10:87957355:G:T, AF_EUR_ = 0.0394, β_EUR_ = 0.028, *P*_EUR_ = 2.1×10^9^, Extended Data Fig. 8d) is reported to reduce expression of Telomerase^46,47^. *EXOSC7* at 3p21.31 locus (lead variant: chr3:44976290:A:G, AF_EUR_ = 0.0917, β_EUR_ = -0.019, *P*_EUR_ = 1.4×10^9^, Extended Data Fig. 8e) is a member of the EXOSC family, whose other members has been identified in previous LTL GWAS loci^4,48^. The EXOSC family forms exosome, which contributes to the processing of the template of telomere, human telomerase RNA^49^.

### Genetic ancestry-and sex-specific loci

We identified 3 novel loci in AFR meta-analysis and 1 novel locus in EAS meta-analysis that were not significant in EUR meta-analysis (Supplementary Table 5). For example, chr15:90797341:G:A at 15q26.1 was specific to AFR (AF_AFR_ 0.015, *β*_AFR_ = -0.15, *P*_AFR_ = 6.3×10^11^, AF < 0.001 in other genetic ancestries, Fig. 4a and b, Extended Data Fig. 7e), which was located at the intron of *BLM*. BLM helps resolve or overcome the G-quadruplex structure formed on the G-rich telomere strand when the telomere replicates^50^. Another example, chr10:109948435:A:G was significant only in AFR (AF_AFR_ = 0.776, *β*_AFR_ = 0.045, *P*_AFR_ = 6.2×10^11^, Fig. 4c and d). Though the lead variant is not rare in other genetic ancestries (AF 0.15-0.51), the effect estimates were smaller, which was consistent in a previous multi-ancestry GWAS for LTL in NHLBI Trans-Omics for Precision Medicine (TOPMed^24^, Supplementary Table 7).

**Fig. 4:**
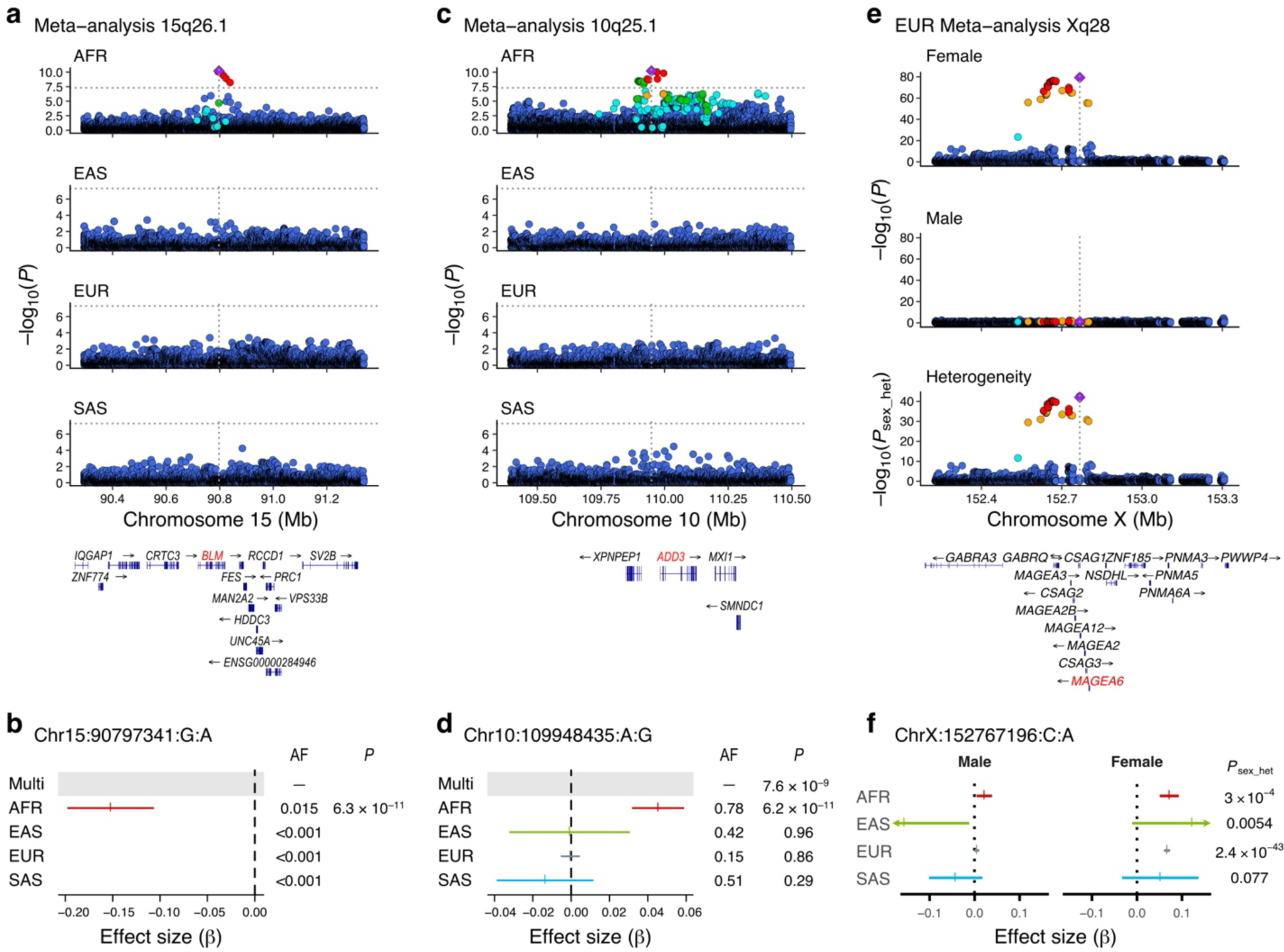
Genetic ancestry- and sex-specific loci in meta-analyses of common-variant genome-wide association studies. Genome-wide association studies in the *All of Us* Research Program and UK Biobank using common variants (minor allele frequency > 0.1%) were meta-analyzed in each genetic ancestry with fixed effect model. Genome-wide significant loci (*P* < 5×10^8^) specific in African-like population (AFR) (**a**-**d**) and specific in females (**e** and **f**) were displayed by locuszoom plot (**a,c,e**) and effect estimates and 95% confidence intervals in each population (**b,d,f**). AF: allele frequency, AFR: African-like population, EAF: effect allele frequency, EAS: East Asian-like population, EUR: European-like population, SAS: South Asian-like population.

We assessed if there was heterogeneity between sexes for genomic predispositions. Sex-stratified GWAS showed high genetic correlations between sexes (0.96, *P* = 7.0×10^404^ in EUR, Supplementary Table 8). Heterogeneity test in EUR showed that one of the previously reported loci at Xq28 was only significant in females, which was consistent in other genetic ancestries (Fig. 4e and f). The lead variant at this locus was chrX:152767196:C:A (AF_Female_ = 0.15, β_Female_ = 0.066, *P*_Female_ = 4.1×10^80^; AF_Male_ = 0.15, β_Male_ = 0.0046, *P*_Male_ = 0.093; *P*_Sex-heterogeneity_ = 2.4×10^43^), a missense variant of *MAGEA6*.

### Multi-ancestry GWAS meta-analysis identified 3 additional novel loci

Finally, multi-ancestry meta-analysis was performed using MR-MEGA with the first 3 PCs of genetic ancestry (Extended Data Fig. 9a and b). There was no evidence of inflation with λ_GC_ being 1.086, yielding more power than previous multi-ancestry GWAS meta-analysis with comparable sample size^51^ (Extended Data Fig. 9c). We found 184 independent loci, of which 13 were novel, and 3 were not found in genetic ancestry specific analyses. *MUS81* prioritized by PoPS at novel 11q13.1 locus (lead variant chr11:65814557:A:G, AF = 0.596, *P* = 6.4×10^9^, Extended Data Fig. 9d) is required for telomere recombination via ALT^52^. *AXIN1* prioritized at novel 16p13.3 locus (lead variant chr16:797597:T:C, AF = 0.297, *P* = 3.6×10^10^, Extended Data Fig. 9e) is a regulator of Wnt signaling^53^, which is a known contributor to telomere maintenance^54,55^.

Overall, our GWASs and meta-analyses identified 199 out of 252 (78.9%) previously reported GWAS loci. The majority of novel loci (28/36, 77.8%) were directionally consistent with previous multi-ancestry GWAS in TOPMed^24^. We found 25 subtelomeric regions (within 2 Mb from telomeres) with significant loci out of 46 subtelomeric regions (54.3%), which may indicate the mechanisms that subtelomeric regions control global telomere length, such as TERRA^56^. The locuszoom plots of all GWAS meta-analysis loci were shown in Supplementary Fig. 5 with available FINEMAP^57^ and PoPS annotations.

### Rare variant association study (RVAS) identified 7 novel genes

To assess the contribution of the rare variations and direct identification of causal genes to LTL, we performed gene-based aggregation tests using deleterious coding variants with alternative AF below 0.1% (Method). RVAS detected 26 genes significantly (*P* < 2.5×10^6^) associated with LTL, of which 7 were not detected by previous RVASs^4,25^ (Fig. 5a, Supplementary Table 9).

**Fig. 5:**
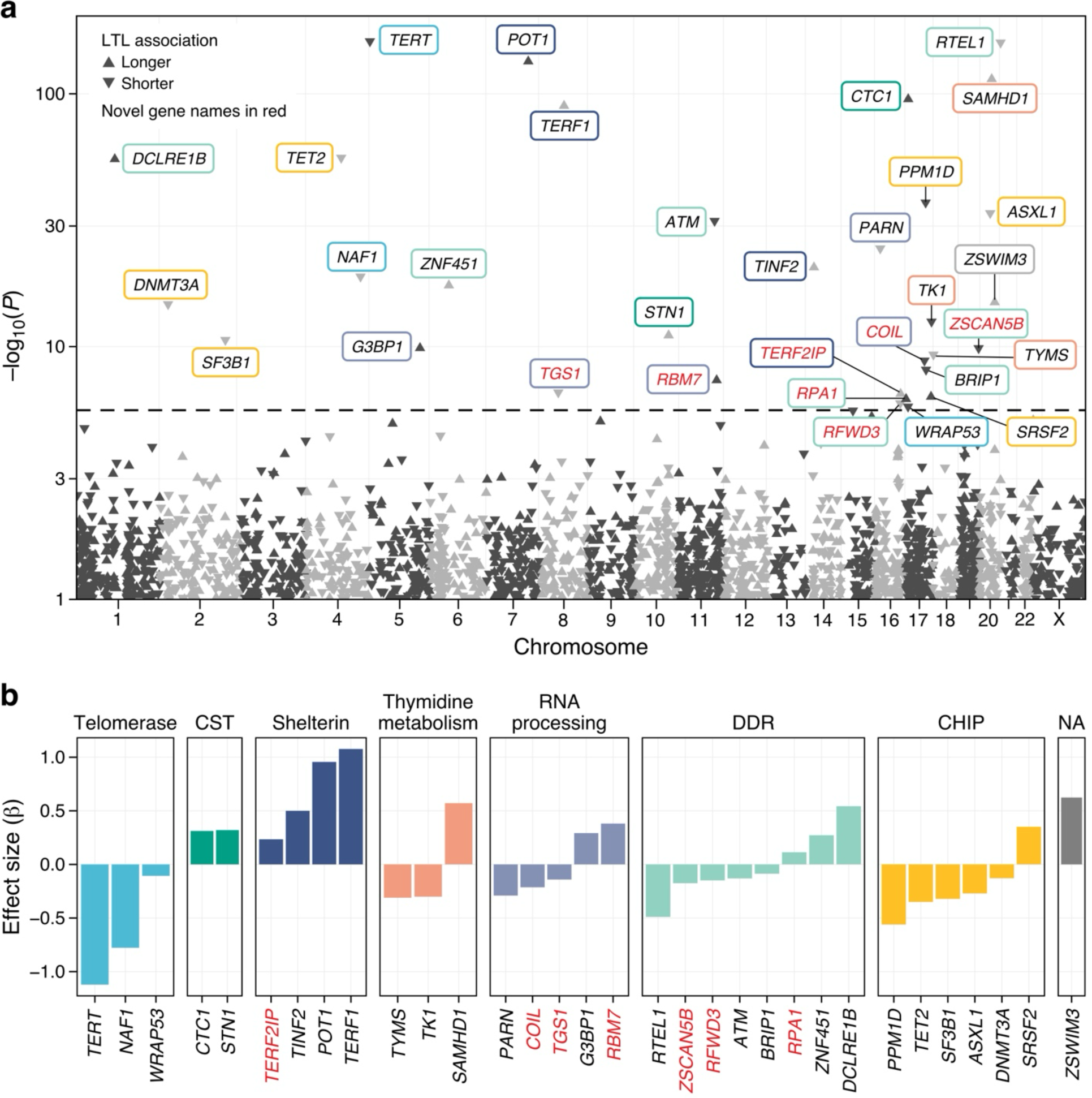
Meta-analysis of rare variant aggregation test for leukocyte telomere length in the *All of Us* Research Program and UK Biobank. We performed multi-ancestry meta-analysis of genome-wide rare variant aggregation tests for leukocyte telomere length (LTL) in the *All of Us* Research Program and UK Biobank. **a,** Upward triangle means association with longer LTL, downward with shorter LTL. Red text indicates novel genes. Box colors indicate functional categories. **b,** genome-wide significant genes (*P* < 2.5×10^6^) were classified by function and plotted with the effect size on the y-axis. Red text indicates novel genes.

Four novel genes, *TGS1*, *RPA1*, *RFWD3*, and *ZSCAN5B*, were located in common variant GWAS loci (Supplementary Fig. 6a-d) but were not described in previous RVAS^4,25^, which all had evidence of involvement in telomere biology. TGS1 forms the 2,2,7-trimethylguanosine cap structure at the human telomerase RNA 5′ end, which recruits telomerase to telomeres and engages Cajal bodies in telomere maintenance^58^. RPA1 binds to telomeres, and gain-of-function mutations in RPA1 cause short telomeres^59^. RFWD3 and ZSCAN5B play roles in DNA damage response^60,61^.

Three novel genes, *TERF2IP*, *RBM7*, and *COIL*, were not located in common variant GWAS loci (Supplementary Fig. 6e-g). *TERF2IP* encodes RAP1, the last component of the Shelterin complex not found in previous GWAS^62^. *RBM7* is a component of the nuclear exosome targeting (NEXT) complex that processes telomerase RNA^49^. *COIL*, which encodes coilin, is debated whether it is involved in telomere maintenance. Despite coilin knockout mice and human cells not showing defects in telomere maintenance, a number of indirect evidence suggest its involvement^63^. Our finding provides orthogonal evidence that coilin is involved in telomere maintenance.

All telomerase component genes found in RVAS were associated with shorter LTL (Fig. 5b). This is consistent with the fact that we primarily included putative loss-of-function or deleterious missense variants in RVAS. On the other hand, all CST component genes were associated with longer LTL, which is consistent with the function of CST to suppress telomerase^64^. All the components of the Shelterin complex, including *POT1*, were consistently associated with longer

LTL. Though the Shelterin complex protects the telomere from instability^65^, loss of function mutations in *POT1* elongates telomeres with unknown mechanisms^16^. Genes related to thymidine metabolism showed the effects in both directions. *TYMS* and *TK1* were associated with shorter LTL, and *SAMHD1* was associated with longer LTL. These effects were directionally consistent with findings in the knockout experiments of these genes^66^.

We found six CHIP-related driver genes associated with LTL. *PPM1D*, *ASXL1*, *DNMT3A*, *SF3B1*, and *TET2* were associated with shorter LTL; however, as CHIP is associated with shorter LTL, we could not exclude that these associations were derived from the residual somatic mutations in the genotypes. *SRSF2*, also a known CHIP driver gene, was associated with longer LTL. CHIP might also influence this association since large clones of *SRSF2* CHIP are associated with longer LTL^25^, and CHIP with small clones was likely excluded from the genotype call.

### Polygenic score (PGS) improved the predictive performance in AFR

We assessed the incremental contribution of increasing genetic ancestry diversity and sample size by meta-analysis on the performance of PGS predictive performance. We re-ran GWAS in AoU, excluding validating (n = 2,000) and testing (n = 8,000) individuals from both EUR and AFR and derived PGS. Compared to PGS derived from UKB alone (partial *R*^2^: 0.00161 in AFR, 0.0712 in EUR), PGS derived from the meta-analysis of UKB and AoU improved the predictive value with prominent improvement in AFR (partial *R*^2^: 0.0425 in AFR, 0.0769 in EUR, Extended Data Fig. 10, Supplementary Table 10). The partial *R*^2^ in AFR was higher in PGS derived from the AFR meta-analysis (n = 51,895, partial R2 = 0.0425) than PGS derived from the EUR meta-analysis encompassing over 10 times more samples (n = 548,812, partial *R*^2^ = 0.0319) suggesting the importance of the diversity in the derivation dataset.

## Discussion

In this study, we estimated LTL in diverse AoU participants via blood-based WGS and evaluated associations with various health-related traits and diseases/conditions. Leveraging diverse genetic ancestries in AoU, we observed that these associations were highly heterogeneous across genetic ancestries and sexes. We further described that the variance of LTL was geographically heterogeneous across the U.S. Lastly, meta-analysis for common variant GWAS and RVAS with UKB revealed 36 novel loci and 7 novel genes, which also uncovered the existence of genetic ancestry- and sex-specific loci.

AoU provides a unique opportunity to evaluate LTL-trait associations across diverse genetic ancestries in a consistent platform, revealing heterogeneities that have been difficult to detect in other datasets. Since qPCR can be affected by the variants at the control sequence^4^, which potentially differs across genetic ancestry, our approach using WGS data is advantageous using whole genome as the reference to compare across genetic ancestries. The heterogeneous associations between LTL and traits indicate some differences among genetic ancestries and sexes may modify the associations between traits and LTL. Further investigation into the mechanisms of heterogeneity may enable novel insights regarding healthy aging across contexts.

The variation in LTL distribution across the U.S. aligns with general population health trends, including life expectancy^67^ and cardiometabolic risk factors, such as BMI^68^, diabetes^69^, hypertension, smoking, and poor diet^70^. Prior research has shown that the health outcomes (e.g., life expectancy, years potential life lost, diabetes, fair or poor health), access to clinical care (e.g., uninsured rates), health behaviors (e.g., smoking, excessive drinking, obesity), and social and economic factors tend to be more favorable in urban areas compared to rural areas in the U.S., despite rural area having more favorable physical environment characteristics (e.g., better air quality and housing availability)^71^. The overlapping geographical heterogeneity in LTL and these traits indicates a potential contextual role of LTL in general health.

We found genetic ancestry- and sex-specific genetic predispositions for LTL, though the majority of the associated loci were shared across these backgrounds. Despite the much smaller sample sizes in those dissimilar to EUR, we found several genetic ancestry-specific variants associated with LTL only in those genetic ancestries, highlighting the critical need for more diverse representation in genetic studies. Ongoing initiatives to expand genomic data across diverse genetic ancestries, such as those spearheaded by AoU and others, along with recent development of methods tailored for multi-ancestry genomic studies, will further genetic discovery and address the disparities in human genetic studies. In addition, the consistent and differential genetic associations across genetic ancestries and sexes implicate important generalizable insights about LTL regulation.

Important limitations should be considered in the interpretation of these findings. First, in the present study, the measured LTL is the average of all the chromosomes and cell types, while LTL has heterogeneity across chromosomes^72^. Long-read sequencing-derived WGS data at the population scale will add further granularity to these analyses. Second, considering LTL is associated with a number of health-related traits, the findings in AoU have a limitation for generalizability outside the U.S.

In conclusion, this study revealed phenomic, genomic, and geographic heterogeneity for LTL, emphasizing that context influences both the distribution and phenotypic associations of LTL. Further understanding of the mechanisms underlying these factors aims to facilitate healthy aging across the diverse communities in the U.S.

## Methods

### Study cohorts

The *All of Us* Research Program (AoU) aims to engage a longitudinal cohort of one million or more U.S. participants, with a focus on including populations that have historically been under-represented in biomedical research. Detailed protocol of the AoU cohort and its genomic data have been described previously^73,74^. Briefly, adults 18 years and older who have the capacity to consent and reside in the U.S. or a U.S. territory at present were eligible. Informed consent for all participants was conducted in person or through an eConsent platform that includes primary consent, HIPAA Authorization for Research use of EHRs and other external health data, and Consent for Return of Genomic Results. The protocol was reviewed by the Institutional Review Board (IRB) of the AoU Research Program. The AoU IRB follows the regulations and guidance of the NIH Office for Human Research Protections for all studies, ensuring that the rights and welfare of research participants are overseen and protected uniformly. The details of the cohort construction were summarized in Supplementary Fig. 1. Briefly, we inspected 245,394 whole genome sequence (WGS) data in AoU version 7 release. We excluded flagged samples for poor sequencing quality by AoU (n = 549), genotyping missingness > 0.01 (n = 395), lacking age information (n = 26), and genetically undetermined sex (n = 1,930) to construct a cohort of 242,494 participants included in our analysis. For epidemiologic studies, we further excluded outliers (> 20 median absolute deviations) for principal components (PCs) calculated from sequencing depth (NGS-PCs, n = 442). For genomic studies, we further excluded < 0.75 for random forest probabilities for the largest 5 population groups [African-like population (AFR), Admixed American-like population (AMR), East Asian-like population (EAS), European-like population (EUR), and South Asian-like population (SAS)] in AoU (n = 19,511), missing genotyping array information (n < 21, According to AoU publishing policy, numbers below 21 cannot be published. Precise counts are not calculable due to overlapping samples across multiple QC metrics.), and outliers (> 20 median absolute deviation) for NGS-PCs (n = 261).

The UK Biobank (UKB) is a population-based cohort of >500,000 UK adult residents recruited between 2006 and 2010 and followed prospectively via linkage to national health records^75^. Secondary analyses of the UKB data under Application 7089 were approved by the Massachusetts General Hospital IRB. At the baseline study visit, consented participants underwent phlebotomy and provided detailed information about medical history and medication use. In the present study, the UKB cohort included adults aged 40 to 70 years at blood draw with available genotyping array or whole exome sequences (WES)^76^. We used genotyping array for genome-wide association study for common variants (N = 488,188) and WES for rare variant association study (N = 452,929). We excluded samples with consent retraction, excess heterogeneity, sex discordance, or missingness/heterogeneity/outliers in the genotype, as summarized in Supplementary Fig. 1.

### Leukocyte telomere length (LTL) Estimation

TelSeq^26^ was used in a previous genome-wide association study for WGS-derived LTL^24,25^, while more recent algorithms for WGS-derived LTL estimation have been published, including Telomerecat^77^. We compared the two methods using 1,000 random participants in UKB with available quantitative polymerase chain reaction (qPCR) measurements of LTL and WGS for the same individuals. TelSeq had better agreement with qPCR with similar *R*^2^ to previous report^25^ (Extended Data Fig. 1a). Thus, we estimated the LTL using TelSeq^26^ with available WGS CRAM files in AoU. To align with the computational demand on the AoU workbench system, we modified the TelSeq integrating htslib-1.18^78^ to enable direct CRAM file processing, parallel processing across the cores, and streaming processing from Google Cloud Storage, which achieved 4 times better efficiency (Extended Data Fig. 1c and d). The estimated LTL was verified as identical to the original software in the randomly selected 1,000 European WGS CRAM files in UKB and AoU (Extended Data Fig. 1b). The parameter k for TelSeq was set to 10, in line with the previous reports^24,25^.

We adjusted the estimated telomere length with sequencing depth as described before^24^. Briefly, we performed mosdepth v0.3.5^27^ on each CRAM file to calculate the sequencing depth in 1,000-base pair bins across the genome. Using the resultant depth information, we calculated 200 PCs using NGS-PCA (https://github.com/PankratzLab/NGS-PCA). NGS-PCA was modified to enable stream processing from Google Cloud Storage to deal with large samples on the AoU Workbench system. Due to the limitation of the computing resource available in the AoU workbench, the PCs were separately calculated within each genetic ancestry for GWAS studies and within each sequencing site for epidemiological studies. The first 50 PCs were verified identical to the original software output using 100 random samples in AoU. We used PCs explaining more than 0.1% of the total variance calculated to residualize the LTL (Supplementary Fig. 7). We excluded samples with extreme outliers (over 20 median absolute deviations in each group) with used NGS-derived PCs. The number of PCs used for each group and the number of excluded samples are shown in Supplementary Table 11. LTL was residualized by NGS-PCs to obtain relative LTL adjusted for sequencing heterogeneity. This adjustment increased the power to detect previously known associations with genomic variation and suppressed spurious associations (Extended Data Fig. 1e and f).

### Epidemiology

Phenotypes (outcome) were tested for associations with LTL (exposure) by linear regression model (continuous traits) or logistic regression model (binary traits) with adjustment for age, sex, first ten genetic PCs, and sequencing site. Analyses with cases less than 21 were excluded. Phenotype definitions were listed in Supplementary Table 12, unless described below.

#### Wearable device data

Details of wearable device data in AoU were described previously^79^. Briefly, Participants who provided primary consent to be part of the AoU and share EHR data had an opportunity to provide their data from wearable devices under the Bring Your Own Device program. Participants connected their own Fitbit device account with the AoU Participant Portal and agreed to share their complete data on their device over all time, including previous data, not just recent data. A participant could stop sharing their data at any time. Participants’ data had direct identifiers removed, and all datetime fields were subjected to date shifting by a random number between 1 and 365 days in accordance with approved AoU privacy policies. We analyzed the median of steps per day and the median sleep duration per day.

#### Neighborhood deprivation index

Neighborhood deprivation indices^80^ for each 3-digit prefix of ZIP codes were calculated by AoU and accessible in the controlled tier. Briefly, a neighborhood deprivation index for each census tract in the United States was calculated based on a principal component analysis of six different 2015 American Community Survey measures (fraction of households receiving public assistance income or food stamps or Supplemental Nutrition Assistance Program (SNAP) participation in the past 12 months, the fraction of population 25 and older with the educational attainment of at least high school graduation includes GED equivalency, median household income in the past 12 months in 2015 inflation-adjusted dollars, the fraction of population with no health insurance coverage, the fraction of population with income in past 12 months below poverty level, and fraction of houses that are vacant); rescaling and normalizing forces the index to range from 0 to 1, with a higher index being more deprived.

#### Phecode

We ascertained phenome-wide clinical outcomes based on phecode^33,34^ (https://www.phewascatalog.org/phecodes, version 1.2) using ICD-9/ICD-10 codes curated from the AoU electronic health records. For a given subject, if any ICD codes from the inclusion criteria of a phecode were recorded, the subject was classified as a case. Conversely, if none of these ICD codes were recorded, the subject was classified as a control. If the number of cases is greater than 10, association with LTL was tested by logistic regression model, and heterogeneity of the association was tested by Cochran’s Q test, both with Bonferroni correction.

#### Hematologic cancer, non-hematologic cancer, clonal hematopoiesis of indeterminate potential (CHIP), and coronary artery disease (CAD)

Hematologic cancer, non-hematologic cancer, and CAD were defined by any of the recorded International Classification of Diseases (ICD9 and ICD10), Current Procedural Terminology 4 (CPT4), Healthcare Common Procedure Coding System (HCPCS), and Diagnosis Related Group (DRG) codes, and SNOMED terminology listed in Supplementary Tables 13-15. CHIP is ascertained in AoU as described before^81^. Briefly, Mutect2^82^ detected the putative somatic mutations using WGS data. Mutations with minimal allelic depth of less than 5 were excluded to reduce false positives. Then, the putative variants were cross-referenced with the previously identified CHIP-related variants^83^.

### Geographic analysis

Coefficients for LTL were calculated for each 3-digit prefix of ZIP codes by the linear regression model, adjusting with age, sex, first 10 genetic PCs, and sequencing site. The ZIP codes starting with 000 were excluded from the analyses (n = 147). The individuals with discordant state and ZIP code information were also excluded from the analyses. For cluster analysis, the coefficient was weighted by inverse variance and analyzed with SaTScan version 10.1.3^84^ with a 1,000-kilometer restriction of cluster size. Figures were plotted using R package sf^85^ and ggplot2^86^, excluding groups with less than 21 participants in each granularity to align with the publishing policy of AoU. The shape files and population information were downloaded from the U.S. Census Bureau (https://www.census.gov/).

### Genetic ancestry ascertainment

Genetically similar population groups were predicted by Data and Research Center (DRC) using random forest probability > 0.75 calculated with the first 16 genetic PCs. Briefly, 151,159 high-quality sites, which can be called accurately in both the training set (Human Genome Diversity Project and 1000 Genomes)^87,88^ and target data (AoU short read WGS), were identified by autosomal and bi-allelic single nucleotide variants only, allele frequency (AF) > 0.1%, call rate > 99%, and linkage disequilibrium (LD)-pruned with a cutoff of *R*^2^ = 0.1 similarly with gnomAD^89^. Genetic PCs were then derived using the hwe_normalized_pca in Hail at this high-quality variant sites. A random forest classifier was trained on the training set using variants obtained from gnomAD on the autosomal exons of protein-coding transcripts in GENCODE v42^90^. The AoU samples were projected into the PCA space of the training data, and the classifier was applied.

### Genotype-based sample filtering

All the samples flagged by AoU CDR and GC due to genotype heterogeneity were excluded from all analyses (n = 549). Briefly, eight median absolute deviations away from the median residuals in any of the following metrics were excluded after residualization by the first 16 genetic PCs: number of deletions, number of insertions, number of single nucleotide polymorphisms, number of variants not present in gnomAD 3.1, insertion to deletion ratio, transition to transversion ratio, and heterozygous to homozygous ratio (single nucleotide polymorphisms and Indel separately). We further excluded samples with variant calling missingness over 5%, without information on genetically estimated sex ploidy by the DRAGEN pipeline, and missing in the array genotype file provided by AoU at the time of analysis (December 2023).

### Genotype and variant quality control

#### Genotyping array

We use array-based genotypes provided by AoU for the Regenie Step 1 and by UKB for both Steps 1 and 2. Variants with missingness > 1%, minor AF < 1%, and Hardy-Weinberg Equilibrium mid-p adjusted p-value < 1×10^6^ were excluded in each genetic ancestry group. Variants were then pruned with variant window 1,000 kb, variant sliding window 100 kb, and *R*^2^ 0.5. In addition, we excluded variants located in high linkage disequilibrium regions^91^.

#### WGS in AoU

We used the WGS-based genotype provided by AoU for the Regenie Step 2. AoU CDR and GC marked genotypes that were low-quality or failed allele-specific variant quality score recalibration calculated by GATK^92^. We filtered out those genotypes in our analysis and split multi-allelic variants into biallelic by Hail (https://github.com/hail-is/hail). Variants were filtered out by PLINK2.00 if missingness > 0.05 or Hardy-Weinberg Equilibrium *P* < 1×10^6^ with mid-p adjustment in each genetic ancestry. We further excluded variants located in low-complexity regions and segmental duplications with over 95% similarity. The numbers of included variants are listed in Supplementary Table 16.

#### WES in UKB

For genotype-level quality control, we first used Hail’s split_multi_hts function to divide multiallelic sites and filtered out low-quality genotypes; genotyping quality ≤ 20, depth ≤ 10 or > 200, (DP_Reference_ + DP_Alternate_)/(DP_Total_) > 0.9 and DP_Alternate_/DP_Total_ > 0.2 for heterozygous genotypes, DP_Alternate_/DP_Total_ > 0.9 for alternate homozygous genotypes. This process retained 26,645,535 variants in 454,756 samples. We excluded 6,289,813 variants due to high missingness (>10%), Hardy-Weinberg deviation (P_HWE_ < 1×10^15^), or low-complexity regions, leaving 20,355,722 variants.

### GWAS for common variant

We followed the previously recommended fully adjusted two-step procedure^93^. We residualized the estimated LTL by age, sex (imputed from genotype by DRAGEN pipeline provided by AoU Genome Center [GC] and DRC), the first 10 PCs calculated from genotype (provided by AoU GC and DRC), the PCs from NGS-PCA^24^ (https://github.com/PankratzLab/NGS-PCA), and sequencing site in each genetic ancestry group. We included all the NGS PCs, explaining at least 0.1% of the total calculated variance (the number of PCs included is shown in Supplementary Table 11). Regenie^94^ Step 1 was performed with blocks of sizes 1,000 for LD computation, adjusting for age, sex, first 10 genetic PCs, sequencing site, and NGS-PCs explaining > 0.1% variance. LTL was rank normalized using --apply-rint option. Regenie Step 2 was performed by adjusting for the same variable in Step 1 for the WGS-based genotype with larger than 0.1% of minor AF. LD score regression was performed using 1000G WGS-derived LD scores to assess inflation (Supplementary Table 3) and calculate the genetic correlation in EUR between AoU and UKB. We applied genomic control by multiplying the standard error by the square root of the LD Score regression intercept and recalculated *P* values accordingly. We defined the genome-wide significance threshold as 5×10^8^. An independent locus was defined if the significant variants were more than 1 Mb apart. Novel locus was defined if the lead or sentinel variants reported in the previous 17 GWAS studies^4,19,21,24,25,48,51,95–104^ (listed in Supplementary Table 17) were more than 1 Mb apart from all the significant variants in the locus. Additionally, we investigated the summary statistics from LTL GWAS in UKB and revised the significant loci to align the definition of significance in AoU. We excluded the loci that overlapped with the aforementioned definition with these summary statistics from novel definitions, in addition to excluding previously reported sentinel variants in UKB^4^.

### GWAS Meta-analysis with UKB

We performed GWAS for each genetic ancestry (random forest probability > 0.75 using genetic PCs) with NHLBI Trans-Omics for Precision Medicine (TOPMed) imputed genotype aligned to GRCh38, which aligned the method with the GWAS in AoU. We used GWAMA^105^ in each genetic ancestry group and filtered out the variants supported by only one cohort. We excluded the variants with high heterogeneity (*P* < 10^6^) from the locus definition, which excluded *HBB* locus in line with the previously suspected artifact at this locus derived from the qPCR control used in UKB^4^. LD Score regression intercept and ratio were calculated to confirm the controlled genome-wide test statistics (Supplementary Table 6). We further performed a multi-ancestry meta-analysis using MR-MEGA^106^ with three PCs as previously performed^107^.

### Rare variant association study

We used the same condition for Regenie Step1 as the common variant association study. Step 2 was performed with alternative AF less than 0.1%. To prioritize deleterious missense variants, we generated a missense score using 30 functional annotations available in dbNFSP (version 4.2), as previously described^108^. Briefly, we predicted the deleteriousness of missense variants using 30 (MetaRNN_pred, SIFT_pred, SIFT4G_pred, Polyphen2_HDIV_pred, Polyphen2_HVAR_pred, LRT_pred, MutationTaster_pred, MutationAssessor_pred, FATHMM_pred, PROVEAN_pred, VEST4_rankscore, MetaSVM_pred, MetaLR_pred, M-CAP_pred, REVEL_rankscore, MutPred_rankscore, MVP_rankscore, MPC_rankscore, PrimateAI_pred, DEOGEN2_pred, BayesDel_addAF_pred, BayesDel_noAF_pred, ClinPred_pred, LIST-S2_pred, CADD_phred, DANN_rankscore, fathmm-MKL_coding_pred, fathmm-XF_coding_pred, Eigen-phred_coding, Eigen-PC-shred_coding) annotations and computed the proportion of deleterious predictions (missense score). We then created masks for aggregation tests using predicted loss-of-function variants and missense variants with missense scores higher than 0.6. Variants were annotated using VEP version 105. Multi-ancestry meta-analyses were performed using GWAMA with a fixed effect inverse variance weighted method.

### LD-score regression

Genetic correlations and LD Score regression intercept and ratio were calculated by using LDSC software version 1.0.1^39^. LD scores were computed using the 1000 Genomes Project phase 3 release data and variants in the HapMap3 project^109^ with minor allele count ≧ 5.

### Polygenic prioritization of candidate causal genes

We implemented PoPS (v0.2)^41^, a similarity-based gene prioritization method designed to leverage the full genome-wide signal to nominate causal genes independent of methods utilizing GWAS data proximal to the gene. PoPS leverages polygenic enrichments of gene features, including cell-type-specific gene expression, curated biological pathways, and protein-protein interaction networks to train a linear model to compute a PoPS score for each gene. We used the EUR reference panel for multi-ancestry summary statistics, while genetic ancestry-specific analyses leveraged genetic ancestry-specific reference panels. The analysis was limited to autosomes.

### Finemapping

LD-informed statistical finemapping by FINEMAP (version 1.4.2)^57^ was conducted using the results from genetic ancestry-wise meta-analysis with LD information with matched genetic ancestry in AoU. Finemapping was restricted to the variants with *P* < 0.05 to reduce the number of variants included. The LD matrices were computed using WGS data in AoU with LDstore2 (version 2.0)^57^ software. Finemapping was conducted by FINEMAP software using default prior probability and other conditions except for using a maximum number of causal variants as 10. We used marginal posterior inclusion probability as a measure of causal probability of the variants. We excluded loci detected in the human major histocompatibility complex region and chromosome X from the finemapping.

### Polygenic score (PGS)

For epidemiological analyses, we calculated PGS in EUR in AoU. Variant weights were calculated by GWAS in EUR in UKB using LDpred2^110^ software. We selected the best parameters using randomly selected 2,000 individuals from EUR in AoU, which were excluded from the subsequent analyses. PGS for the rest of EUR individuals in AoU were computed using PLINK2 software.

The performance of PRS was evaluated by the partial *R*^2^.

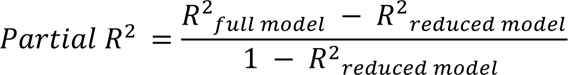

The model was adjusted by age, age^2^, sex, the first 10 genetic PCs, and sequencing site.

For the performance evaluation after meta-analysis, we conducted GWAS in the AoU cohort separately in AFR and EUR, leaving 10,000 randomly selected individuals in each genetic ancestry to avoid sample overlap. We then meta-analyzed these results with those from the UKB GWAS for respective genetic ancestries, creating meta-analysis results. The weights for polygenic scoring were derived from this genetic ancestry-specific meta-analysis and optimized using LDpred2^110^ software. PGS for the holdout samples were computed using PLINK2 software. We selected the best parameters using randomly selected 2,000 individuals from holdout individuals and tested performance on the remaining 8,000 holdout individuals. The performance of PRS was evaluated by the partial *R*^2^ as described above. The parameters selected and performance metrics for each PGS were summarized in Supplementary Table 10. The distribution of original PGSs before standardization is shown in Supplementary Fig. 8.

### Heritability estimation

We performed GCTA GREML^111^ (GCTA version 1.94.2) using 10,000 random unrelated samples at most in each genetic ancestry group with variants minor AF > 1%. The model was adjusted with the same covariates included in the GWAS studies.

### Software

FINEMAP 1.4.2, GCTA version 1.94.2, GWAMA ver. 2.2.2, Hail 0.2, LDSC v1.0.1, LDstore 2.0, MAGMA v1.10, mosdepth v0.3.5, MR-MEGA ver. 0.2, NGS-PCA v.0.0.2 (modified), PLINK 2.00 (Stable beta 7, 16 Jan), PoPS v0.2, R 4.2 (R packages: ggplot2 version 3.5.1, locuszoomr version 0.3.5, metafor version 3.8-1, sf version 1.0-15, qqman version 0.1.9), Regenie v3.4, SaTScan version 10.1.3, TelSeq v0.0.2 (modified).

## Acknowledgment

We acknowledge the studies and participants who provided biological samples and data for AoU and UKB. UKB Resource was used under application number 7089. Secondary use of the UKB data was approved by the Massachusetts General Hospital institutional review board (2021P002228). T.N. is supported by the National Heart Lung Blood Institute (K99HL165024). S.K. is supported by the National Heart Lung Blood Institute (K99HL169733). A.P.P is supported by the National Heart Lung Blood Institute (K08HL168238). P.N. is supported by the National Heart Lung Blood Institute (R01HL168894).

## Data Availability

GWAS summary statistics generated in this study will be publicly available at Cardiovascular Disease Knowledge Portal (https://cvd.hugeamp.org/) upon publication. The estimated LTL will be returned to AoU upon publication. The genotypes and phenotypes of UKB and AoU participants are available by application to the UKB (https://www.ukbiobank.ac.uk/register-apply/) and AoU (https://allofus.nih.gov/), respectively. The previously published GWAS summary statistic for LTL in UKB (https://figshare.com/s/caa99dc0f76d62990195) and meta-analysis (https://figshare.com/s/f6de1a56ad7c448c1f4c) are publicly available. The previously published full GWAS summary statistics for LTL in TOPMed are available upon application on dbGap (phs001974).

## Code Availability

Modified TelSeq and NGS-PCA will be available in GitHub upon publication. The codes used for the standard analyses will be available at Zenodo upon publication.

## Figures and Legends

**Extended Data Fig. 1:**
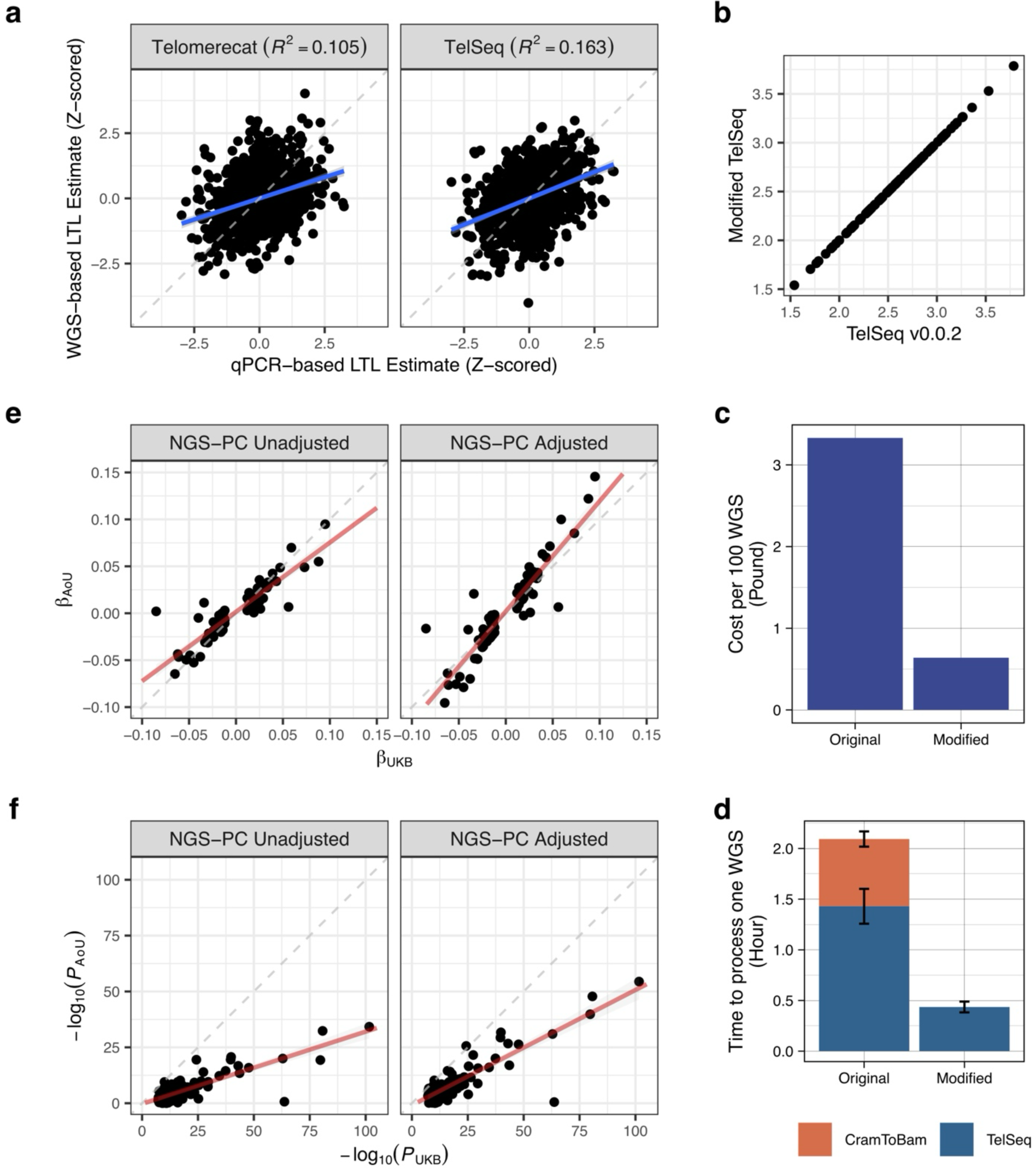
Telomere length estimation using whole genome sequence data. **a**, Telomerecat and TelSeq were compared for the agreement with quantitative polymerase chain reaction measurements within the same random 1,000 samples in the UK Biobank (UKB) for leukocyte telomere length (LTL) estimation using whole genome sequence (WGS) data. *R*^2^ was calculated by Pearson correlation. **b**, Estimated LTL was compared between modified TelSeq vs original TelSeq (v0.0.2) for random 1,000 samples in UKB. **c** and **d**, Modified TelSeq software was compared with original software for time (**c**) and cost (**d**) for random 100 WGS on UKB-RAP. **e** and **f**, The effect estimates (**e**) and *P*-values (**f**) of the independent sentinel variants in previous UKB for LTL were compared with the *All of Us* Research Program (AoU) between original LTL call and LTL adjusted by WGS-depth derived principal components (NGS-PCs) in European-like population.

**Extended Data Fig. 2:**
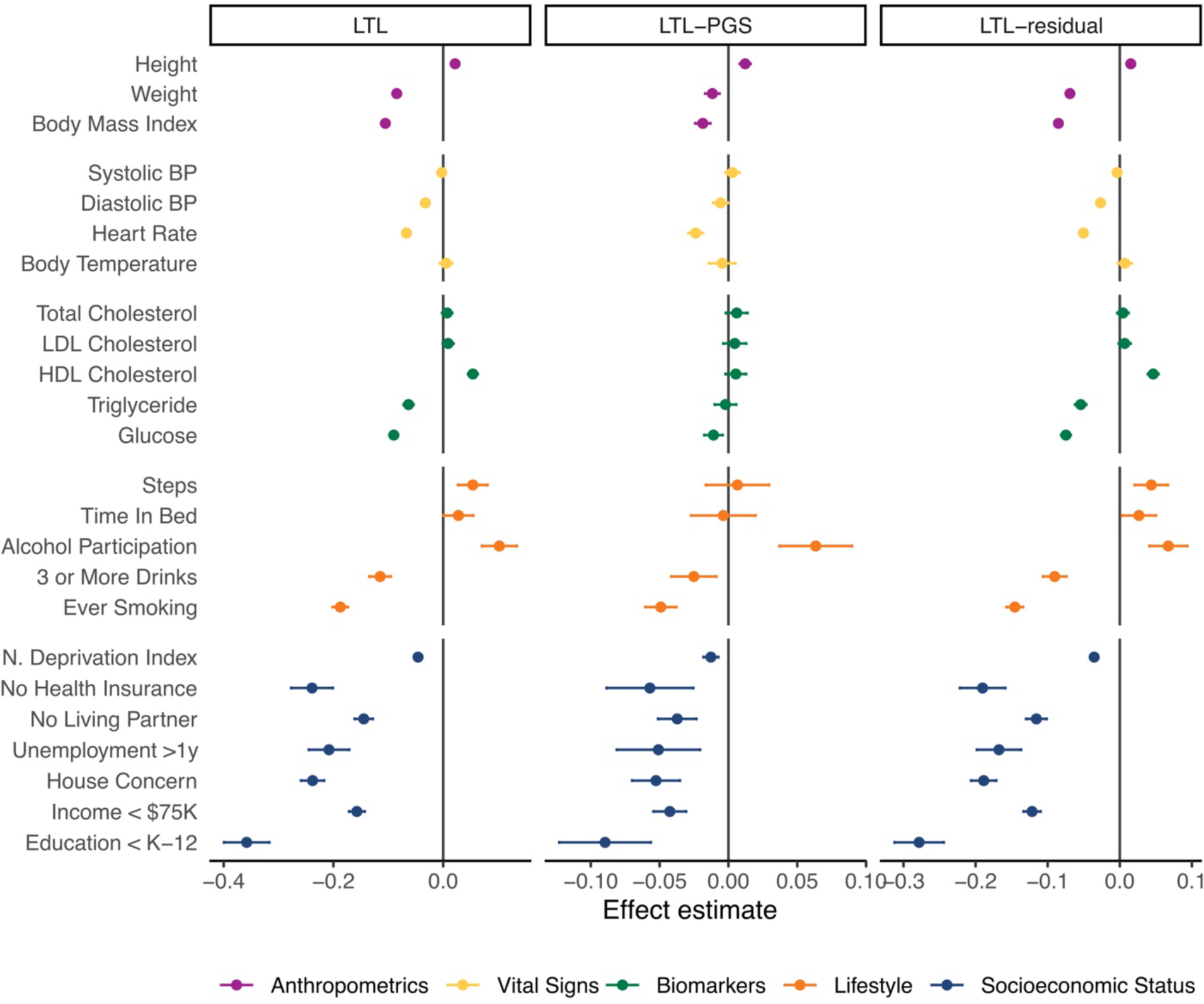
Associations between leukocyte telomere length (LTL)/LTL-polygenic score (PGS)/LTL-residual and health-related traits in European-like population in the *All of Us* Research Program. The effects of leukocyte telomere length (LTL), polygenic score for LTL (LTL-PGS), and LTL residualized by LTL-PGS (LTL-residual) on health-related traits were tested in European-like population using linear regression model (continuous trait) and logistic regression model (binary trait) adjusted for age, sex, the first 10 genetic principal components, and sequencing site. BP: blood pressure, LDL: low-density lipoprotein, HDL: high-density lipoprotein, N. Deprivation Index: neighborhood deprivation index.

**Extended Data Fig. 3:**
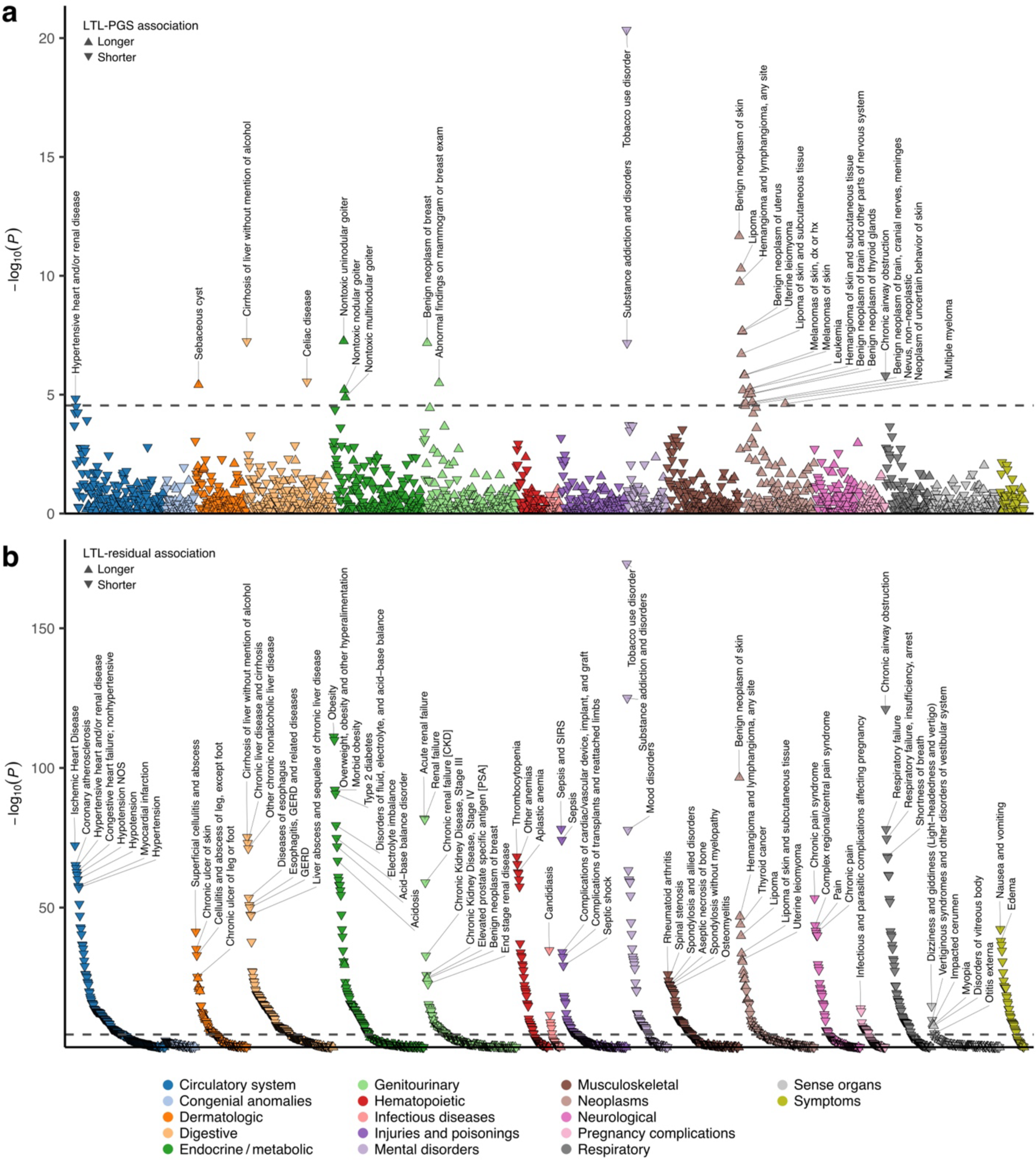
Phenome-wide association study of leukocyte telomere length-polygenic score (LTL-PGS) and LTL residualized by LTL-PGS (LTL-residual). Associations between polygenic score for LTL (LTL-PGS) (**a**) and LTL residualized by LTL-PGS (LTL-residual) (**b**) and prevalent phecodes were assessed by logistic regression model adjusting with age, sex, first 10 genetic principal components, and sequencing site in the *All of Us* Research Program. Upward triangles indicate positive associations, and downward triangles indicate negative associations. Phecodes with less than 10 cases were excluded from the analyses.

**Extended Data Fig. 4:**
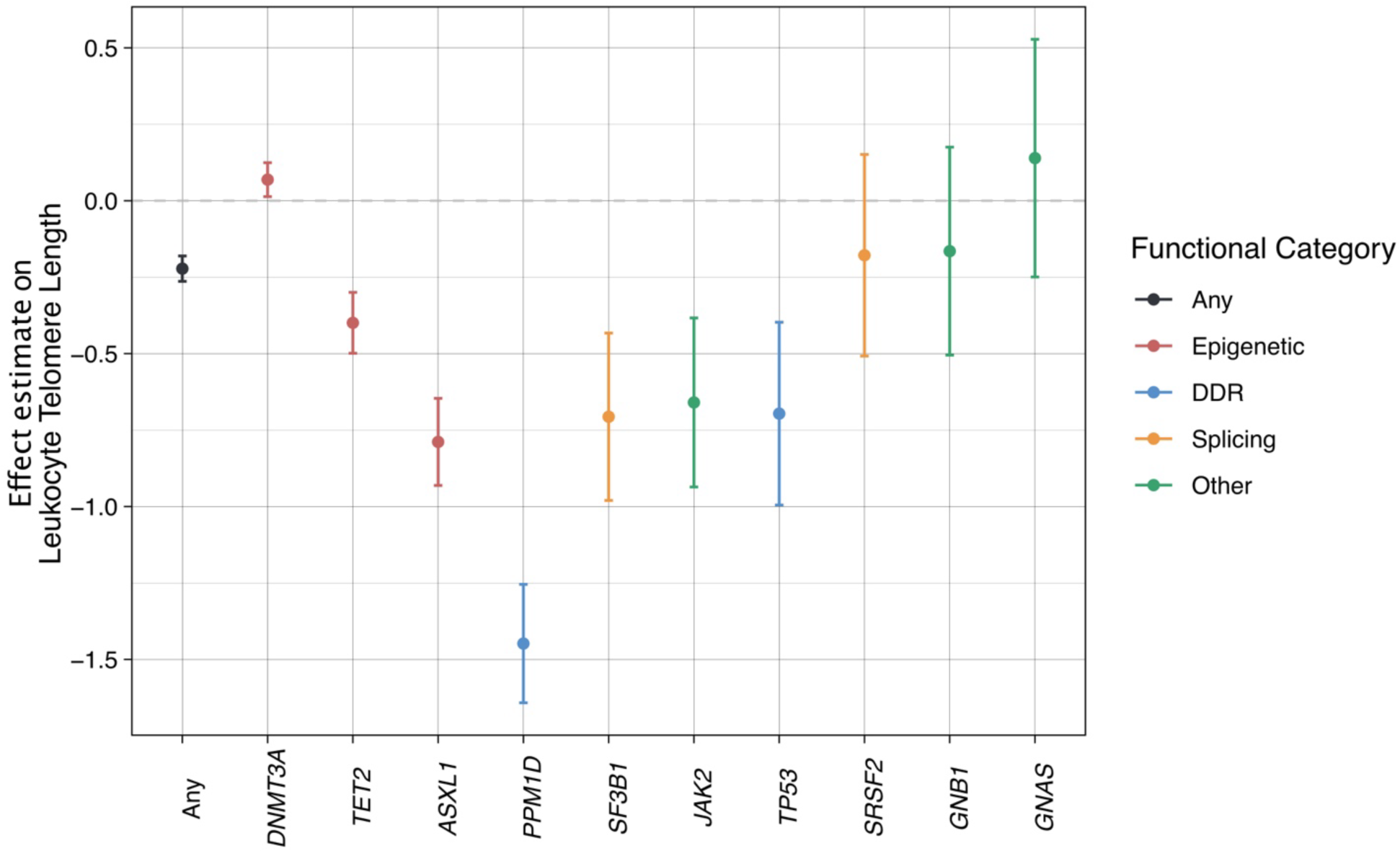
Association with clonal hematopoiesis of indeterminate potential (CHIP). Associations between leukocyte telomere length (LTL) and clonal hematopoiesis of indeterminate potential (CHIP) were assessed by logistic regression model adjusting with age, sex, first 10 genetic principal components, and sequencing site. DDR: DNA damage repair.

**Extended Data Fig. 5:**
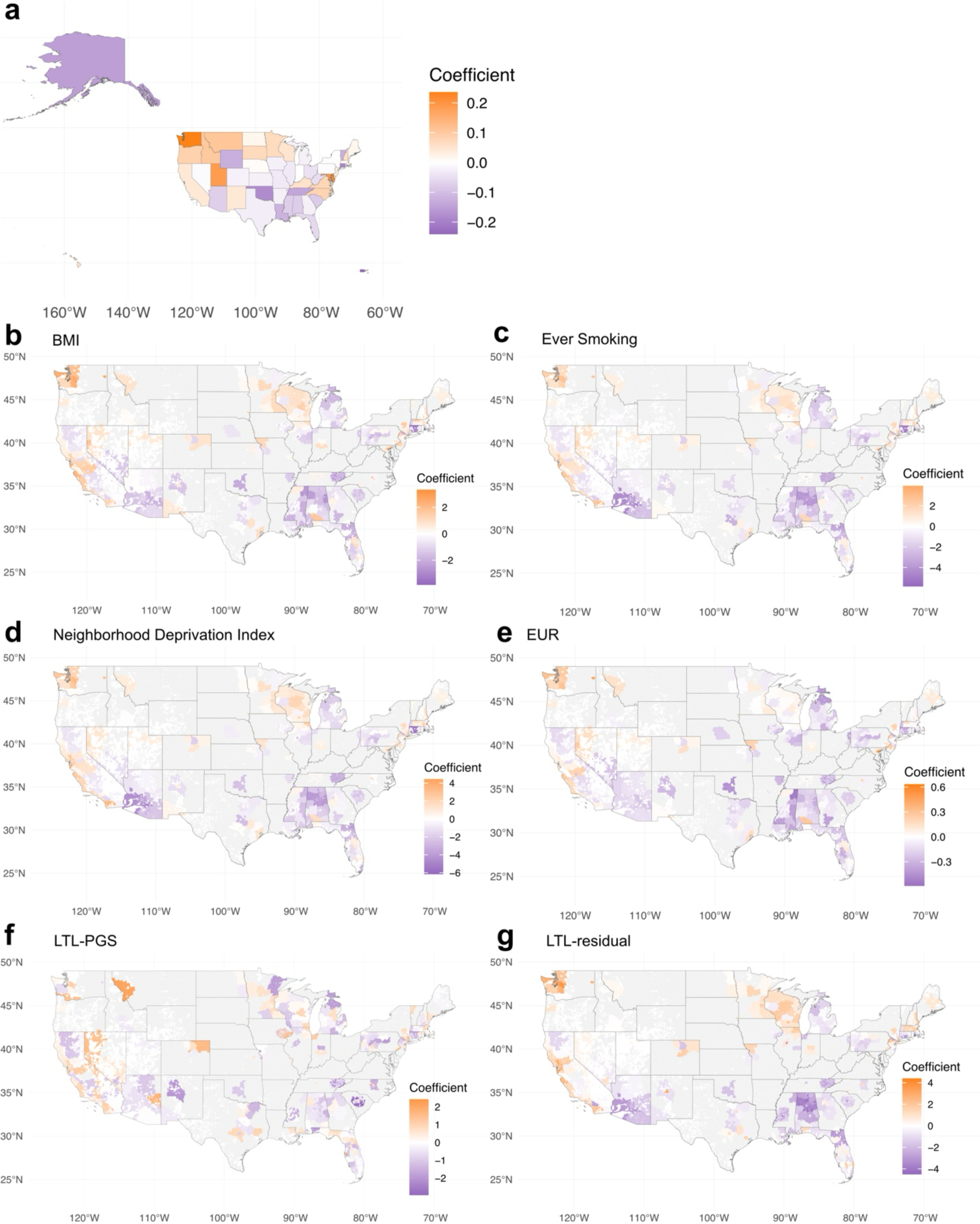
State-level heterogeneity of leukocyte telomere length (LTL) in the United States and sensitivity analyses of geographic heterogeneity. **a**, State-level heterogeneity of the leukocyte telomere length (LTL) across the United States was tested with linear regression model adjusting for age, sex, first 10 genetic principal components (PCs), and sequencing site. **b**-**d**, Geographical heterogeneity at 3 ZIP code prefix level was tested with linear regression model adjusted for additional covariates (**b**: body mass index [BMI], **c**: ever smoking, **d**: neighborhood deprivation index) in addition to age, sex, first 10 genetic principal components (PCs), and sequencing site. **e**-**g**, Geographical heterogeneity at 3 ZIP code prefix level was tested in European-like population (EUR) for LTL (**e**), LTL-PGS (**f**), and LTL-residual (**g**) with linear regression model adjusted for age, sex, first 10 genetic PCs, and sequencing site.

**Extended Data Fig. 6:**
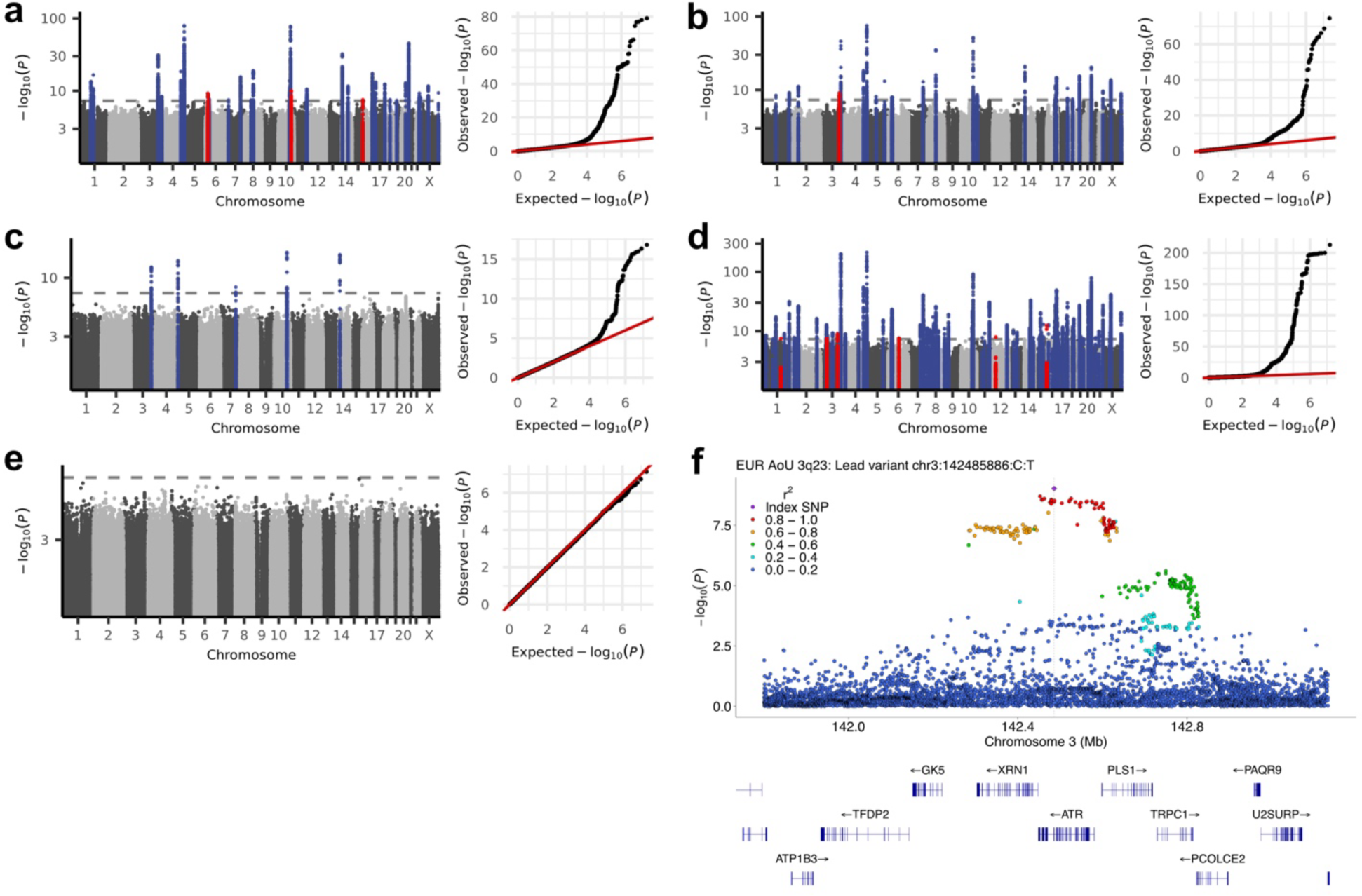
Genome-wide association study in the *All of Us* Research Program. Genome-wide association studies for common variants (minor allele frequency > 0.1%) were performed by Regenie^94^ adjusting for age, sex, the first 10 genetic principal components, and sequencing site in the *All of Us* Research Program separately for African-like population (AFR) (**a**), Admixed American-like population (AMR) (**b**), East Asian-like population (EAS) (**c**), European-like population (EUR) (**d**), and South Asian-like population (SAS) (**e**). Genomic control with linkage disequilibrium score regression intercept was applied. **f**, The locuszoom for a novel locus at 3q23 in EUR.

**Extended Data Fig. 7:**
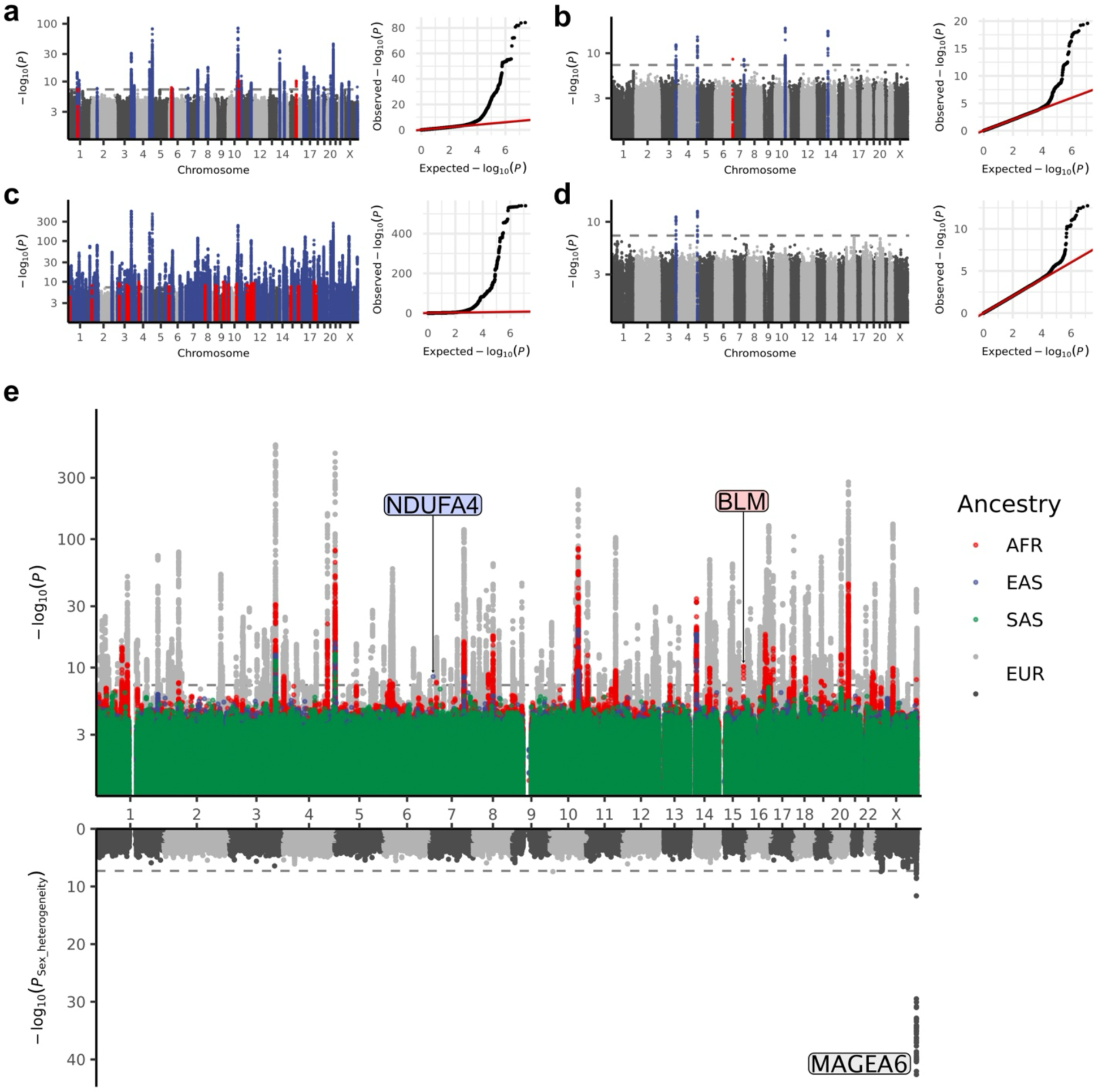
Genome-wide association study meta-analyses for leukocyte telomere length (LTL) with the *All of Us* Research Program and UK Biobank. Fixed effect meta-analyses for common variant genome-wide association studies (GWASs) were performed for the *All of Us* Research Program and UK Biobank, separately for African-like population (AFR) (**a**), East Asian-like population (EAS) (**b**), European-like population (EUR) (**c**), and South Asian-like population (SAS) (**d**). We only included variants found in both cohorts. Blue represents previously known significant loci (*P* < 5×10^8^), and red represents novel significant loci. **e**, Genetic ancestry-and sex-specific genomic predispositions were investigated in the GWAS meta-analyses. Manhattan plots for each genetic ancestry are plotted in different colors depending on the genetic ancestry as the upward part of the Miami plot. The heterogeneity *P* between sexes calculated by GWAMA is plotted downward in the Miami plot.

**Extended Data Fig. 8:**
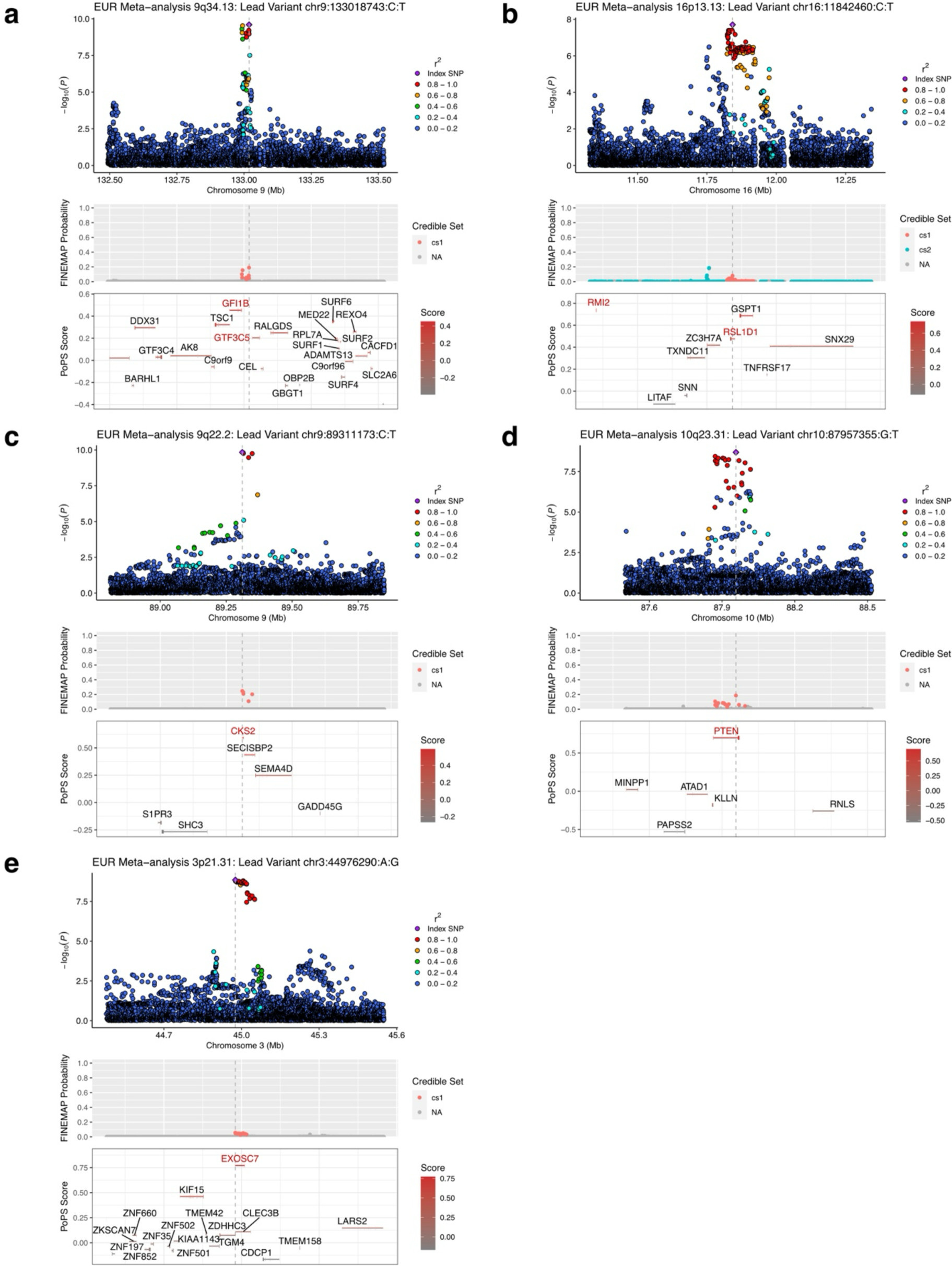
Locuszoom for a part of novel loci in meta-analysis. The locuszoom plots for example novel loci in genome-wide association study meta-analyses with FINEMAP^57^ and PoPS^41^ annotations. Red texts indicate the nearest and PoPS prioritized genes. EUR: European-like population.

**Extended Data Fig. 9:**
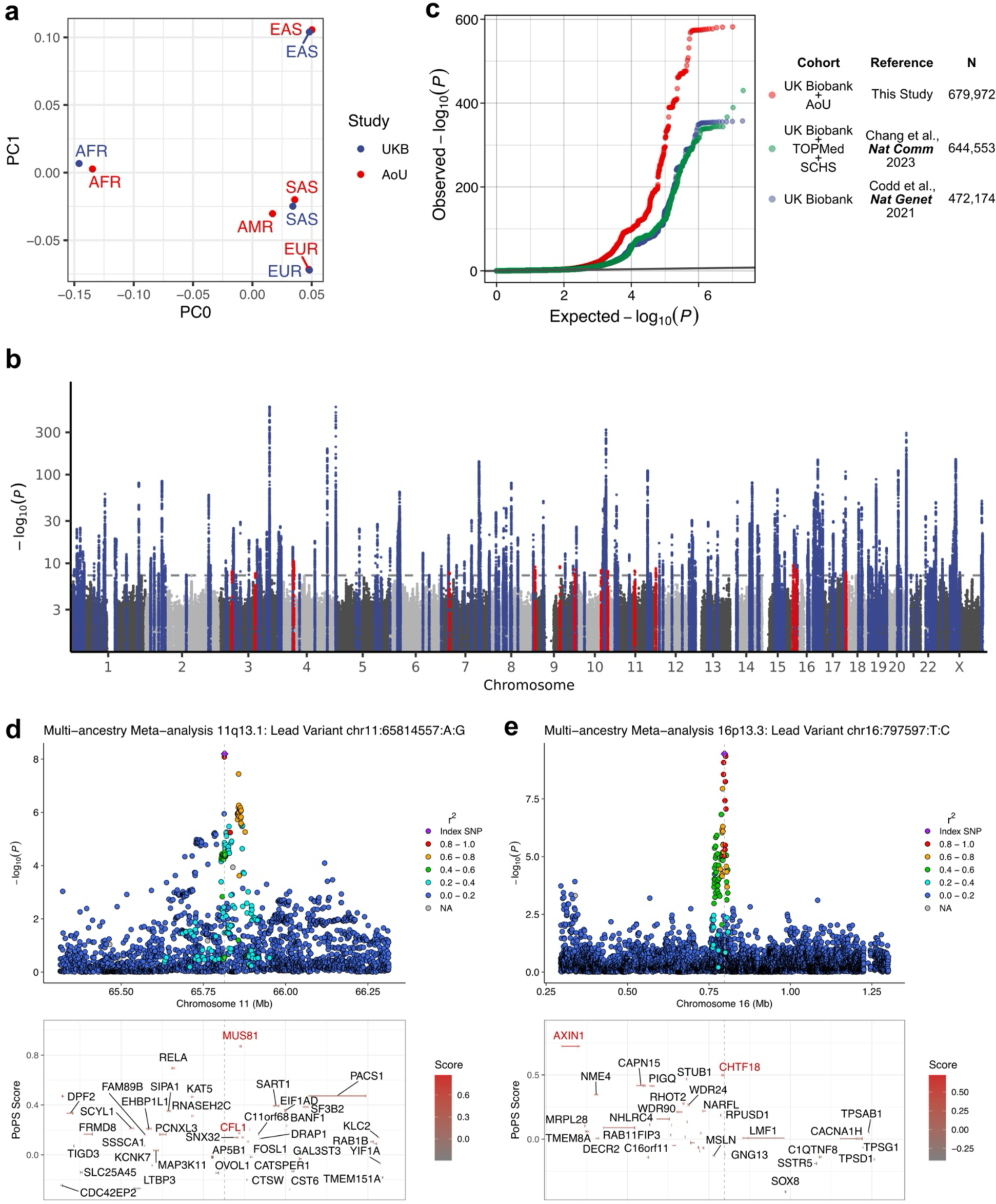
Multi-ancestry meta-analysis for common variant genome-wide association studies. **a**, Principal components calculated by MR-MEGA^106^ to correct the population structures for multi-ancestry genome-wide association study (GWAS) meta-analysis, which effectively distinguished genetic ancestries. **b** and **c**, We performed multi-ancestry GWAS meta-analysis for common variants in the *All of Us* Research Program (AoU) and UK Biobank (UKB). We implemented genomic correction by linkage disequilibrium score regression intercept before meta-analysis and found no evidence of inflation (λ_GC_ = 1.086). Manhattan plot (**b**) shows previously known significant (*P* < 5×10^8^) loci in blue and novel significant loci in red. QQ plot (**c**) of multi-ancestry GWAS in this study (red) and previous meta-analysis of UKB, TransOmics for Precision Medicine (TOPMed), and Singapore Chinese Health Study (SCHS)^51^(green) and UKB^4^ (blue). **d** and **e**, The locuszoom plots for the example novel loci in multi-ancestry GWAS meta-analysis for common variants. Red texts indicate the nearest and PoPS prioritized genes.

**Extended Data Fig. 10:**
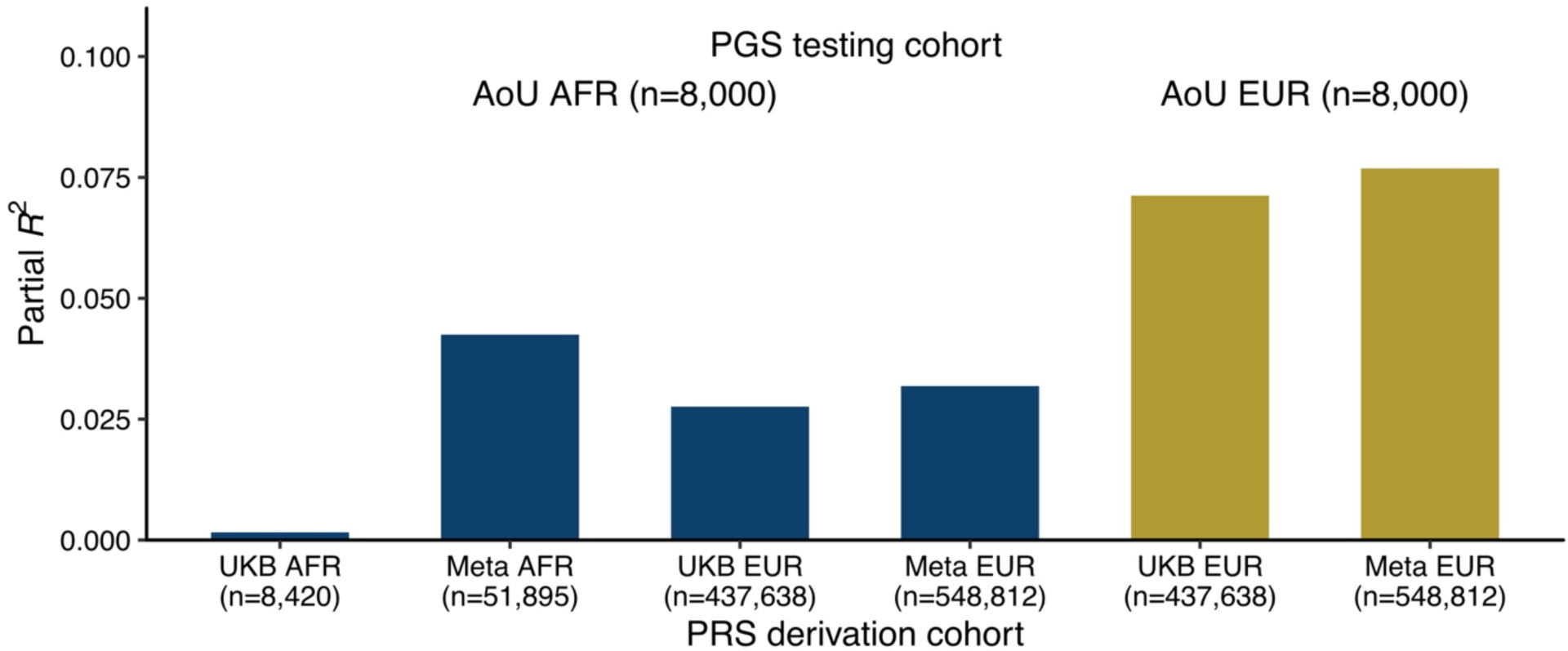
Improved performance of polygenic score in African-like population (AFR). We calculated polygenic scores (PGS) for leukocyte telomere length using LDpred2, with weights derived from training genetic ancestries in the respective cohorts. To do this, we performed genome-wide association studies (GWASs) in the All of Us Research Program (AoU), excluding a random sample of 10,000 individuals per genetic ancestry. The resulting summary statistics were used for meta-analysis with UK Biobank (UKB). We optimized hyperparameters based on the best-performing partial R2 in a random 2,000-sample subset held out from the GWAS. We then evaluated the final performance of the PGS (partial *R*^2^) in the remaining 8,000 held-out samples in AoU. AFR: African-like population, EUR: European-like population.

