## Supplementary Figures 1-4, 6-8 for "Genomic, phenomic, and geographic associations of leukocyte telomere length in the United States"

**Supplementary Fig. 1: Sample selection within the *All of Us* Research Program.**

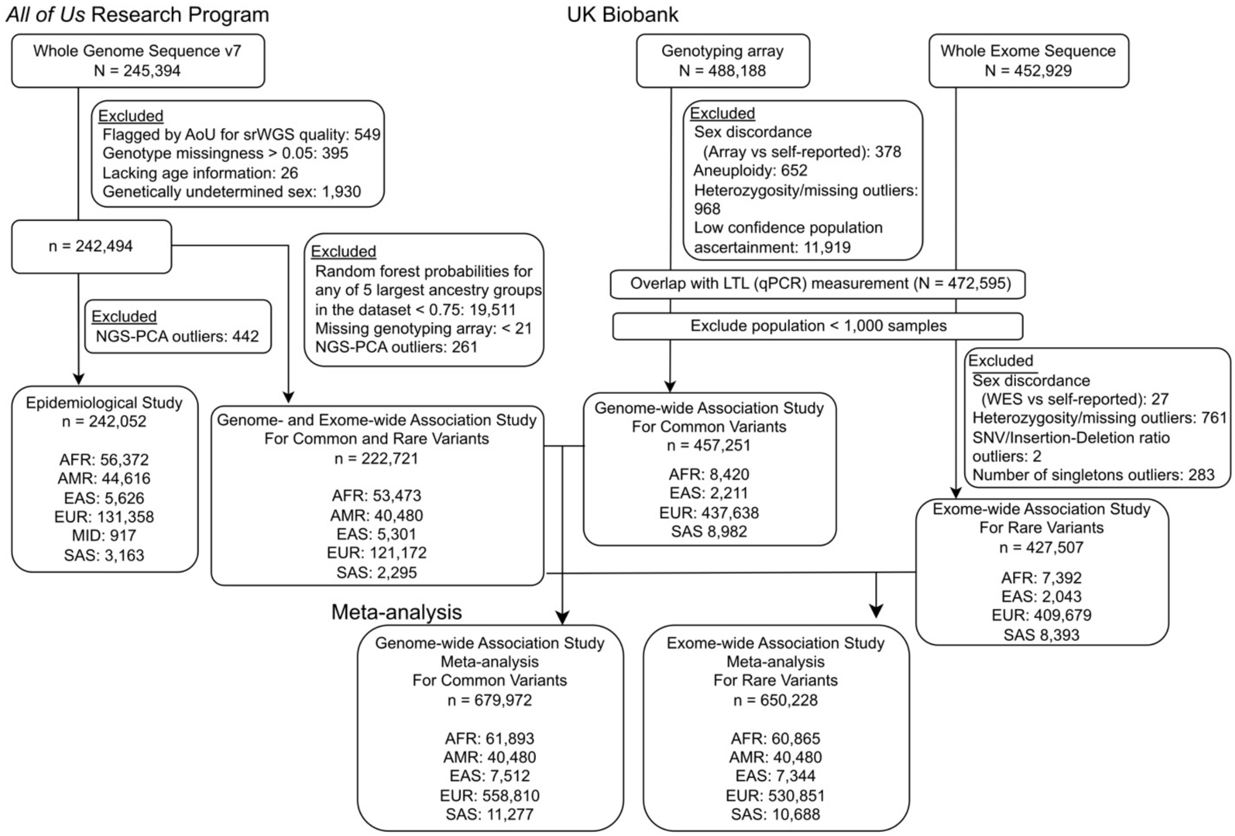


The *All of Us* Research Program (AoU) version 7 released 245,394 WGS data. We excluded flagged samples for sequencing quality by AoU (n = 549), genotyping missingness > 0.01 (n = 395), lacking age information (n = 26), and genetically undetermined sex (n = 1,930) to construct a cohort of 242,494 participants included in our analysis. For epidemiology, we further excluded outliers (> 20 median absolute deviations) for NGS-PCs (n = 442). For genomic studies, we further exclude < 0.75 for random forest probabilities for the largest 5 genetic ancestries [African-like population (AFR), Admixed American-like population (AMR), East Asian-like population (EAS), European-like population (EUR), and South Asian-like population (SAS)] in AoU (n = 19,511), missing genotyping array information (n < 21, According to AoU publishing policy, numbers below 21 cannot be published. Precise counts are not calculable due to overlapping samples across multiple QC metrics.), and outliers (> 20 median absolute deviation) for NGS-PCs (n = 261). We re-analyzed UK Biobank for genome-wide association study (GWAS) for common variants and exome-wide rare variant association study (RVAS) for meta-analyses. We used genotyping array data (N = 488,188) for GWAS and whole exome sequence data (N = 452,929) for RVAS.

**Supplementary Fig. 2: Associations between leukocyte telomere length (LTL) and phecodes and their heterogeneity in the *All of Us* Research Program.**

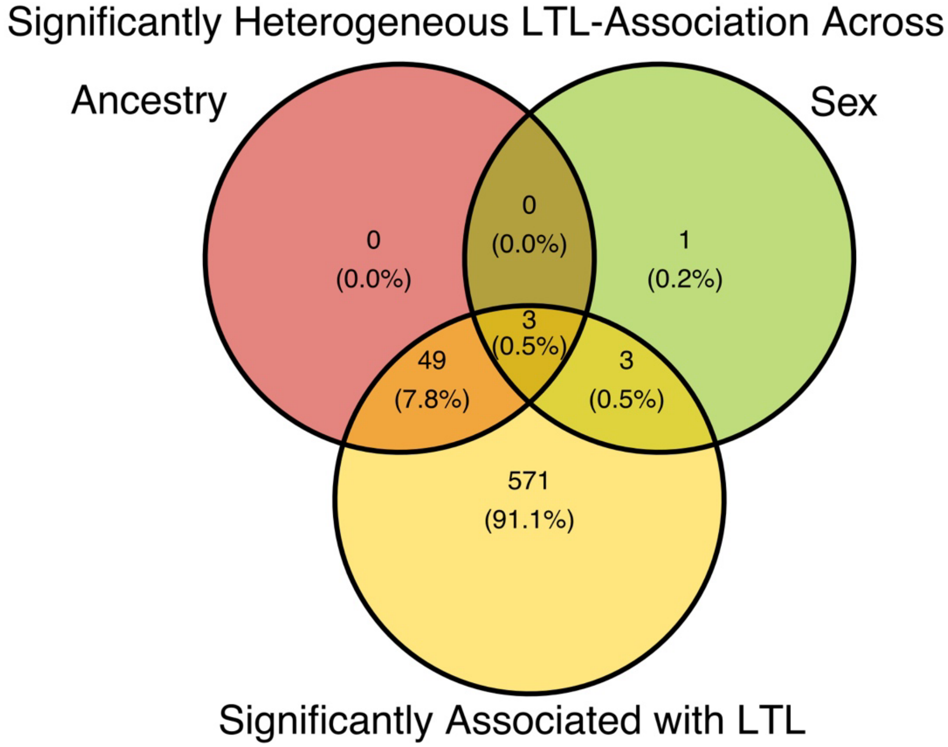


The Venn diagram for the phecodes that was significantly associated with leukocyte telomere length (LTL) (yellow) and those associations were significantly heterogeneous across genetic ancestries (red) and sexes (green) by Cochran's Q test with Bonferroni correction.

**Supplementary Fig. 3: Genome-wide association study for leukocyte telomere length (LTL) in the UK Biobank.**

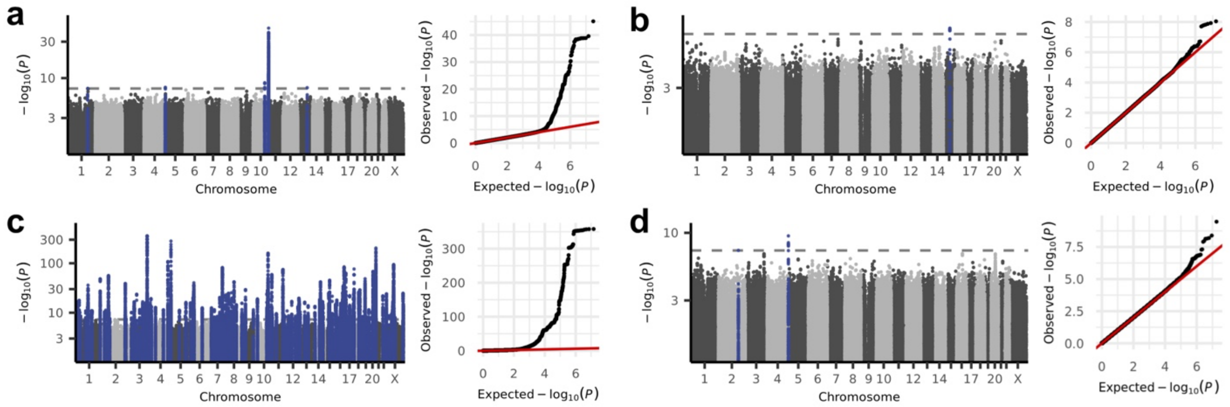


Genome-wide association studies for common variants were performed in the UK Biobank using previously published qPCR measurement of leukocyte telomere length[3] separately for African-like population (AFR) (**a**), East Asian-like population (EAS) (**b**), European-like population (EUR) (**c**), and South Asian-like population (SAS) (**d**). Genomic control with LD Score regression intercept was applied. Blue points in Manhattan plots represent significant loci ($P < 5 \times 10^{-8}$).

**Supplementary Fig. 4: Effect estimate comparison for leukocyte telomere length (LTL) between cohorts.**

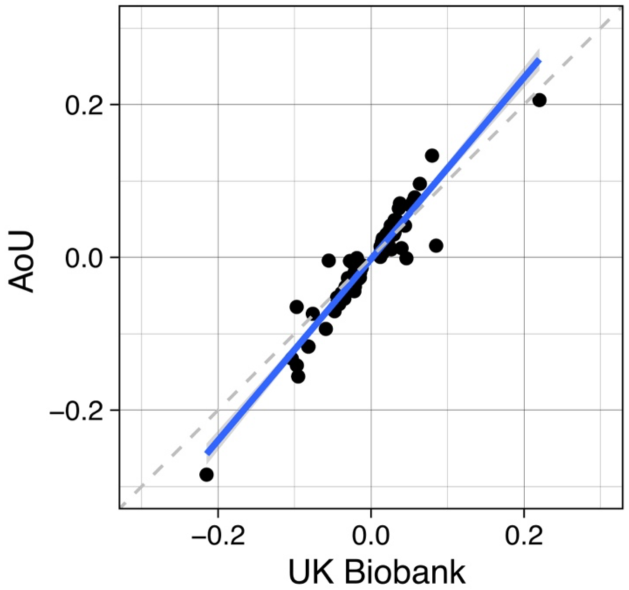


Effect estimates of lead variants in significant loci in the *All of Us* Research Program (AoU) were compared with the UK Biobank.

**Supplementary Fig. 5: Locuszoom plots with FINEMAP and PoPS annotations for significant loci in meta-analysis of common variant genome-wide association studies.**

A separate PDF is provided.

The locuszoom plots for significant loci in meta-analyses for the *All of Us* Research Program and UK Biobank common variant genome-wide association studies were plotted with FINEMAP[77] and PoPS[78] annotations if available. Red texts indicate the nearest and PoPS prioritized genes.

**Supplementary Fig. 6: Locuszoom plots for common variant genome-wide association study near the novel genes found in the rare variant association study.**

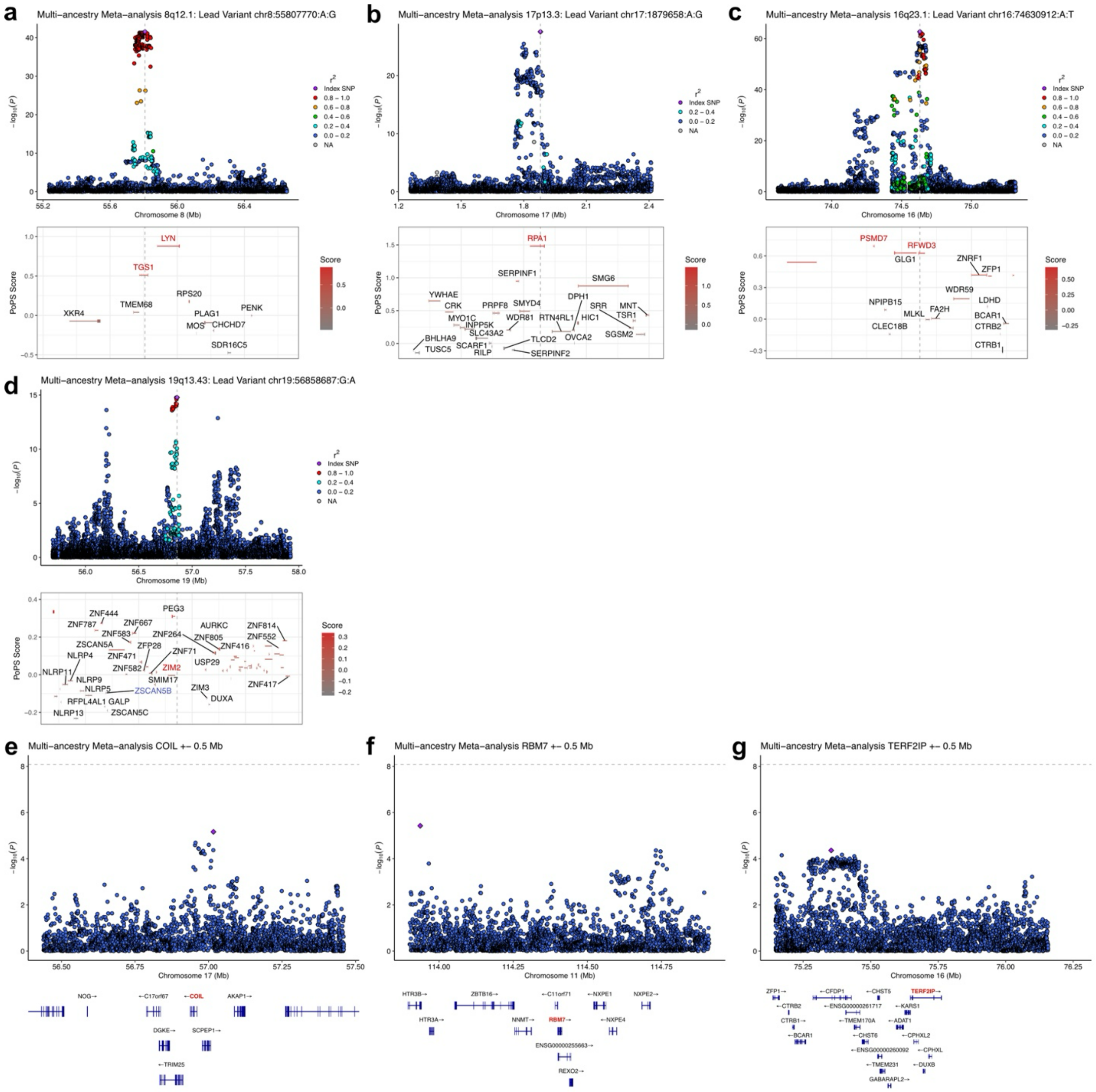


The locuszoom plots for common variant GWAS around the novel genes found in rare variant gene aggregation tests. If the locus overlaps with GWAS locus (**a**-**d**), PoPS scores were plotted below, and red texts indicate nearest and PoPS prioritized genes.

**Supplementary Fig. 7: Number of NGS-derived principal components used to adjust the estimated leukocyte telomere length.**

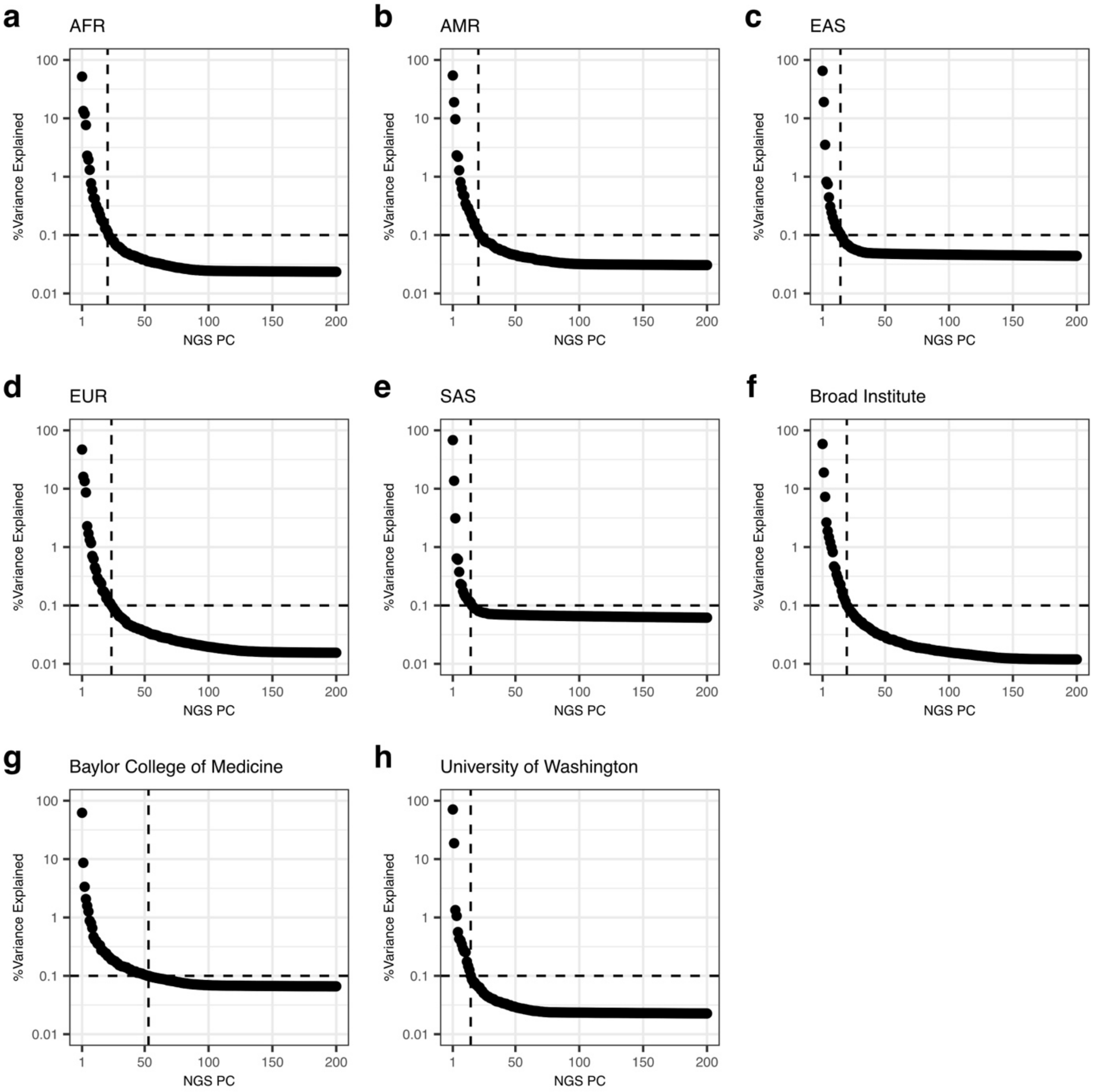


We calculated principal components (PCs) by modified NGS-PCA software[18] using sequencing depth for every 1,000 base-pairs across the genome calculated by mosdepth[22]. Calculations were separately done for sequencing center (for epidemiology) and genetic ancestry (for genomic studies) to reduce computational burden. PCs explaining over 0.1 % of the calculated total variance were used to adjust the leukocyte telomere length estimated by TelSeq[22].

**Supplementary Fig. 8: Distribution of polygenic score for leukocyte telomere length.**

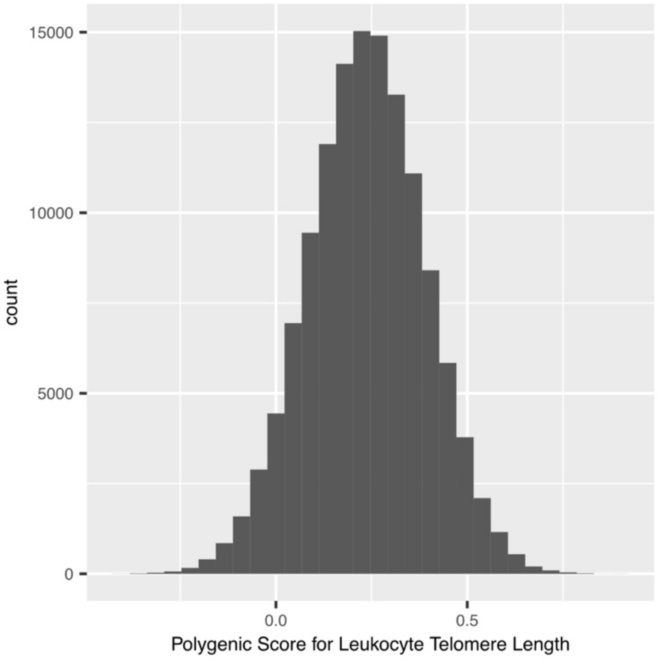


The distribution of the original polygenic score for leukocyte telomere length before standardization was plotted for European-like population in the *All of Us* Research Program.
