## Supplementary Figure 5 for "Genomic, phenomic, and geographic associations of leukocyte telomere length in the United States"

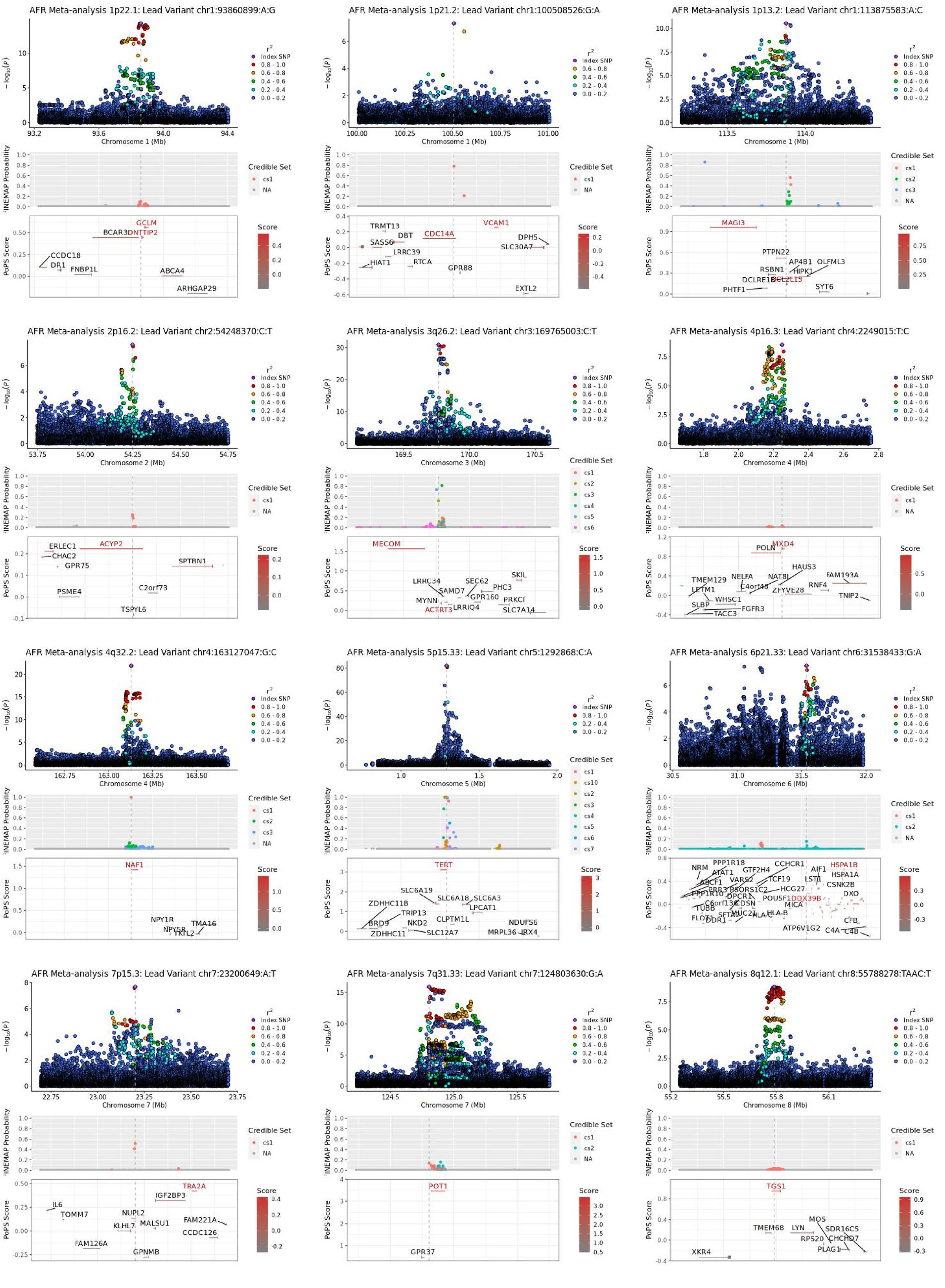

AFR Meta-analysis 1p22.1: Lead Variant chr1:93860899:A:G
Chromosome 1 (Mb)
GCLM
BCAR3
DNTTIP2
CCDC18
DR1
FNBP1L
ABCA4
ARHGAP29
AFR Meta-analysis 1p21.2: Lead Variant chr1:100508526:G:A
Chromosome 1 (Mb)
TRMT13
DBT
CDC14A
VCAM1
SASS6
DPH5
SLC30A7
LRRC39
HIAT1
RTCA
GPR88
EXTL2
AFR Meta-analysis 1p13.2: Lead Variant chr1:113875583:A:C
Chromosome 1 (Mb)
MAGI3
PTPN22
AP4B1
OLFML3
RSBN1
HIPK1
DCLRE1B
BCL2L15
PHTF1
SYT6
AFR Meta-analysis 2p16.2: Lead Variant chr2:54248370:C:T
Chromosome 2 (Mb)
ERLEC1
ACYP2
CHAC2
GPR75
SPTBN1
PSME4
C2orf73
TSPYL6
AFR Meta-analysis 3q26.2: Lead Variant chr3:169765003:C:T
Chromosome 3 (Mb)
MECOM
SKIL
LRRC34
SEC62
PHC3
SAMD7
GPR160
MYNN
PRKCI
ACTRT3
LRRIQ4
SLC7A14
AFR Meta-analysis 4p16.3: Lead Variant chr4:2249015:T:C
Chromosome 4 (Mb)
POLN
MXD4
HAUS3
FAM193A
TMEM129
NELFA
NAT8L
LETM1
C4orf48
ZFYVE28
RNF4
TNIP2
WHSC1
SLBP
FGFR3
TACC3
AFR Meta-analysis 4q32.2: Lead Variant chr4:163127047:G:C
Chromosome 4 (Mb)
NAF1
NPY1R
NPY5R
TMA16
TKTL2
AFR Meta-analysis 5p15.33: Lead Variant chr5:1292868:C:A
Chromosome 5 (Mb)
TERT
SLC6A19
SLC6A18
SLC6A3
ZDHHC11B
LPCAT1
TRIP13
BRD9
NKD2
CLPTM1L
NDUFS6
ZDHHC11
SLC12A7
MRPL36
IRX4
AFR Meta-analysis 6p21.33: Lead Variant chr6:31538433:G:A
Chromosome 6 (Mb)
NRM
PPP1R18
CCHCR1
AIF1
HSPA1B
ATAT1
GTF2H4
HSPA1A
ABCF1
VARS2
TCF19
LST1
CSNK2B
PPP1R10
PRR3
PSORS1C2
HCG27
DXO
TUBB
C6orf136
DPCR1
POU5F1
DDX39B
CDSN
MICA
FLOT1
SFTA2
MUC21
HLA-C
HLA-B
CFB
DDR1
ATP6V1G2
C4A
C4B
AFR Meta-analysis 7p15.3: Lead Variant chr7:23200649:A:T
Chromosome 7 (Mb)
TRA2A
IGF2BP3
IL6
TOMM7
NUPL2
MALSU1
FAM221A
KLHL7
CCDC126
FAM126A
GPNMB
AFR Meta-analysis 7q31.33: Lead Variant chr7:124803630:G:A
Chromosome 7 (Mb)
POT1
GPR37
AFR Meta-analysis 8q12.1: Lead Variant chr8:55788278:TAAC:T
Chromosome 8 (Mb)
TGS1
MOS
TMEM68
LYN
SDR16C5
RPS20
CHCHD7
PLAG1
XKR4
$-\log_{10}(P)$
FINEMAP Probability
PoPS Score
$r^2$
Index SNP
Credible Set
Score

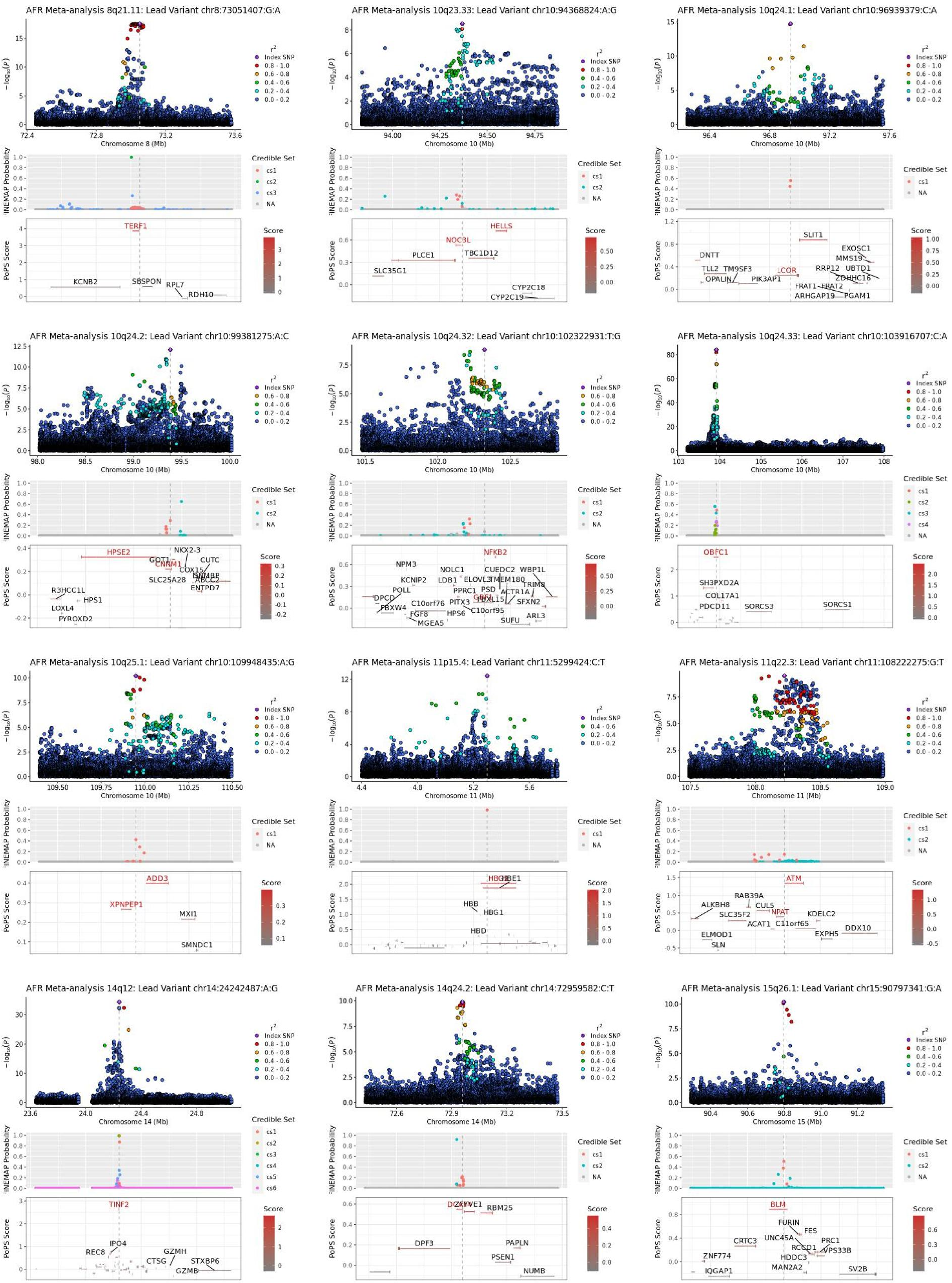
AFR Meta-analysis 8q21.11: Lead Variant chr8:73051407:G:A
−log10(p)
Chromosome 8 (Mb)
r²
Index SNP
0.8 - 1.0
0.6 - 0.8
0.4 - 0.6
0.2 - 0.4
0.0 - 0.2
FINEMAP Probability
Credible Set
cs1
cs2
cs3
NA
PoPS Score
Score
TERF1
KCNB2
SBSPON
RPL7
RDH10
AFR Meta-analysis 10q23.33: Lead Variant chr10:94368824:A:G
Chromosome 10 (Mb)
HELLS
NOC3L
PLCE1
TBC1D12
SLC35G1
CYP2C18
CYP2C19
AFR Meta-analysis 10q24.1: Lead Variant chr10:96939379:C:A
SLIT1
DNTT
EXOSC1
MMS19
UBTD1
RRP12
TLL2
TM9SF3
LCOR
ZDHHC16
OPALIN
PIK3AP1
FRAT1
FRAT2
ARHGAP19
PGAM1
AFR Meta-analysis 10q24.2: Lead Variant chr10:99381275:A:C
HPSE2
NKX2-3
GOT1
CNNM1
CUTC
COX15
GPNMB
SLC25A28
ABCC2
ENTPD7
R3HCC1L
HPS1
LOXL4
PYROXD2
AFR Meta-analysis 10q24.32: Lead Variant chr10:102322931:T:G
NFKB2
NPM3
NOLC1
CUEDC2
WBP1L
KCNIP2
LDB1
ELOVL3
TMEM180
TRIM8
POLL
PPRC1
PSD
ACTR1A
DPCD
C10orf76
PITX3
GBF1
FBXL15
SFXN2
FBXW4
C10orf95
FGF8
HPS6
SUFU
ARL3
MGEA5
AFR Meta-analysis 10q24.33: Lead Variant chr10:103916707:C:A
cs4
OBFC1
SH3PXD2A
COL17A1
PDCD11
SORCS3
SORCS1
AFR Meta-analysis 10q25.1: Lead Variant chr10:109948435:A:G
ADD3
XPNPEP1
MXI1
SMNDC1
AFR Meta-analysis 11p15.4: Lead Variant chr11:5299424:C:T
Chromosome 11 (Mb)
HBG2
HBE1
HBB
HBG1
HBD
AFR Meta-analysis 11q22.3: Lead Variant chr11:108222275:G:T
ATM
RAB39A
ALKBH8
CUL5
SLC35F2
NPAT
KDELC2
ACAT1
C11orf65
DDX10
ELMOD1
EXPH5
SLN
AFR Meta-analysis 14q12: Lead Variant chr14:24242487:A:G
Chromosome 14 (Mb)
cs5
cs6
TINF2
IPO4
REC8
GZMH
CTSG
STXBP6
GZMB
AFR Meta-analysis 14q24.2: Lead Variant chr14:72959582:C:T
DCAF4
ZFYVE1
RBM25
DPF3
PAPLN
PSEN1
NUMB
AFR Meta-analysis 15q26.1: Lead Variant chr15:90797341:G:A
Chromosome 15 (Mb)
BLM
FURIN
FES
CRTC3
UNC45A
PRC1
RCCD1
VPS33B
ZNF774
HDDC3
IQGAP1
MAN2A2
SV2B

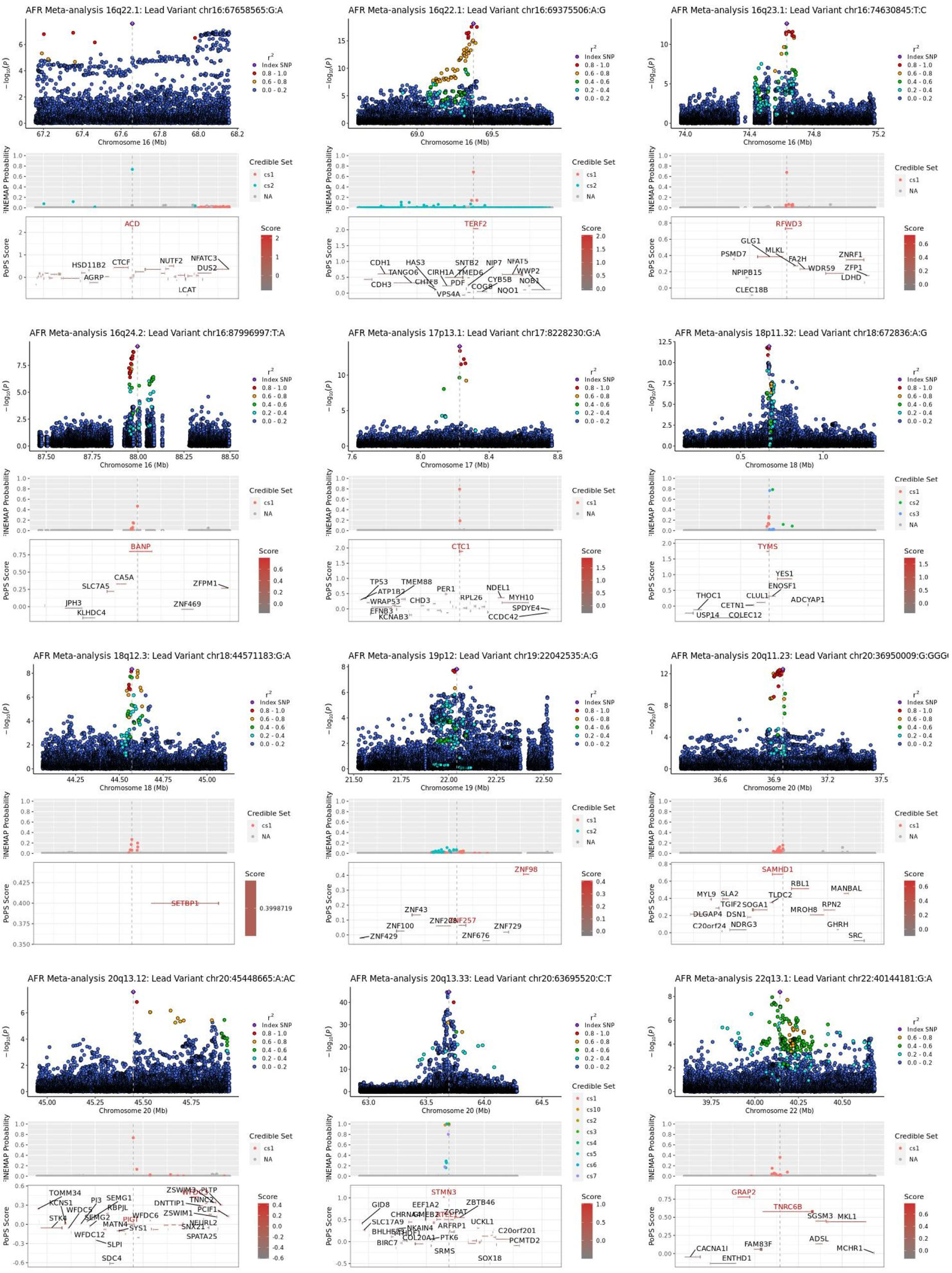
AFR Meta-analysis 16q22.1: Lead Variant chr16:67658565:G:A
ACD
HSD11B2
CTCF
NUTF2
NFATC3
DUS2
AGRP
LCAT
AFR Meta-analysis 16q22.1: Lead Variant chr16:69375506:A:G
TERF2
CDH1
HAS3
SNTB2
NIP7
NFAT5
TANGO6
CIRH1A
TMED6
WWP2
CDH3
CHTF8
PDF
CYB5B
NOB1
COG8
NQO1
VPS4A
AFR Meta-analysis 16q23.1: Lead Variant chr16:74630845:T:C
RFWD3
GLG1
PSMD7
MLKL
FA2H
ZNRF1
WDR59
ZFP1
NPIPB15
LDHD
CLEC18B
AFR Meta-analysis 16q24.2: Lead Variant chr16:87996997:T:A
BANP
CA5A
SLC7A5
ZFPM1
JPH3
ZNF469
KLHDC4
AFR Meta-analysis 17p13.1: Lead Variant chr17:8228230:G:A
CTC1
TP53
TMEM88
ATP1B2
PER1
NDEL1
WRAP53
CHD3
RPL26
MYH10
EFNB3
SPDYE4
KCNAB3
CCDC42
AFR Meta-analysis 18p11.32: Lead Variant chr18:672836:A:G
TYMS
YES1
ENOSF1
THOC1
CLUL1
ADCYAP1
CETN1
USP14
COLEC12
AFR Meta-analysis 18q12.3: Lead Variant chr18:44571183:G:A
SETBP1
0.3998719
AFR Meta-analysis 19p12: Lead Variant chr19:22042535:A:G
ZNF98
ZNF43
ZNF100
ZNF208
ZNF257
ZNF729
ZNF676
ZNF429
AFR Meta-analysis 20q11.23: Lead Variant chr20:36950009:G:GGG
SAMHD1
RBL1
MANBAL
MYL9
SLA2
TLDC2
TGIF2
SOGA1
RPN2
DLGAP4
DSN1
MROH8
C20orf24
NDRG3
GHRH
SRC
AFR Meta-analysis 20q13.12: Lead Variant chr20:45448665:A:AC
TOMM34
ZSWIM3
PLTP
KCNS1
PI3
SEMG1
TNNC2
DNTTIP1
WFDC5
RBPJL
PCIF1
STK4
SEMG2
WFDC6
ZSWIM1
MATN4
PIGT
NEURL2
WFDC12
SYS1
SNX21
SPATA25
SLPI
SDC4
AFR Meta-analysis 20q13.33: Lead Variant chr20:63695520:C:T
STMN3
ZBTB46
GID8
EEF1A2
ZGPAT
SLC17A9
UCKL1
NKAIN4
ARFRP1
C20orf201
BIRC7
COL20A1
PTK6
PCMTD2
SRMS
SOX18
AFR Meta-analysis 22q13.1: Lead Variant chr22:40144181:G:A
GRAP2
TNRC6B
SGSM3
MKL1
ADSL
CACNA1I
FAM83F
MCHR1
ENTHD1

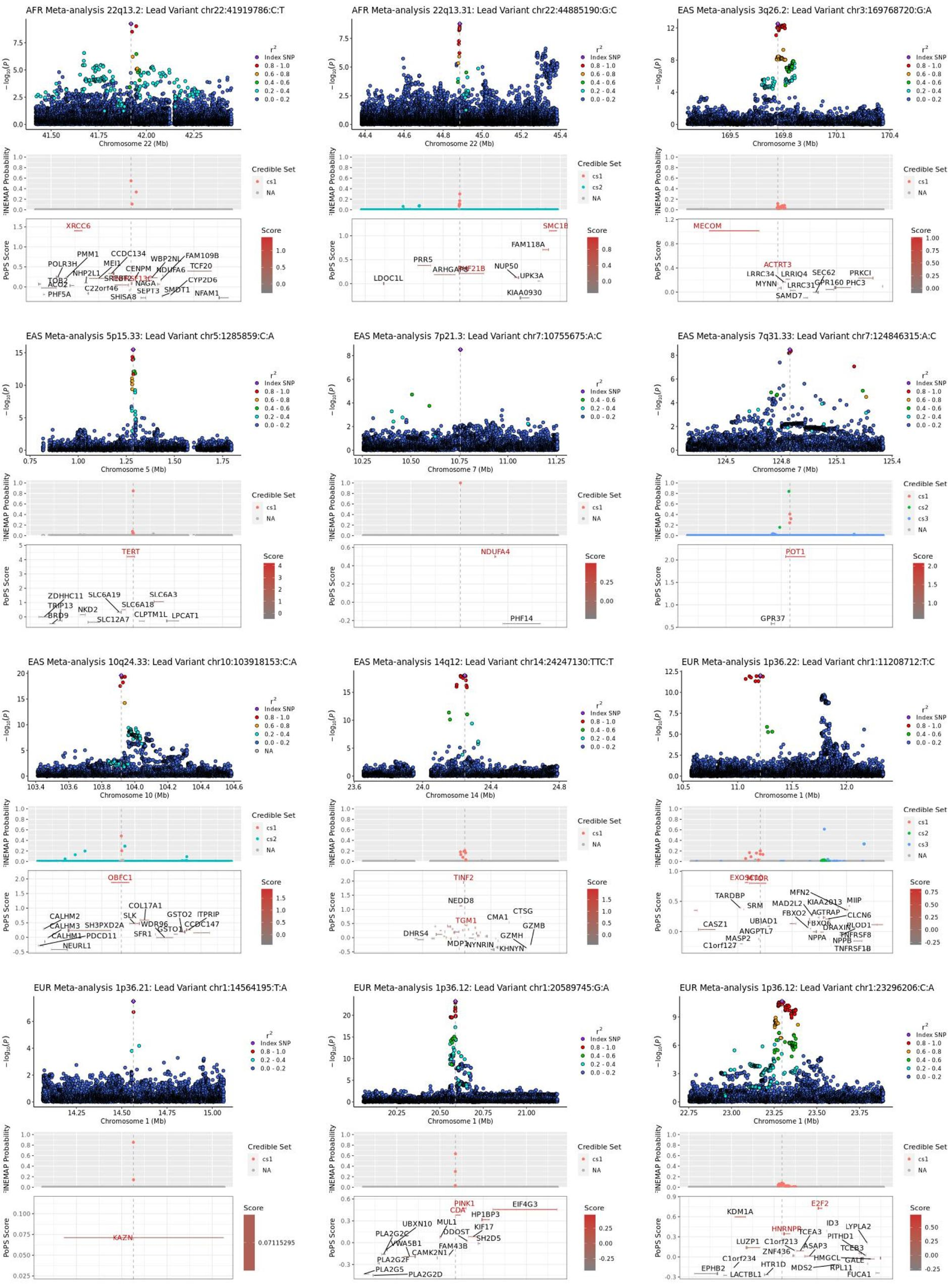

AFR Meta-analysis 22q13.2: Lead Variant chr22:41919786:C:T
Chromosome 22 (Mb)
XRCC6
POLR3H
PMM1
CCDC134
WBP2NL
FAM109B
TCF20
NHP2L1
MEI1
CENPM
NDUFA6
CYP2D6
TOB2
ACO2
C22orf46
NAGA
SMDT1
PHF5A
SEPT3
SHISA8
NFAM1
AFR Meta-analysis 22q13.31: Lead Variant chr22:44885190:G:C
Chromosome 22 (Mb)
SMC1B
FAM118A
PRR5
ARHGAP8
PHF21B
NUP50
UPK3A
LDOC1L
KIAA0930
EAS Meta-analysis 3q26.2: Lead Variant chr3:169768720:G:A
Chromosome 3 (Mb)
MECOM
ACTRT3
LRRC34
LRRIQ4
SEC62
PRKCI
MYNN
LRRC31
GPR160
PHC3
SAMD7
EAS Meta-analysis 5p15.33: Lead Variant chr5:1285859:C:A
Chromosome 5 (Mb)
TERT
ZDHHC11
SLC6A19
SLC6A3
TRIP13
SLC6A18
BRD9
NKD2
CLPTM1L
LPCAT1
SLC12A7
EAS Meta-analysis 7p21.3: Lead Variant chr7:10755675:A:C
Chromosome 7 (Mb)
NDUFA4
PHF14
EAS Meta-analysis 7q31.33: Lead Variant chr7:124846315:A:C
Chromosome 7 (Mb)
POT1
GPR37
EAS Meta-analysis 10q24.33: Lead Variant chr10:103918153:C:A
Chromosome 10 (Mb)
OBFC1
COL17A1
CALHM2
SLK
GSTO2
ITPRIP
CALHM3
SH3PXD2A
WDR96
GSTO1
CCDC147
CALHM1
PDCD11
SFR1
NEURL1
EAS Meta-analysis 14q12: Lead Variant chr14:24247130:TTC:T
Chromosome 14 (Mb)
TINF2
NEDD8
CTSG
CMA1
TGM1
GZMB
DHRS4
GZMH
MDP1
NYNRIN
KHNYN
EUR Meta-analysis 1p36.22: Lead Variant chr1:11208712:T:C
Chromosome 1 (Mb)
TARDBP
MFN2
KIAA2013
MIIP
SRM
MAD2L2
AGTRAP
CLCN6
FBXO2
FBXO6
UBIAD1
PLOD1
CASZ1
DRAXIN
TNFRSF8
ANGPTL7
NPPA
NPPB
MASP2
C1orf127
TNFRSF1B
EUR Meta-analysis 1p36.21: Lead Variant chr1:14564195:T:A
Chromosome 1 (Mb)
KAZN
EUR Meta-analysis 1p36.12: Lead Variant chr1:20589745:G:A
Chromosome 1 (Mb)
PINK1
CDA
EIF4G3
HP1BP3
UBXN10
MUL1
KIF17
PLA2G2C
DDOST
SH2D5
VWA5B1
FAM43B
CAMK2N1
PLA2G2F
PLA2G5
PLA2G2D
EUR Meta-analysis 1p36.12: Lead Variant chr1:23296206:C:A
Chromosome 1 (Mb)
E2F2
KDM1A
ID3
LYPLA2
HNRNPR
TCEA3
PITHD1
LUZP1
C1orf213
ASAP3
TCEB3
ZNF436
HMGCL
GALE
C1orf234
HTR1D
MDS2
RPL11
EPHB2
LACTBL1
FUCA1
$-\log_{10}(p)$
FINEMAP Probability
PoPS Score
$r^2$
Index SNP
0.8 - 1.0
0.6 - 0.8
0.4 - 0.6
0.2 - 0.4
0.0 - 0.2
NA
Credible Set
cs1
cs2
cs3
Score

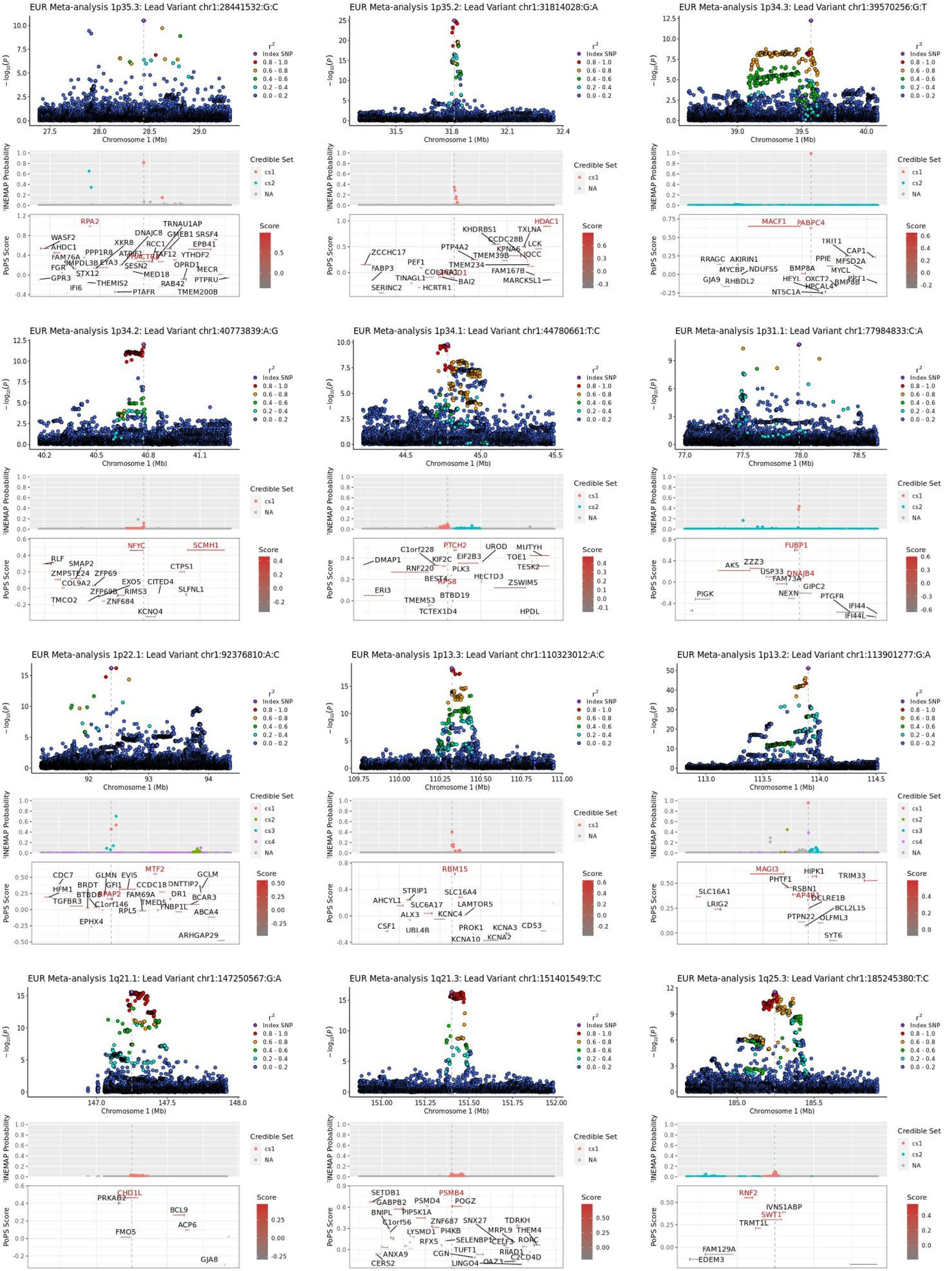
EUR Meta-analysis 1p35.3: Lead Variant chr1:28441532:G:C
EUR Meta-analysis 1p35.2: Lead Variant chr1:31814028:G:A
EUR Meta-analysis 1p34.3: Lead Variant chr1:39570256:G:T
EUR Meta-analysis 1p34.2: Lead Variant chr1:40773839:A:G
EUR Meta-analysis 1p34.1: Lead Variant chr1:44780661:T:C
EUR Meta-analysis 1p31.1: Lead Variant chr1:77984833:C:A
EUR Meta-analysis 1p22.1: Lead Variant chr1:92376810:A:C
EUR Meta-analysis 1p13.3: Lead Variant chr1:110323012:A:C
EUR Meta-analysis 1p13.2: Lead Variant chr1:113901277:G:A
EUR Meta-analysis 1q21.1: Lead Variant chr1:147250567:G:A
EUR Meta-analysis 1q21.3: Lead Variant chr1:151401549:T:C
EUR Meta-analysis 1q25.3: Lead Variant chr1:185245380:T:C
Chromosome 1 (Mb)
Index SNP
0.8 - 1.0
0.6 - 0.8
0.4 - 0.6
0.2 - 0.4
0.0 - 0.2
Credible Set
FINEMAP Probability
PoPS Score
Score

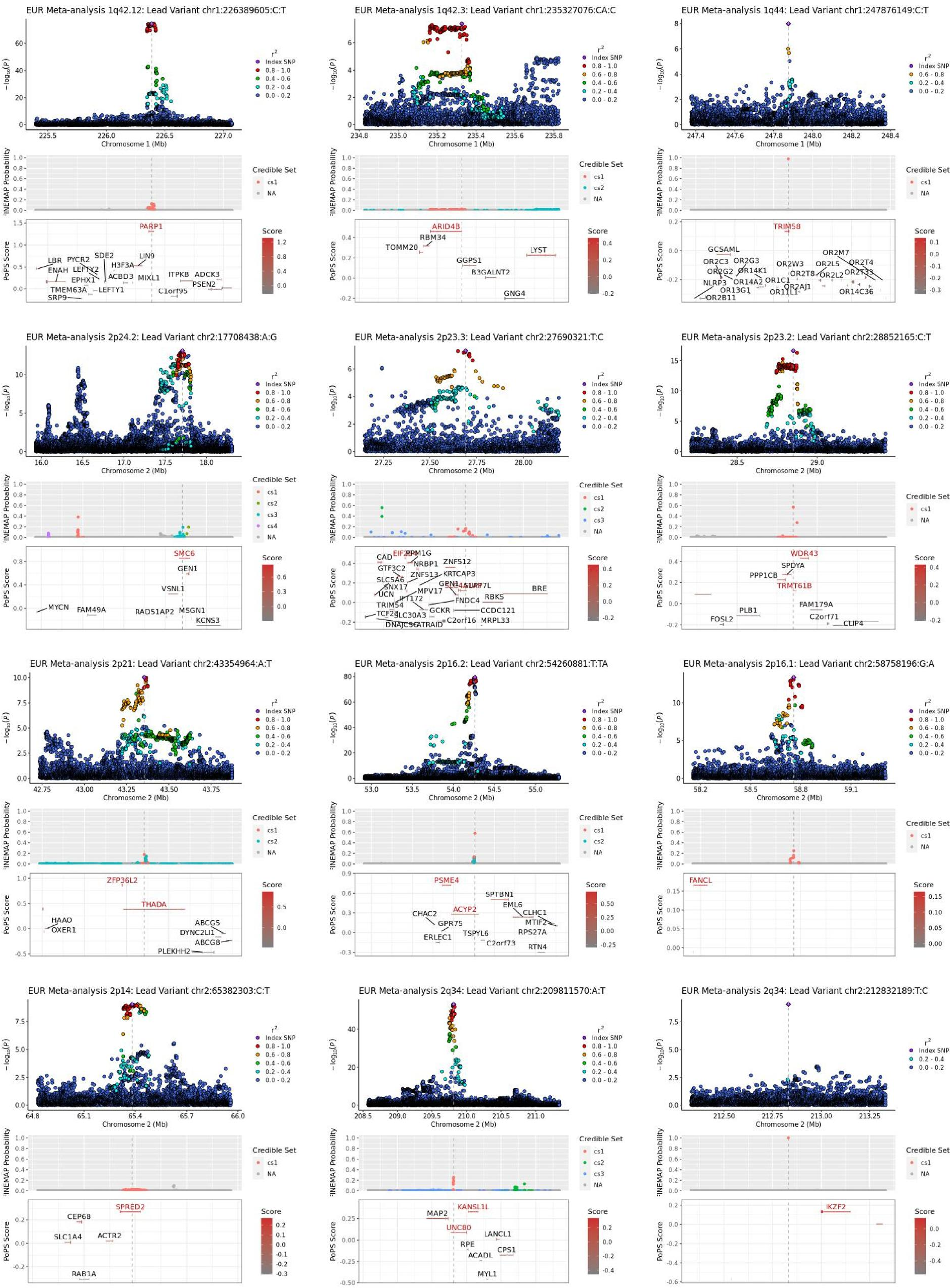
EUR Meta-analysis 1q42.12: Lead Variant chr1:226389605:C:T
EUR Meta-analysis 1q42.3: Lead Variant chr1:235327076:CA:C
EUR Meta-analysis 1q44: Lead Variant chr1:247876149:C:T
EUR Meta-analysis 2p24.2: Lead Variant chr2:17708438:A:G
EUR Meta-analysis 2p23.3: Lead Variant chr2:27690321:T:C
EUR Meta-analysis 2p23.2: Lead Variant chr2:28852165:C:T
EUR Meta-analysis 2p21: Lead Variant chr2:43354964:A:T
EUR Meta-analysis 2p16.2: Lead Variant chr2:54260881:T:TA
EUR Meta-analysis 2p16.1: Lead Variant chr2:58758196:G:A
EUR Meta-analysis 2p14: Lead Variant chr2:65382303:C:T
EUR Meta-analysis 2q34: Lead Variant chr2:209811570:A:T
EUR Meta-analysis 2q34: Lead Variant chr2:212832189:T:C
Index SNP
Credible Set
PoPS Score
Score
PARP1
ARID4B
TRIM58
SMC6
WDR43
TRMT61B
ZFP36L2
THADA
PSME4
ACYP2
FANCL
SPRED2
KANSL1L
UNC80
IKZF2

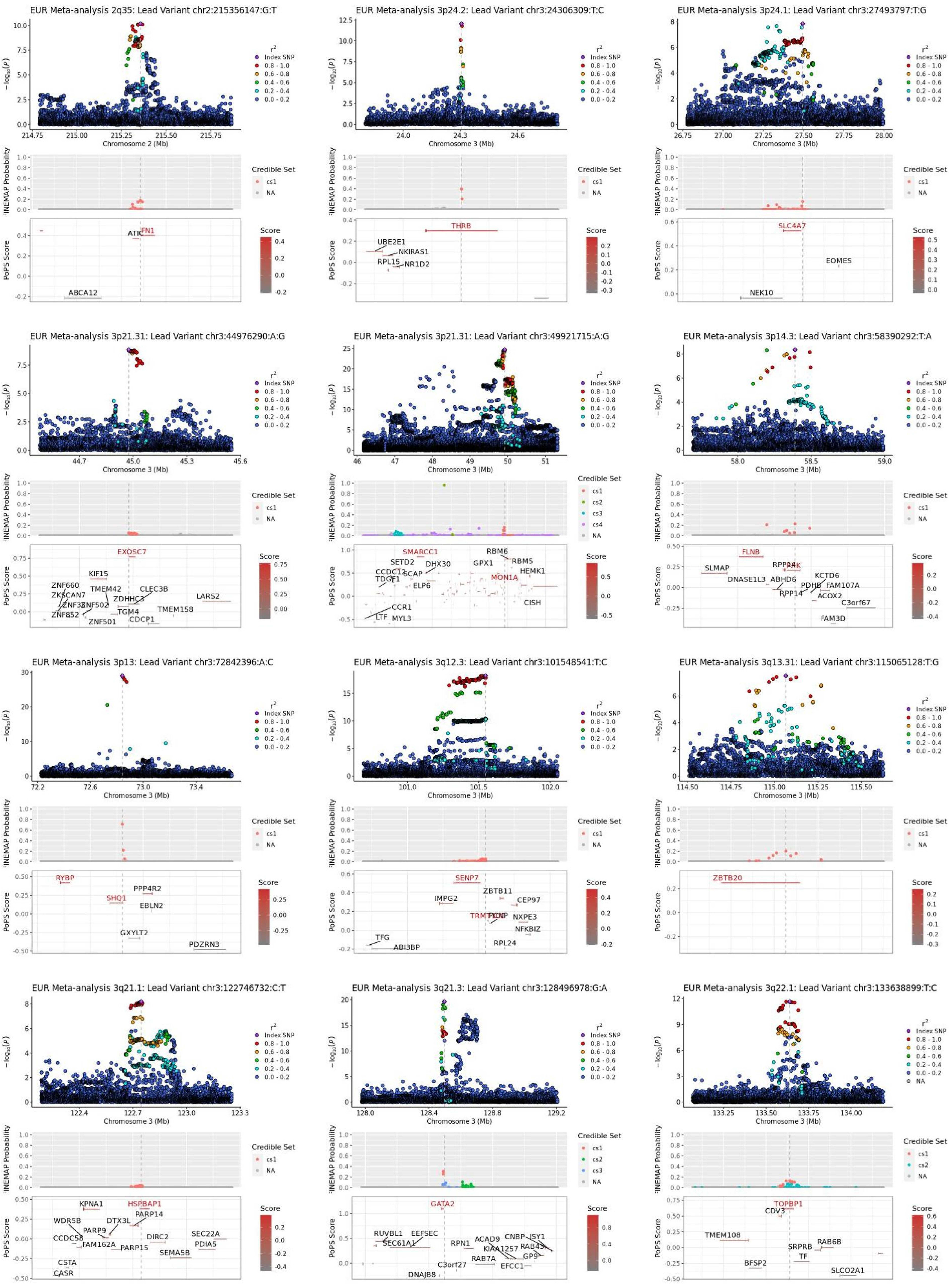
EUR Meta-analysis 2q35: Lead Variant chr2:215356147:G:T
EUR Meta-analysis 3p24.2: Lead Variant chr3:24306309:T:C
EUR Meta-analysis 3p24.1: Lead Variant chr3:27493797:T:G
EUR Meta-analysis 3p21.31: Lead Variant chr3:44976290:A:G
EUR Meta-analysis 3p21.31: Lead Variant chr3:49921715:A:G
EUR Meta-analysis 3p14.3: Lead Variant chr3:58390292:T:A
EUR Meta-analysis 3p13: Lead Variant chr3:72842396:A:C
EUR Meta-analysis 3q12.3: Lead Variant chr3:101548541:T:C
EUR Meta-analysis 3q13.31: Lead Variant chr3:115065128:T:G
EUR Meta-analysis 3q21.1: Lead Variant chr3:122746732:C:T
EUR Meta-analysis 3q21.3: Lead Variant chr3:128496978:G:A
EUR Meta-analysis 3q22.1: Lead Variant chr3:133638899:T:C
−log10(P)
Chromosome 2 (Mb)
Chromosome 3 (Mb)
r2
Index SNP
0.8 - 1.0
0.6 - 0.8
0.4 - 0.6
0.2 - 0.4
0.0 - 0.2
NA
FINEMAP Probability
Credible Set
cs1
cs2
cs3
cs4
NA
PoPS Score
Score
ATIC
FN1
ABCA12
THRB
UBE2E1
NKIRAS1
RPL15
NR1D2
SLC4A7
EOMES
NEK10
EXOSC7
KIF15
ZNF660
ZKSCAN7
TMEM42
CLEC3B
ZNF502
ZDHHC3
LARS2
TGM4
TMEM158
ZNF501
CDCP1
SMARCC1
RBM6
RBM5
SETD2
DHX30
GPX1
HEMK1
SCAP
MON1A
ELP6
CISH
CCR1
LTF
MYL3
FLNB
SLMAP
DNASE1L3
ABHD6
KCTD6
PDHB
FAM107A
RPP14
ACOX2
C3orf67
FAM3D
RYBP
PPP4R2
SHQ1
EBLN2
GXYLT2
PDZRN3
SENP7
ZBTB11
IMPG2
CEP97
NXPE3
NFKBIZ
RPL24
TFG
ABI3BP
ZBTB20
KPNA1
HSPBAP1
WDR5B
DTX3L
PARP14
PARP9
CCDC58
FAM162A
DIRC2
SEC22A
PARP15
PDIA5
SEMA5B
CSTA
CASR
GATA2
RUVBL1
EEFSEC
SEC61A1
RPN1
ACAD9
CNBP
ISY1
KIAA1257
RAB43
RAB7A
GP9
C3orf27
EFCC1
DNAJB8
TOPBP1
CDV3
TMEM108
RAB6B
SRPRB
TF
BFSP2
SLCO2A1

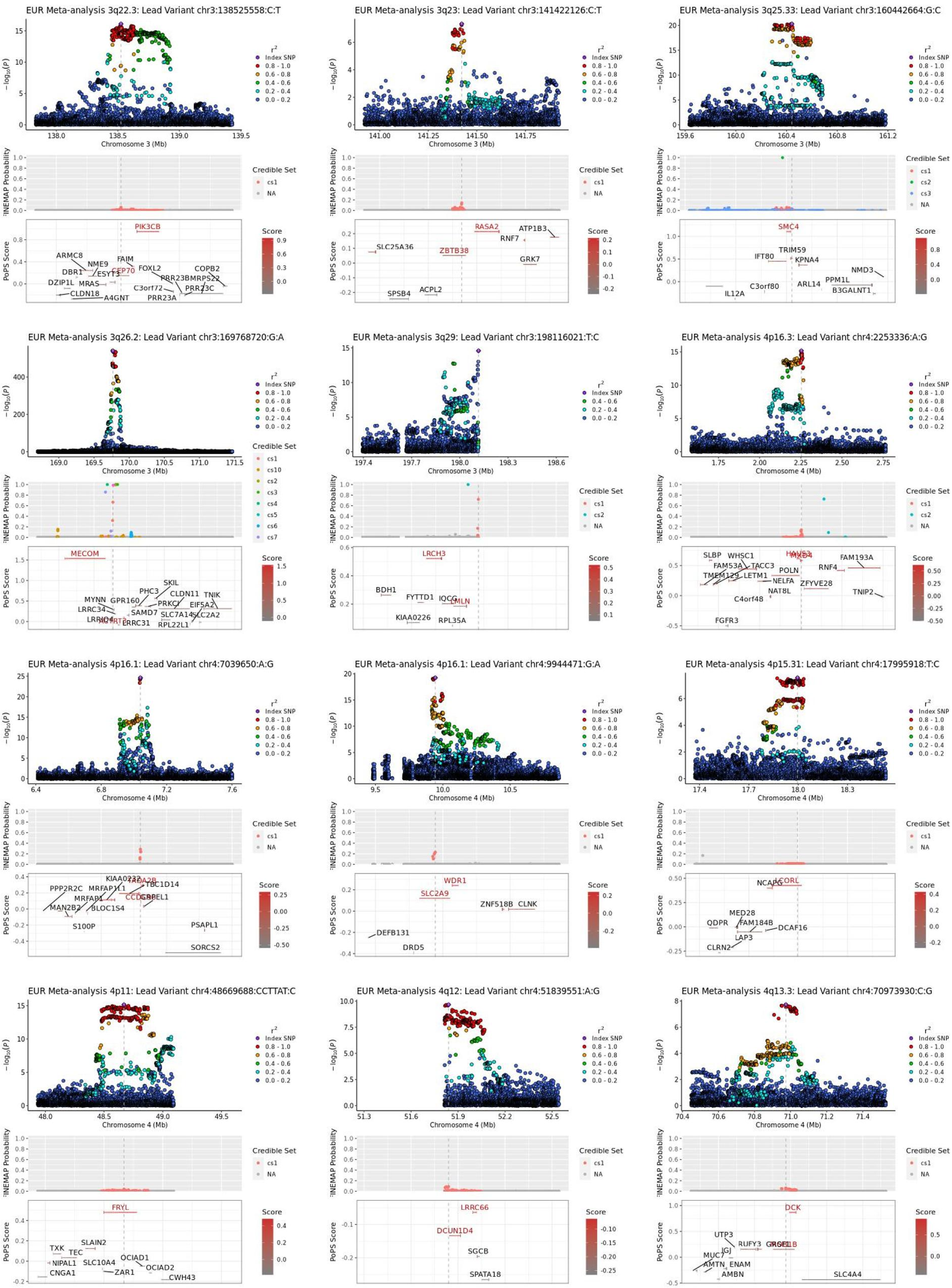
EUR Meta-analysis 3q22.3: Lead Variant chr3:138525558:C:T
EUR Meta-analysis 3q23: Lead Variant chr3:141422126:C:T
EUR Meta-analysis 3q25.33: Lead Variant chr3:160442664:G:C
EUR Meta-analysis 3q26.2: Lead Variant chr3:169768720:G:A
EUR Meta-analysis 3q29: Lead Variant chr3:198116021:T:C
EUR Meta-analysis 4p16.3: Lead Variant chr4:2253336:A:G
EUR Meta-analysis 4p16.1: Lead Variant chr4:7039650:A:G
EUR Meta-analysis 4p16.1: Lead Variant chr4:9944471:G:A
EUR Meta-analysis 4p15.31: Lead Variant chr4:17995918:T:C
EUR Meta-analysis 4p11: Lead Variant chr4:48669688:CCTTAT:C
EUR Meta-analysis 4q12: Lead Variant chr4:51839551:A:G
EUR Meta-analysis 4q13.3: Lead Variant chr4:70973930:C:G
Chromosome 3 (Mb)
Chromosome 4 (Mb)
FINEMAP Probability
PoPS Score
Credible Set
Score
Index SNP

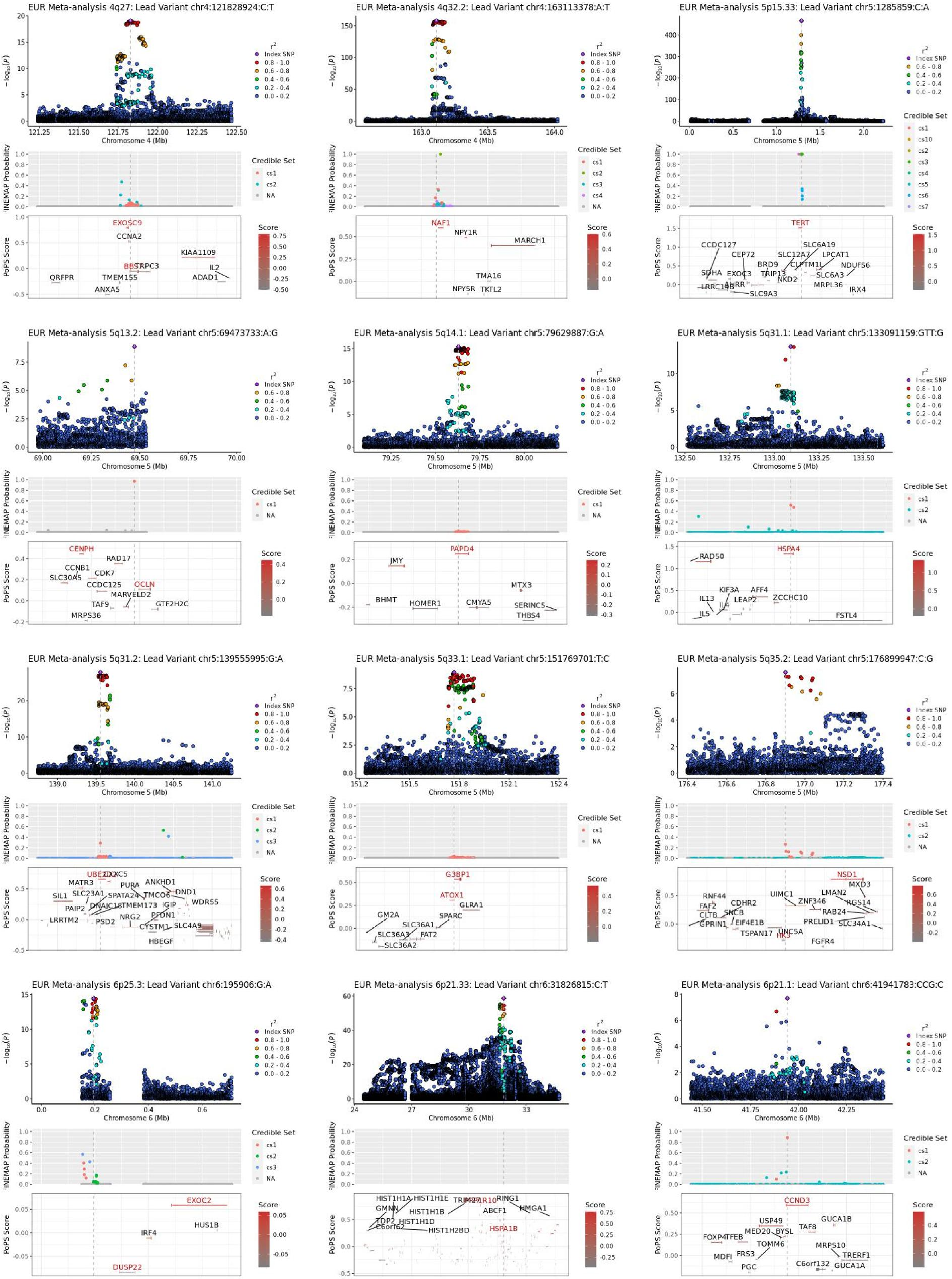
EUR Meta-analysis 4q27: Lead Variant chr4:121828924:C:T
Chromosome 4 (Mb)
FINEMAP Probability
PoPS Score
EXOSC9
CCNA2
KIAA1109
BBS7
TRPC3
IL2
QRFPR
TMEM155
ADAD1
ANXA5
EUR Meta-analysis 4q32.2: Lead Variant chr4:163113378:A:T
NAF1
NPY1R
MARCH1
TMA16
NPY5R
TKTL2
EUR Meta-analysis 5p15.33: Lead Variant chr5:1285859:C:A
Chromosome 5 (Mb)
TERT
CCDC127
SLC6A19
CEP72
SLC12A7
LPCAT1
BRD9
CLPTM1L
NDUFS6
SDHA
EXOC3
TRIP13
SLC6A3
AHRR
NKD2
MRPL36
LRRC14B
IRX4
SLC9A3
EUR Meta-analysis 5q13.2: Lead Variant chr5:69473733:A:G
CENPH
RAD17
CCNB1
CDK7
SLC30A5
CCDC125
OCLN
MARVELD2
TAF9
GTF2H2C
MRPS36
EUR Meta-analysis 5q14.1: Lead Variant chr5:79629887:G:A
PAPD4
JMY
MTX3
BHMT
HOMER1
CMYA5
SERINC5
THBS4
EUR Meta-analysis 5q31.1: Lead Variant chr5:133091159:GTT:G
RAD50
HSPA4
KIF3A
AFF4
IL13
LEAP2
ZCCHC10
IL4
IL5
FSTL4
EUR Meta-analysis 5q31.2: Lead Variant chr5:139555995:G:A
UBE2D2
CXXC5
MATR3
PURA
ANKHD1
SIL1
SLC23A1
SPATA24
TMCO6
DND1
PAIP2
DNAJC18
TMEM173
WDR55
IGIP
LRRTM2
PFDN1
PSD2
NRG2
CYSTM1
SLC4A9
HBEGF
EUR Meta-analysis 5q33.1: Lead Variant chr5:151769701:T:C
G3BP1
ATOX1
GLRA1
GM2A
SPARC
SLC36A1
SLC36A3
FAT2
SLC36A2
EUR Meta-analysis 5q35.2: Lead Variant chr5:176899947:C:G
NSD1
MXD3
RNF44
LMAN2
UIMC1
FAF2
CDHR2
ZNF346
RGS14
CLTB
SNCB
RAB24
EIF4E1B
PRELID1
GPRIN1
SLC34A1
TSPAN17
UNC5A
HK3
FGFR4
EUR Meta-analysis 6p25.3: Lead Variant chr6:195906:G:A
Chromosome 6 (Mb)
EXOC2
HUS1B
IRF4
DUSP22
EUR Meta-analysis 6p21.33: Lead Variant chr6:31826815:C:T
HIST1H1A
HIST1H1E
RING1
GMNN
HIST1H1B
TRIM27
HMGA1
ABCF1
TDP2
HIST1H1D
HSPA1B
C6orf62
HIST1H2BD
EUR Meta-analysis 6p21.1: Lead Variant chr6:41941783:CCG:C
CCND3
USP49
GUCA1B
TAF8
FOXP4
TFEB
MED20
BYSL
TOMM6
MRPS10
MDFI
FRS3
TRERF1
PGC
C6orf132
GUCA1A
$r^2$
Index SNP
0.8 - 1.0
0.6 - 0.8
0.4 - 0.6
0.2 - 0.4
0.0 - 0.2
Credible Set
cs1
cs2
cs3
cs4
cs5
cs6
cs7
cs10
NA
Score

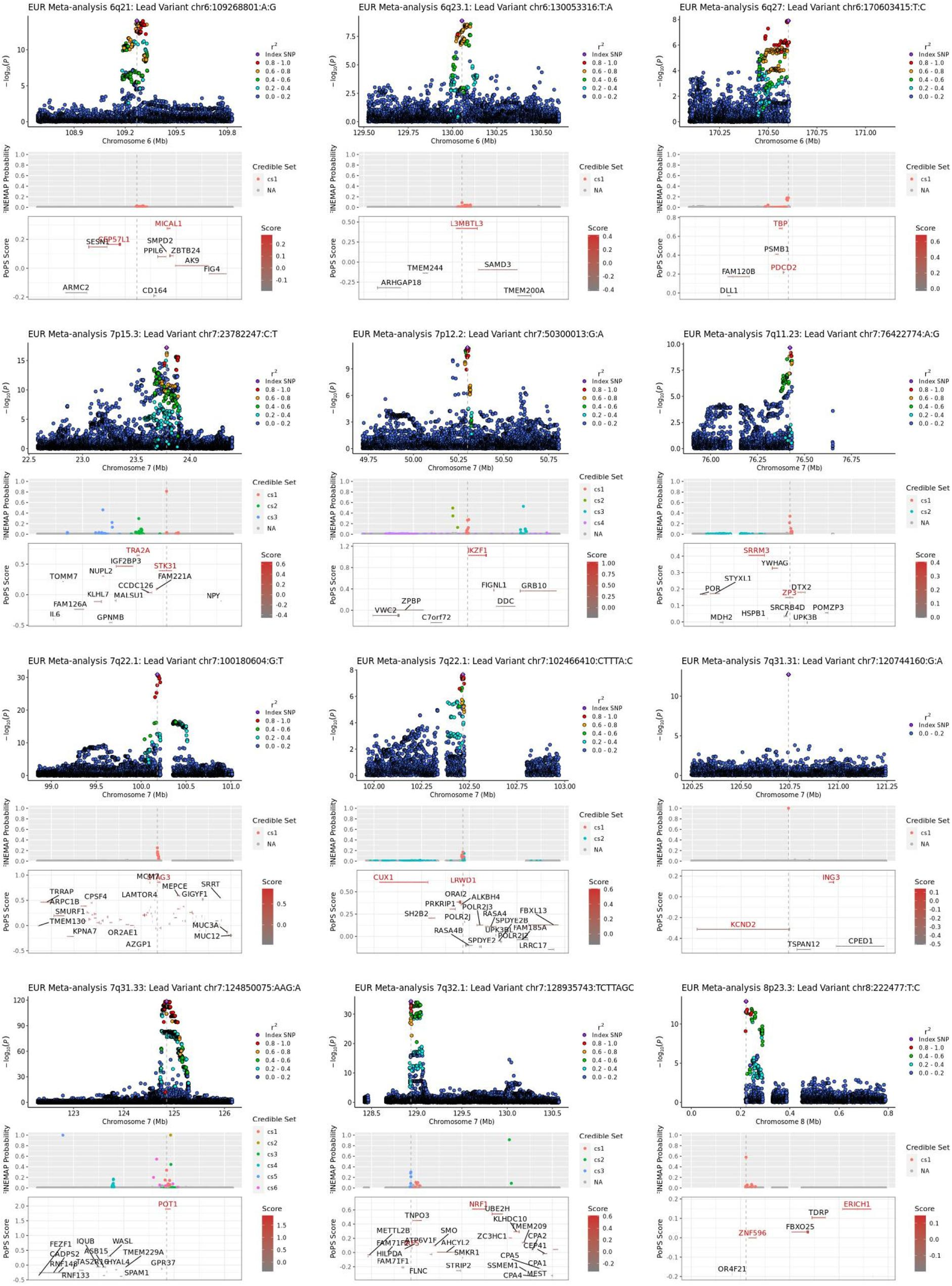

EUR Meta-analysis 6q21: Lead Variant chr6:109268801:A:G
EUR Meta-analysis 6q23.1: Lead Variant chr6:130053316:T:A
EUR Meta-analysis 6q27: Lead Variant chr6:170603415:T:C
EUR Meta-analysis 7p15.3: Lead Variant chr7:23782247:C:T
EUR Meta-analysis 7p12.2: Lead Variant chr7:50300013:G:A
EUR Meta-analysis 7q11.23: Lead Variant chr7:76422774:A:G
EUR Meta-analysis 7q22.1: Lead Variant chr7:100180604:G:T
EUR Meta-analysis 7q22.1: Lead Variant chr7:102466410:CTTTA:C
EUR Meta-analysis 7q31.31: Lead Variant chr7:120744160:G:A
EUR Meta-analysis 7q31.33: Lead Variant chr7:124850075:AAG:A
EUR Meta-analysis 7q32.1: Lead Variant chr7:128935743:TCTTAGC
EUR Meta-analysis 8p23.3: Lead Variant chr8:222477:T:C
$-\log_{10}(P)$
Chromosome 6 (Mb)
Chromosome 7 (Mb)
Chromosome 8 (Mb)
$r^2$
Index SNP
0.8 - 1.0
0.6 - 0.8
0.4 - 0.6
0.2 - 0.4
0.0 - 0.2
FINEMAP Probability
Credible Set
cs1
cs2
cs3
cs4
cs5
cs6
NA
PoPS Score
Score
MICAL1
SESN1
CEP57L1
SMPD2
PPIL6
ZBTB24
AK9
FIG4
ARMC2
CD164
L3MBTL3
TMEM244
SAMD3
ARHGAP18
TMEM200A
TBP
PSMB1
PDCD2
FAM120B
DLL1
TRA2A
IGF2BP3
STK31
NUPL2
TOMM7
CCDC126
FAM221A
KLHL7
MALSU1
NPY
FAM126A
IL6
GPNMB
IKZF1
FIGNL1
GRB10
ZPBP
VWC2
DDC
C7orf72
SRRM3
YWHAG
STYXL1
POR
DTX2
ZP3
SRCRB4D
POMZP3
MDH2
HSPB1
UPK3B
MCM7
STAG3
TRRAP
MEPCE
SRRT
ARPC1B
CPSF4
LAMTOR4
GIGYF1
SMURF1
TMEM130
MUC3A
KPNA7
OR2AE1
MUC12
AZGP1
CUX1
LRWD1
ORAI2
ALKBH4
PRKRIP1
POLR2J3
SH2B2
POLR2J
RASA4
FBXL13
SPDYE2B
FAM185A
RASA4B
UPK3BL
POLR2J2
SPDYE2
LRRC17
ING3
KCND2
TSPAN12
CPED1
POT1
FEZF1
IQUB
WASL
CADPS2
ASB15
RNF148
TAS2R16
HYAL4
TMEM229A
GPR37
RNF133
SPAM1
NRF1
UBE2H
TNPO3
KLHDC10
TMEM209
METTL2B
SMO
ZC3HC1
FAM71F2
ATP6V1F
AHCYL2
CEP41
CPA2
HILPDA
SMKR1
CPA5
CPA1
FAM71F1
STRIP2
SSMEM1
FLNC
CPA4
MEST
ERICH1
TDRP
FBXO25
ZNF596
OR4F21

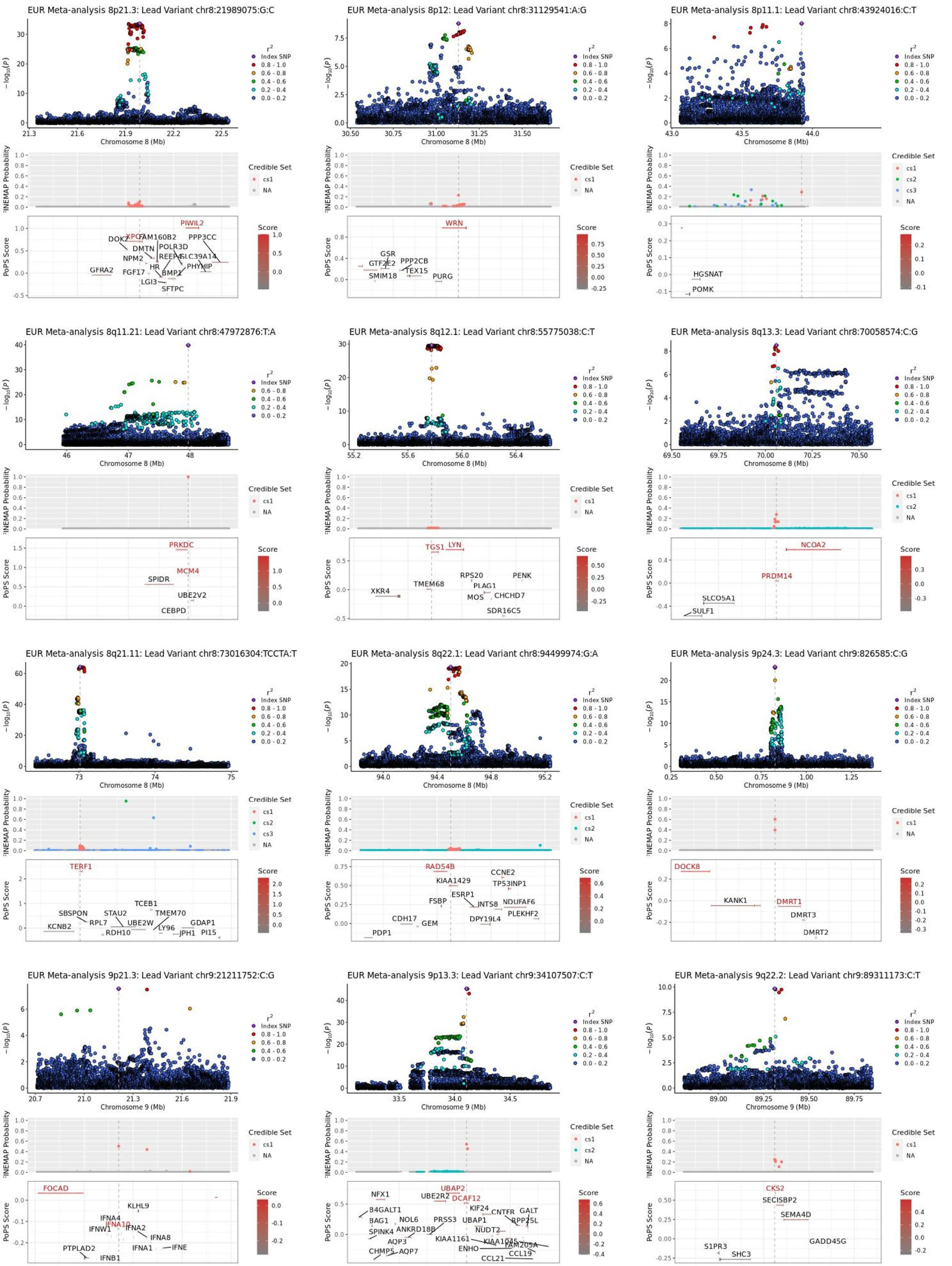
EUR Meta-analysis 8p21.3: Lead Variant chr8:21989075:G:C
EUR Meta-analysis 8p12: Lead Variant chr8:31129541:A:G
EUR Meta-analysis 8p11.1: Lead Variant chr8:43924016:C:T
EUR Meta-analysis 8q11.21: Lead Variant chr8:47972876:T:A
EUR Meta-analysis 8q12.1: Lead Variant chr8:55775038:C:T
EUR Meta-analysis 8q13.3: Lead Variant chr8:70058574:C:G
EUR Meta-analysis 8q21.11: Lead Variant chr8:73016304:TCCTA:T
EUR Meta-analysis 8q22.1: Lead Variant chr8:94499974:G:A
EUR Meta-analysis 9p24.3: Lead Variant chr9:826585:C:G
EUR Meta-analysis 9p21.3: Lead Variant chr9:21211752:C:G
EUR Meta-analysis 9p13.3: Lead Variant chr9:34107507:C:T
EUR Meta-analysis 9q22.2: Lead Variant chr9:89311173:C:T
Chromosome 8 (Mb)
Chromosome 9 (Mb)
$-\log_{10}(P)$
FINEMAP Probability
PoPS Score
$r^2$
Index SNP
0.8 - 1.0
0.6 - 0.8
0.4 - 0.6
0.2 - 0.4
0.0 - 0.2
Credible Set
cs1
cs2
cs3
NA
Score
PIWIL2
XPO7
DOK2
FAM160B2
PPP3CC
DMTN
POLR3D
NPM2
REEP4
SLC39A14
GFRA2
FGF17
HR
BMP1
PHYHIP
LGI3
SFTPC
WRN
GSR
GTF2E2
PPP2CB
SMIM18
TEX15
PURG
HGSNAT
POMK
PRKDC
MCM4
SPIDR
UBE2V2
CEBPD
TGS1
LYN
RPS20
PENK
TMEM68
PLAG1
XKR4
MOS
CHCHD7
SDR16C5
NCOA2
PRDM14
SLCO5A1
SULF1
TERF1
TCEB1
SBSPON
STAU2
TMEM70
RPL7
UBE2W
GDAP1
KCNB2
LY96
RDH10
JPH1
PI15
RAD54B
CCNE2
KIAA1429
TP53INP1
ESRP1
FSBP
INTS8
NDUFAF6
PLEKHF2
CDH17
DPY19L4
GEM
PDP1
DOCK8
KANK1
DMRT1
DMRT3
DMRT2
FOCAD
KLHL9
IFNA4
IFNW1
IFNA10
IFNA2
IFNA8
IFNE
IFNA1
PTPLAD2
IFNB1
NFX1
UBAP2
UBE2R2
DCAF12
B4GALT1
KIF24
CNTFR
GALT
BAG1
NOL6
PRSS3
UBAP1
RPP25L
SPINK4
ANKRD18B
NUDT2
AQP3
KIAA1161
KIAA1045
FAM205A
CHMP5
AQP7
ENHO
CCL19
CCL21
CKS2
SECISBP2
SEMA4D
GADD45G
S1PR3
SHC3

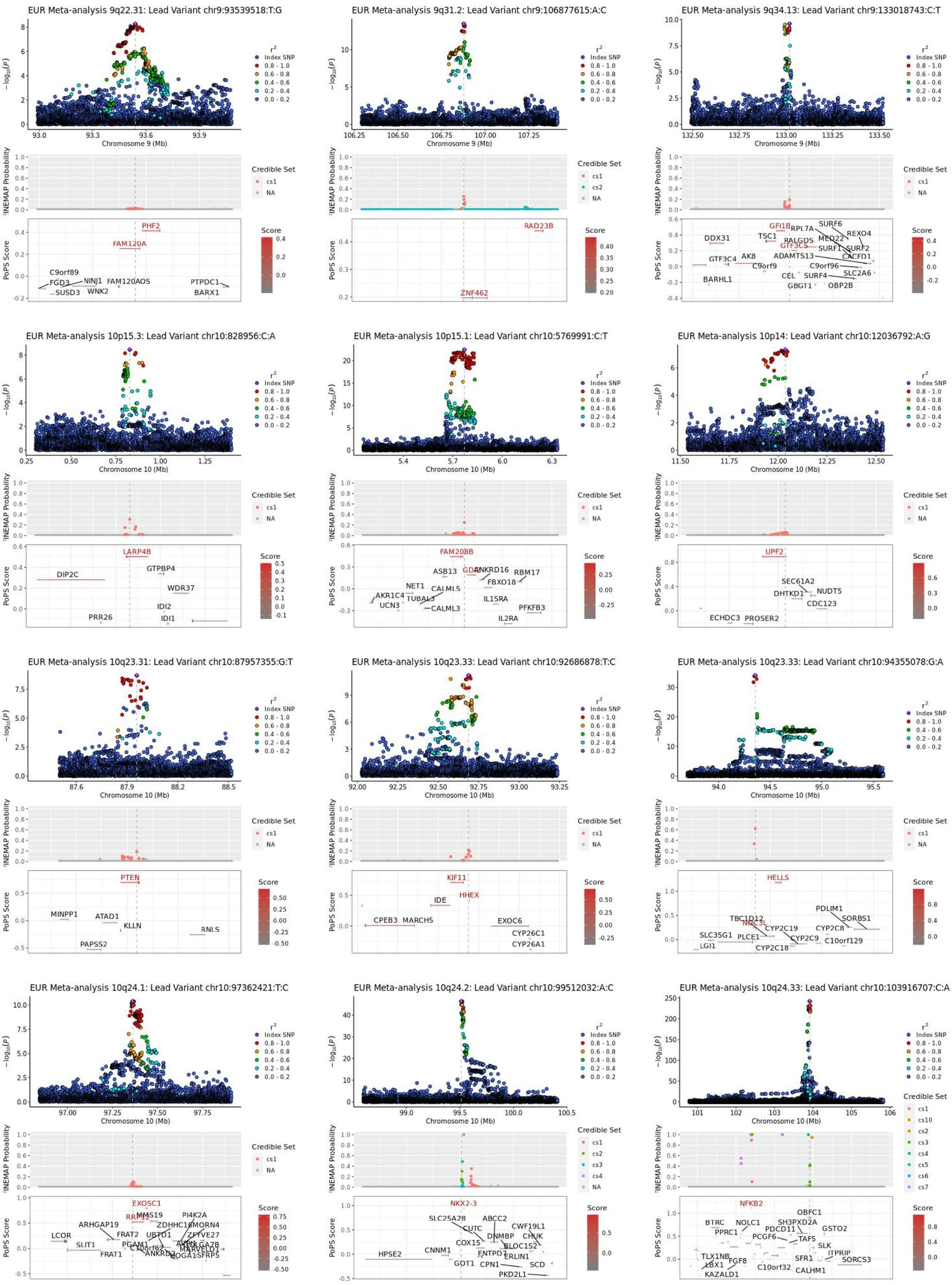
EUR Meta-analysis 9q22.31: Lead Variant chr9:93539518:T:G
EUR Meta-analysis 9q31.2: Lead Variant chr9:106877615:A:C
EUR Meta-analysis 9q34.13: Lead Variant chr9:133018743:C:T
Chromosome 9 (Mb)
PHF2
FAM120A
C9orf89
FGD3
NINJ1
SUSD3
WNK2
FAM120AOS
PTPDC1
BARX1
RAD23B
ZNF462
GFI1B
GTF3C5
DDX31
TSC1
RPL7A
SURF6
MED22
REXO4
RALGDS
SURF1
SURF2
GTF3C4
AK8
ADAMTS13
CACFD1
C9orf9
C9orf96
CEL
SURF4
SLC2A6
BARHL1
GBGT1
OBP2B
EUR Meta-analysis 10p15.3: Lead Variant chr10:828956:C:A
EUR Meta-analysis 10p15.1: Lead Variant chr10:5769991:C:T
EUR Meta-analysis 10p14: Lead Variant chr10:12036792:A:G
Chromosome 10 (Mb)
LARP4B
DIP2C
GTPBP4
WDR37
IDI2
IDI1
PRR26
FAM208B
ASB13
GDI2
ANKRD16
RBM17
FBXO18
NET1
CALML5
AKR1C4
TUBAL3
UCN3
CALML3
IL15RA
PFKFB3
IL2RA
UPF2
SEC61A2
DHTKD1
NUDT5
CDC123
ECHDC3
PROSER2
EUR Meta-analysis 10q23.31: Lead Variant chr10:87957355:G:T
EUR Meta-analysis 10q23.33: Lead Variant chr10:92686878:T:C
EUR Meta-analysis 10q23.33: Lead Variant chr10:94355078:G:A
PTEN
MINPP1
ATAD1
KLLN
RNLS
PAPSS2
KIF11
HHEX
IDE
CPEB3
MARCH5
EXOC6
CYP26C1
CYP26A1
HELLS
PDLIM1
SORBS1
TBC1D12
NOC3L
CYP2C19
CYP2C8
SLC35G1
PLCE1
CYP2C9
C10orf129
LGI1
CYP2C18
EUR Meta-analysis 10q24.1: Lead Variant chr10:97362421:T:C
EUR Meta-analysis 10q24.2: Lead Variant chr10:99512032:A:C
EUR Meta-analysis 10q24.33: Lead Variant chr10:103916707:C:A
EXOSC1
PI4K2A
ARHGAP19
MMS19
ZDHHC16
MORN4
LCOR
FRAT2
UBTD1
ZFYVE27
SLIT1
PGAM1
FRAT1
ANKRD2
SFRP5
NKX2-3
SLC25A28
ABCC2
CUTC
CWF19L1
DNMBP
CHUK
COX15
BLOC1S2
ENTPD7
ERLIN1
HPSE2
CNNM1
GOT1
CPN1
SCD
PKD2L1
NFKB2
OBFC1
BTRC
NOLC1
SH3PXD2A
PPRC1
PDCD11
GSTO2
PCGF6
TAF5
SLK
TLX1NB
ITPRIP
LBX1
FGF8
SFR1
SORCS3
KAZALD1
C10orf32
CALHM1
−log10(p)
FINEMAP Probability
PoPS Score
Credible Set
Score
Index SNP

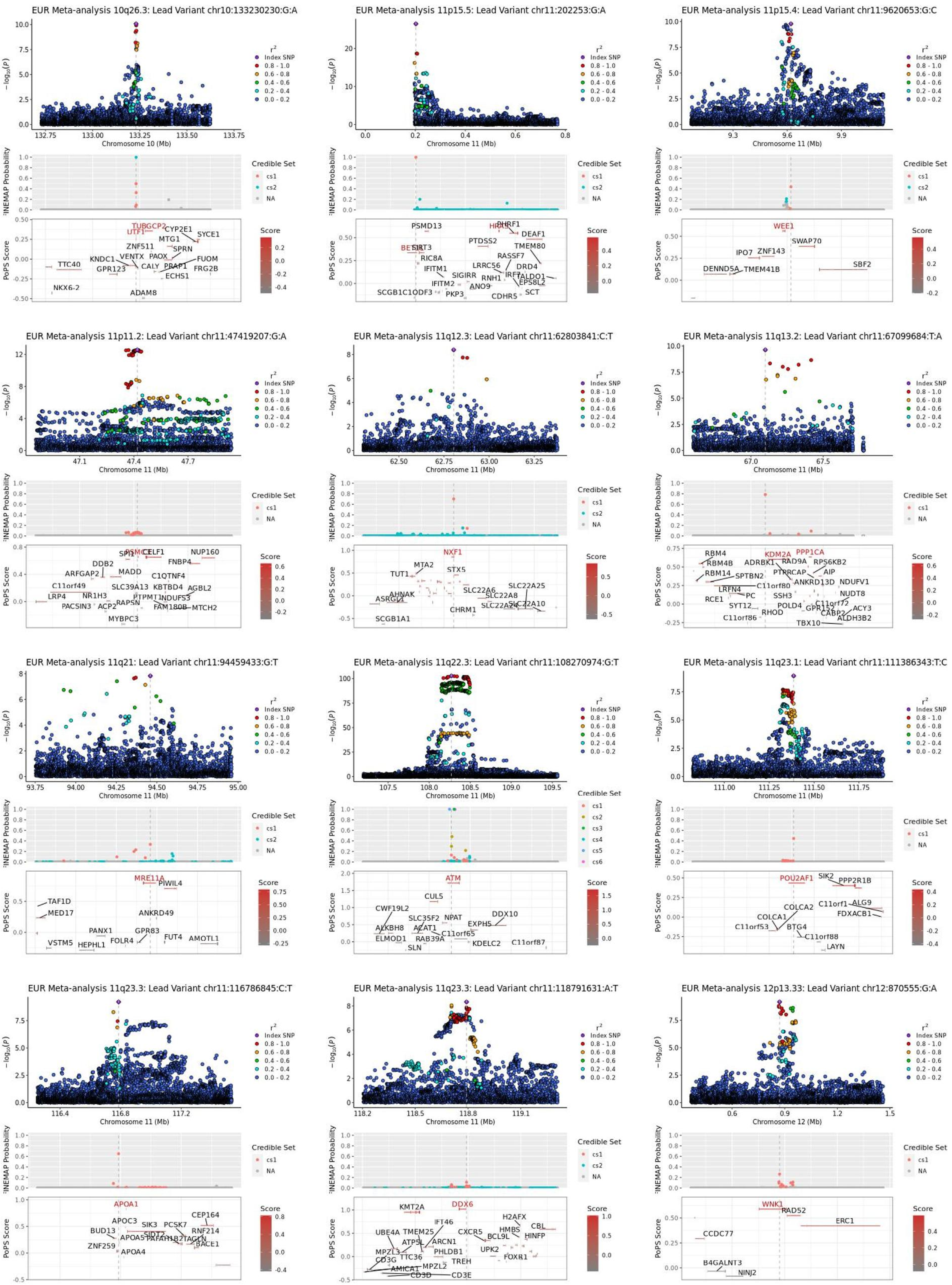
EUR Meta-analysis 10q26.3: Lead Variant chr10:133230230:G:A
Chromosome 10 (Mb)
TUBGCP2
UTF1
CYP2E1
SYCE1
MTG1
ZNF511
SPRN
TTC40
KNDC1
VENTX
PAOX
FUOM
GPR123
CALY
PRAP1
FRG2B
ECHS1
NKX6-2
ADAM8
EUR Meta-analysis 11p15.5: Lead Variant chr11:202253:G:A
Chromosome 11 (Mb)
PSMD13
PHRF1
DEAF1
BET1L
SIRT3
RIC8A
PTDSS2
TMEM80
RASSF7
IFITM1
LRRC56
DRD4
SIGIRR
RNH1
IRF7
TALDO1
IFITM2
ANO9
EPS8L2
SCGB1C1
ODF3
PKP3
SCT
CDHR5
EUR Meta-analysis 11p15.4: Lead Variant chr11:9620653:G:C
Chromosome 11 (Mb)
WEE1
SWAP70
IPO7
ZNF143
DENND5A
TMEM41B
SBF2
EUR Meta-analysis 11p11.2: Lead Variant chr11:47419207:G:A
Chromosome 11 (Mb)
PSMC3
SPI1
CELF1
NUP160
DDB2
FNBP4
ARFGAP2
MADD
C1QTNF4
C11orf49
SLC39A13
KBTBD4
AGBL2
LRP4
NR1H3
PTPMT1
NDUFS3
PACSIN3
ACP2
RAPSN
FAM180B
MTCH2
MYBPC3
EUR Meta-analysis 11q12.3: Lead Variant chr11:62803841:C:T
Chromosome 11 (Mb)
NXF1
MTA2
TUT1
STX5
AHNAK
SLC22A6
SLC22A25
ASRGL1
SLC22A8
SLC22A10
CHRM1
SCGB1A1
EUR Meta-analysis 11q13.2: Lead Variant chr11:67099684:T:A
Chromosome 11 (Mb)
RBM4
KDM2A
PPP1CA
RBM4B
ADRBK1
RAD9A
RPS6KB2
RBM14
SPTBN2
PTPRCAP
AIP
ANKRD13D
NDUFV1
LRFN4
C11orf80
NUDT8
RCE1
PC
SSH3
C11orf72
SYT12
POLD4
GPR152
ACY3
CABP2
C11orf86
RHOD
ALDH3B2
TBX10
EUR Meta-analysis 11q21: Lead Variant chr11:94459433:G:T
Chromosome 11 (Mb)
MRE11A
PIWIL4
TAF1D
MED17
ANKRD49
PANX1
GPR83
FUT4
AMOTL1
VSTM5
HEPHL1
FOLR4
EUR Meta-analysis 11q22.3: Lead Variant chr11:108270974:G:T
Chromosome 11 (Mb)
ATM
CUL5
CWF19L2
SLC35F2
NPAT
DDX10
ALKBH8
ACAT1
EXPH5
ELMOD1
C11orf65
RAB39A
KDELC2
C11orf87
SLN
EUR Meta-analysis 11q23.1: Lead Variant chr11:111386343:T:C
Chromosome 11 (Mb)
POU2AF1
SIK2
PPP2R1B
C11orf1
ALG9
COLCA2
FDXACB1
COLCA1
C11orf53
BTG4
C11orf88
LAYN
EUR Meta-analysis 11q23.3: Lead Variant chr11:116786845:C:T
Chromosome 11 (Mb)
APOA1
CEP164
APOC3
SIK3
PCSK7
BUD13
RNF214
APOA5
SIDT2
PAFAH1B2
TAGLN
BACE1
ZNF259
APOA4
EUR Meta-analysis 11q23.3: Lead Variant chr11:118791631:A:T
Chromosome 11 (Mb)
KMT2A
DDX6
H2AFX
IFT46
CBL
UBE4A
TMEM25
CXCR5
HMBS
ATP5L
ARCN1
BCL9L
HINFP
MPZL3
PHLDB1
UPK2
FOXR1
CD3G
TTC36
TREH
AMICA1
MPZL2
CD3D
CD3E
EUR Meta-analysis 12p13.33: Lead Variant chr12:870555:G:A
Chromosome 12 (Mb)
WNK1
RAD52
ERC1
CCDC77
B4GALNT3
NINJ2
$-\log_{10}(p)$
$r^2$
Index SNP
0.8 - 1.0
0.6 - 0.8
0.4 - 0.6
0.2 - 0.4
0.0 - 0.2
FINEMAP Probability
Credible Set
cs1
cs2
cs3
cs4
cs5
cs6
NA
PoPS Score
Score

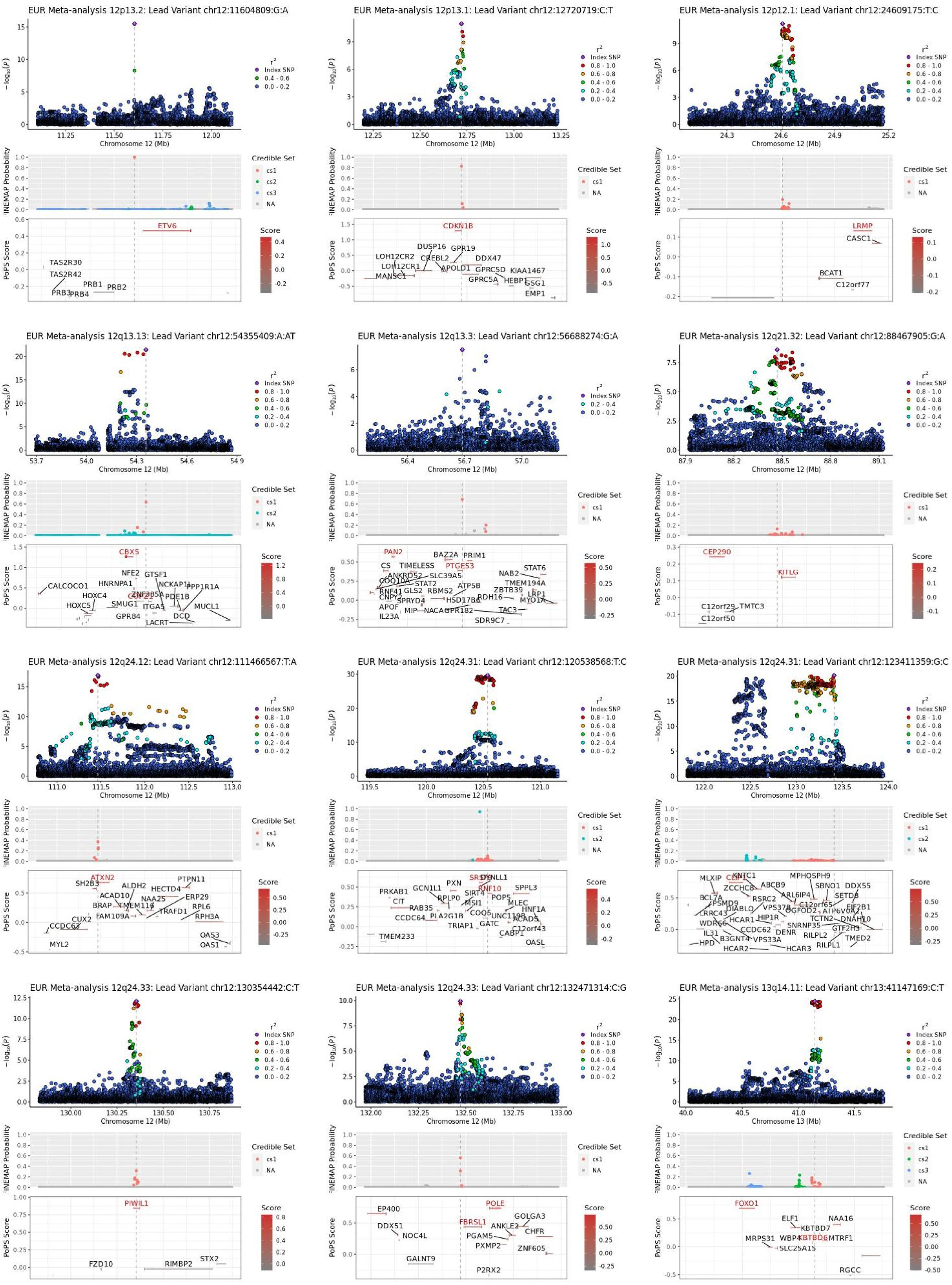
EUR Meta-analysis 12p13.2: Lead Variant chr12:11604809:G:A
ETV6
TAS2R30
TAS2R42
PRB1
PRB2
PRB3
PRB4
EUR Meta-analysis 12p13.1: Lead Variant chr12:12720719:C:T
CDKN1B
DUSP16
GPR19
LOH12CR2
LOH12CR1
CREBL2
APOLD1
DDX47
GPRC5D
KIAA1467
MANSC1
GPRC5A
HEBP1
GSG1
EMP1
EUR Meta-analysis 12p12.1: Lead Variant chr12:24609175:T:C
LRMP
CASC1
BCAT1
C12orf77
EUR Meta-analysis 12q13.13: Lead Variant chr12:54355409:A:AT
CBX5
NFE2
GTSF1
CALCOCO1
HNRNPA1
NCKAP1L
PPP1R1A
HOXC4
PDE1B
HOXC5
SMUG1
ITGA5
MUCL1
GPR84
DCD
LACRT
EUR Meta-analysis 12q13.3: Lead Variant chr12:56688274:G:A
PAN2
BAZ2A
PRIM1
CS
TIMELESS
PTGES3
STAT6
ANKRD52
SLC39A5
NAB2
STAT2
TMEM194A
RNF41
ATP5B
ZBTB39
CNPY2
GLS2
RBMS2
LRP1
SPRYD4
HSD17B6
RDH16
MYO1A
APOF
MIP
NACA
GPR182
TAC3
IL23A
SDR9C7
EUR Meta-analysis 12q21.32: Lead Variant chr12:88467905:G:A
CEP290
KITLG
C12orf29
TMTC3
C12orf50
EUR Meta-analysis 12q24.12: Lead Variant chr12:111466567:T:A
ATXN2
PTPN11
SH2B3
ALDH2
HECTD4
ACAD10
NAA25
ERP29
BRAP
TMEM116
RPL6
TRAFD1
FAM109A
RPH3A
CUX2
CCDC63
OAS3
MYL2
OAS1
EUR Meta-analysis 12q24.31: Lead Variant chr12:120538568:T:C
DYNLL1
PXN
SPPL3
PRKAB1
GCN1L1
RNF10
CIT
RPLP0
SIRT4
POP5
RAB35
MSI1
MLEC
CCDC64
PLA2G1B
COQ5
HNF1A
UNC119B
TRIAP1
GATC
ACADS
C12orf43
CABP1
TMEM233
OASL
EUR Meta-analysis 12q24.31: Lead Variant chr12:123411359:G:C
MLXIP
KNTC1
MPHOSPH9
ZCCHC8
ABCB9
SBNO1
DDX55
BCL7A
RSRC2
ARL6IP4
SETD8
PSMD9
C12orf65
DIABLO
VPS37B
OGFOD2
ATP6V0A2
EIF2B1
LRRC43
HCAR1
HIP1R
TCTN2
DNAH10
WDR66
CCDC62
SNRNP35
GTF2H3
IL31
DENR
RILPL2
HPD
B3GNT4
VPS33A
TMED2
HCAR2
HCAR3
RILPL1
EUR Meta-analysis 12q24.33: Lead Variant chr12:130354442:C:T
PIWIL1
FZD10
RIMBP2
STX2
EUR Meta-analysis 12q24.33: Lead Variant chr12:132471314:C:G
POLE
EP400
GOLGA3
FBRSL1
ANKLE2
DDX51
NOC4L
PGAM5
CHFR
PXMP2
ZNF605
GALNT9
P2RX2
EUR Meta-analysis 13q14.11: Lead Variant chr13:41147169:C:T
FOXO1
ELF1
NAA16
KBTBD7
MRPS31
WBP4
KBTBD6
MTRF1
SLC25A15
RGCC
Chromosome 12 (Mb)
Chromosome 13 (Mb)
$-\log_{10}(p)$
FINEMAP Probability
PoPS Score
$r^2$
Index SNP
0.8 - 1.0
0.6 - 0.8
0.4 - 0.6
0.2 - 0.4
0.0 - 0.2
Credible Set
cs1
cs2
cs3
NA
Score

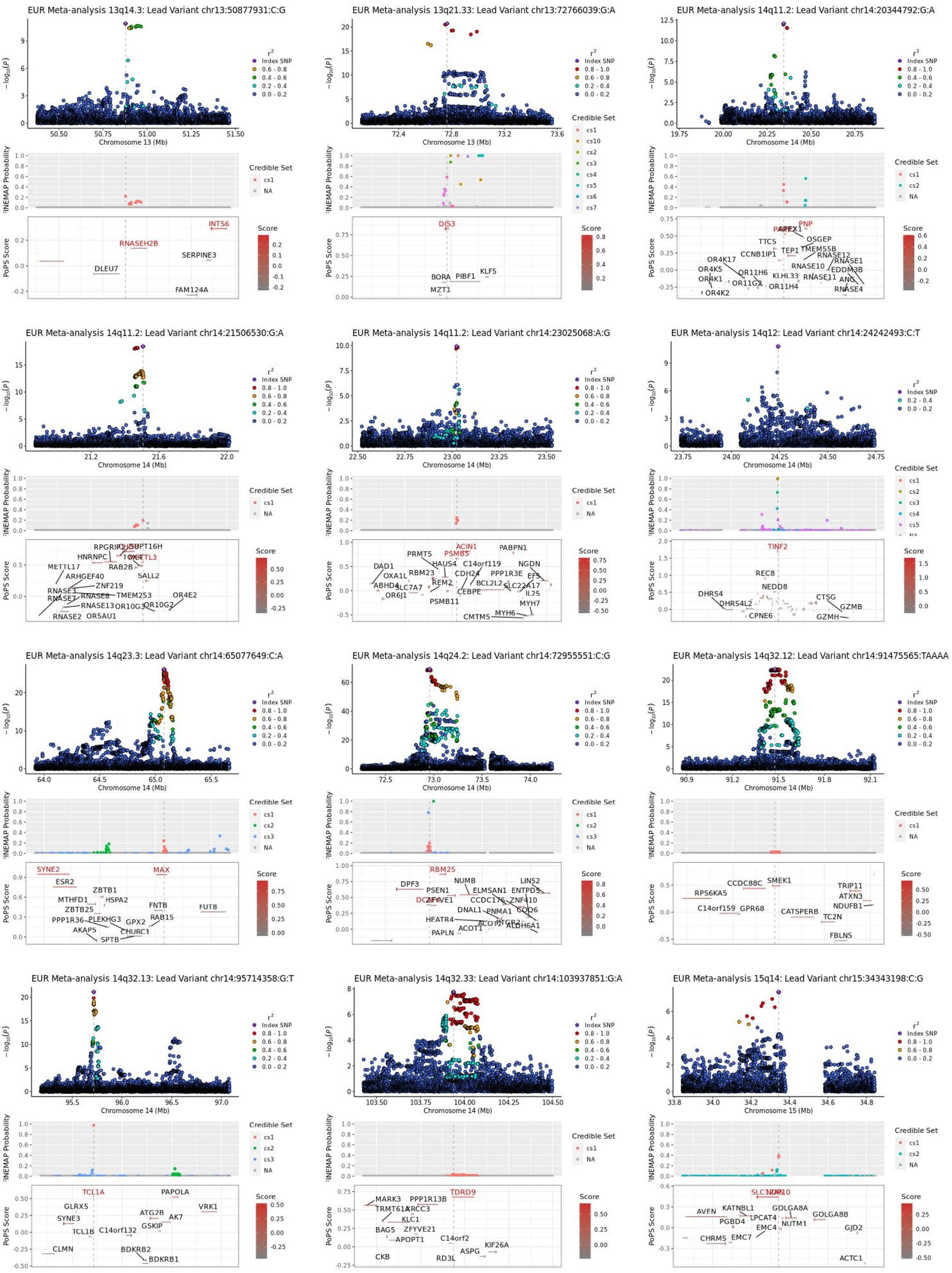

EUR Meta-analysis 13q14.3: Lead Variant chr13:50877931:C:G
Chromosome 13 (Mb)
INTS6
RNASEH2B
SERPINE3
DLEU7
FAM124A
EUR Meta-analysis 13q21.33: Lead Variant chr13:72766039:G:A
Chromosome 13 (Mb)
DIS3
BORA
PIBF1
KLF5
MZT1
EUR Meta-analysis 14q11.2: Lead Variant chr14:20344792:G:A
Chromosome 14 (Mb)
PNP
APEX1
OSGEP
TTC5
TEP1
TMEM55B
CCNB1IP1
RNASE12
RNASE1
RNASE10
EDDM3B
OR4K17
OR4K5
OR11H6
KLHL33
RNASE11
ANG
OR4K1
OR11G2
OR11H4
RNASE4
OR4K2
EUR Meta-analysis 14q11.2: Lead Variant chr14:21506530:G:A
Chromosome 14 (Mb)
RPGRIP1
SUPT16H
HNRNPC
TOX4
METTL3
RAB2B
METTL17
SALL2
ARHGEF40
ZNF219
RNASE3
RNASE8
TMEM253
OR4E2
RNASE7
RNASE13
OR10G3
OR10G2
OR5AU1
RNASE2
EUR Meta-analysis 14q11.2: Lead Variant chr14:23025068:A:G
Chromosome 14 (Mb)
ACIN1
PABPN1
PRMT5
PSMB5
HAUS4
C14orf119
NGDN
DAD1
OXA1L
RBM23
CDH24
PPP1R3E
EFS
ABHD4
REM2
BCL2L2
SLC22A17
SLC7A7
CEBPE
IL25
OR6J1
PSMB11
MYH7
MYH6
CMTM5
EUR Meta-analysis 14q12: Lead Variant chr14:24242493:C:T
Chromosome 14 (Mb)
TINF2
REC8
NEDD8
DHRS4
DHRS4L2
CTSG
GZMB
CPNE6
GZMH
EUR Meta-analysis 14q23.3: Lead Variant chr14:65077649:C:A
Chromosome 14 (Mb)
SYNE2
MAX
ESR2
ZBTB1
MTHFD1
HSPA2
ZBTB25
FNTB
FUT8
RAB15
PPP1R36
PLEKHG3
GPX2
CHURC1
AKAP5
SPTB
EUR Meta-analysis 14q24.2: Lead Variant chr14:72955551:C:G
Chromosome 14 (Mb)
RBM25
NUMB
LIN52
DPF3
PSEN1
ELMSAN1
ENTPD5
DCAF4
ZFYVE1
DNAL1
PNMA1
COQ6
CCDC176
ZNF410
HEATR4
ACOT1
PTGR2
ALDH6A1
PAPLN
ACOT2
EUR Meta-analysis 14q32.12: Lead Variant chr14:91475565:TAAAA
Chromosome 14 (Mb)
SMEK1
CCDC88C
TRIP11
RPS6KA5
ATXN3
NDUFB1
C14orf159
GPR68
CATSPERB
TC2N
FBLN5
EUR Meta-analysis 14q32.13: Lead Variant chr14:95714358:G:T
Chromosome 14 (Mb)
TCL1A
PAPOLA
GLRX5
VRK1
ATG2B
AK7
SYNE3
GSKIP
TCL1B
C14orf132
CLMN
BDKRB2
BDKRB1
EUR Meta-analysis 14q32.33: Lead Variant chr14:103937851:G:A
Chromosome 14 (Mb)
TDRD9
MARK3
PPP1R13B
TRMT61A
XRCC3
KLC1
BAG5
ZFYVE21
APOPT1
C14orf2
KIF26A
ASPG
CKB
RD3L
EUR Meta-analysis 15q14: Lead Variant chr15:34343198:C:G
Chromosome 15 (Mb)
SLC12A6
NOP10
AVEN
KATNBL1
GOLGA8A
GOLGA8B
LPCAT4
PGBD4
NUTM1
EMC4
GJD2
CHRM5
EMC7
ACTC1
r²
Index SNP
0.8 - 1.0
0.6 - 0.8
0.4 - 0.6
0.2 - 0.4
0.0 - 0.2
Credible Set
cs1
cs2
cs3
NA
Score
FINEMAP Probability
PoPS Score

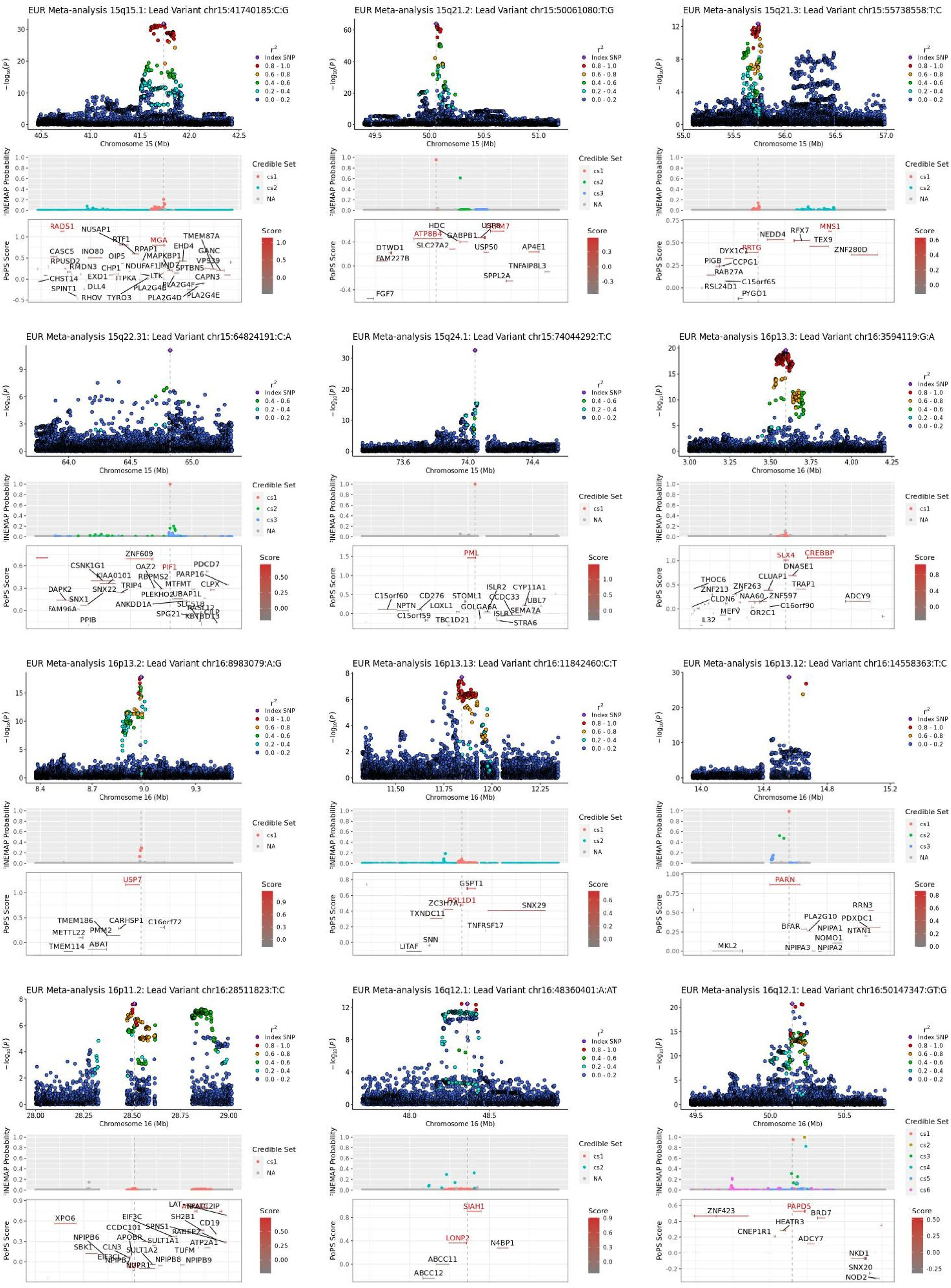

EUR Meta-analysis 15q15.1: Lead Variant chr15:41740185:C:G
EUR Meta-analysis 15q21.2: Lead Variant chr15:50061080:T:G
EUR Meta-analysis 15q21.3: Lead Variant chr15:55738558:T:C
EUR Meta-analysis 15q22.31: Lead Variant chr15:64824191:C:A
EUR Meta-analysis 15q24.1: Lead Variant chr15:74044292:T:C
EUR Meta-analysis 16p13.3: Lead Variant chr16:3594119:G:A
EUR Meta-analysis 16p13.2: Lead Variant chr16:8983079:A:G
EUR Meta-analysis 16p13.13: Lead Variant chr16:11842460:C:T
EUR Meta-analysis 16p13.12: Lead Variant chr16:14558363:T:C
EUR Meta-analysis 16p11.2: Lead Variant chr16:28511823:T:C
EUR Meta-analysis 16q12.1: Lead Variant chr16:48360401:A:AT
EUR Meta-analysis 16q12.1: Lead Variant chr16:50147347:GT:G
$-\log_{10}(P)$
Chromosome 15 (Mb)
Chromosome 16 (Mb)
$r^2$
Index SNP
0.8 - 1.0
0.6 - 0.8
0.4 - 0.6
0.2 - 0.4
0.0 - 0.2
FINEMAP Probability
Credible Set
cs1
cs2
cs3
cs4
cs5
cs6
NA
PoPS Score
Score
RAD51
NUSAP1
MGA
TMEM87A
ATP8B4
HDC
USP8
MNS1
RRTG
NEDD4
RFX7
TEX9
ZNF280D
ZNF609
PIF1
PML
SLX4
CREBBP
USP7
GSPT1
RSL1D1
SNX29
PARN
XPO6
SIAH1
LONP2
N4BP1
ABCC11
ABCC12
PAPD5
ZNF423
BRD7
HEATR3
NKD1
SNX20
NOD2

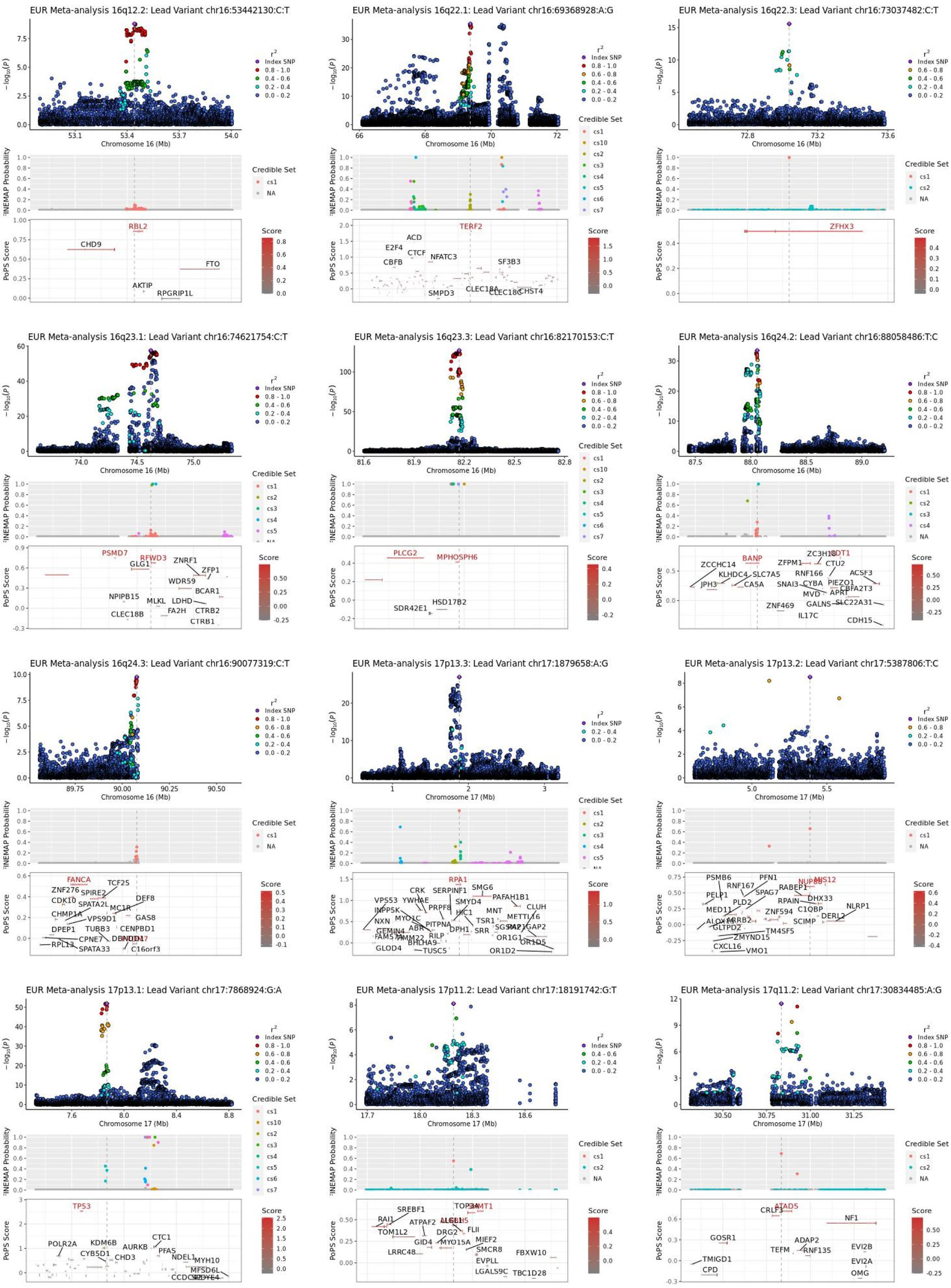

EUR Meta-analysis 16q12.2: Lead Variant chr16:53442130:C:T
Chromosome 16 (Mb)
CHD9
RBL2
FTO
AKTIP
RPGRIP1L
EUR Meta-analysis 16q22.1: Lead Variant chr16:69368928:A:G
Chromosome 16 (Mb)
ACD
E2F4
CTCF
CBFB
NFATC3
TERF2
SF3B3
SMPD3
CLEC18A
CLEC18C
CHST4
EUR Meta-analysis 16q22.3: Lead Variant chr16:73037482:C:T
Chromosome 16 (Mb)
ZFHX3
EUR Meta-analysis 16q23.1: Lead Variant chr16:74621754:C:T
Chromosome 16 (Mb)
PSMD7
RFWD3
GLG1
ZNRF1
ZFP1
WDR59
BCAR1
NPIPB15
MLKL
LDHD
FA2H
CTRB2
CLEC18B
CTRB1
EUR Meta-analysis 16q23.3: Lead Variant chr16:82170153:C:T
Chromosome 16 (Mb)
PLCG2
MPHOSPH6
HSD17B2
SDR42E1
EUR Meta-analysis 16q24.2: Lead Variant chr16:88058486:T:C
Chromosome 16 (Mb)
BANP
ZCCHC14
KLHDC4
SLC7A5
JPH3
CA5A
ZFPM1
ZC3H18
CTU2
RNF166
CYBA
PIEZO1
ACSF3
CBFA2T3
SNAI3
MVD
APRT
ZNF469
GALNS
SLC22A31
IL17C
CDH15
EUR Meta-analysis 16q24.3: Lead Variant chr16:90077319:C:T
Chromosome 16 (Mb)
FANCA
ZNF276
SPIRE2
TCF25
CDK10
SPATA2L
MC1R
DEF8
CHMP1A
VPS9D1
GAS8
DPEP1
TUBB3
CENPBD1
CPNE7
RPL13
SPATA33
C16orf3
EUR Meta-analysis 17p13.3: Lead Variant chr17:1879658:A:G
Chromosome 17 (Mb)
RPA1
CRK
SERPINF1
SMG6
VPS53
YWHAE
SMYD4
PAFAH1B1
INPP5K
PRPF8
HIC1
CLUH
NXN
MYO1C
PITPNA
MNT
METTL16
ABR
TSR1
GEMIN4
DPH1
SRR
SGSM2
RAP1GAP2
FAM57A
TIMM22
RILP
OR1G1
OR1D5
BHLHA9
GLOD4
TUSC5
OR1D2
EUR Meta-analysis 17p13.2: Lead Variant chr17:5387806:T:C
Chromosome 17 (Mb)
PSMB6
PFN1
RNF167
NUP88
MIS12
RABEP1
PELP1
SPAG7
PLD2
RPAIN
DHX33
MED11
C1QBP
NLRP1
ALOX15
ARRB2
ZNF594
SCIMP
DERL2
GLTPD2
TM4SF5
ZMYND15
CXCL16
VMO1
EUR Meta-analysis 17p13.1: Lead Variant chr17:7868924:G:A
Chromosome 17 (Mb)
TP53
POLR2A
KDM6B
CYB5D1
AURKB
CTC1
CHD3
PFAS
NDEL1
MYH10
MFSD6L
CCDC42
EUR Meta-analysis 17p11.2: Lead Variant chr17:18191742:G:T
Chromosome 17 (Mb)
SREBF1
TOP3A
RAI1
ATPAF2
TOM1L2
DRG2
FLII
GID4
MYO15A
MIEF2
LRRC48
SMCR8
FBXW10
EVPLL
LGALS9C
TBC1D28
EUR Meta-analysis 17q11.2: Lead Variant chr17:30834485:A:G
Chromosome 17 (Mb)
ATAD5
CRLF3
NF1
GOSR1
ADAP2
TEFM
RNF135
EVI2B
TMIGD1
EVI2A
CPD
OMG
$-\log_{10}(p)$
FINEMAP Probability
PoPS Score
$r^2$
Index SNP
0.8 - 1.0
0.6 - 0.8
0.4 - 0.6
0.2 - 0.4
0.0 - 0.2
Credible Set
cs1
cs2
cs3
cs4
cs5
cs6
cs7
cs10
NA
Score

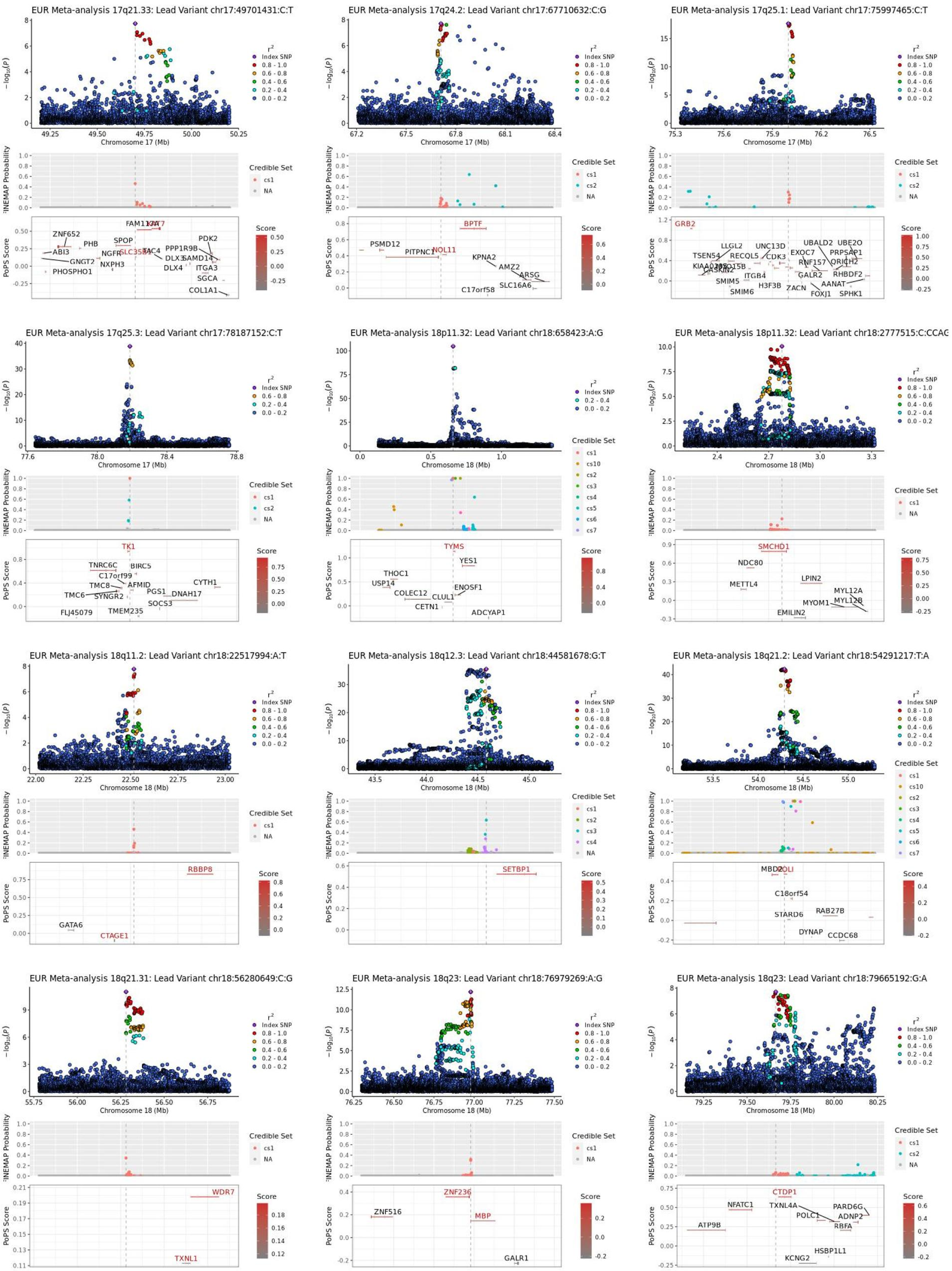

EUR Meta-analysis 17q21.33: Lead Variant chr17:49701431:C:T
EUR Meta-analysis 17q24.2: Lead Variant chr17:67710632:C:G
EUR Meta-analysis 17q25.1: Lead Variant chr17:75997465:C:T
EUR Meta-analysis 17q25.3: Lead Variant chr17:78187152:C:T
EUR Meta-analysis 18p11.32: Lead Variant chr18:658423:A:G
EUR Meta-analysis 18p11.32: Lead Variant chr18:2777515:C:CCAG
EUR Meta-analysis 18q11.2: Lead Variant chr18:22517994:A:T
EUR Meta-analysis 18q12.3: Lead Variant chr18:44581678:G:T
EUR Meta-analysis 18q21.2: Lead Variant chr18:54291217:T:A
EUR Meta-analysis 18q21.31: Lead Variant chr18:56280649:C:G
EUR Meta-analysis 18q23: Lead Variant chr18:76979269:A:G
EUR Meta-analysis 18q23: Lead Variant chr18:79665192:G:A
Chromosome 17 (Mb)
Chromosome 18 (Mb)
FINEMAP Probability
PoPS Score
Credible Set
Score
Index SNP
0.8 - 1.0
0.6 - 0.8
0.4 - 0.6
0.2 - 0.4
0.0 - 0.2
ZNF652
PHB
ABI3
GNGT2
PHOSPHO1
NGFR
NXPH3
SPOP
SLC35B1
FAM117A
TAC4
DLX3
DLX4
PPP1R9B
SAMD14
PDK2
ITGA3
SGCA
COL1A1
PSMD12
PITPNC1
NOL11
BPTF
KPNA2
AMZ2
ARSG
SLC16A6
C17orf58
GRB2
LLGL2
TSEN54
UNC13D
RECQL5
CDK3
CASKIN2
ITGB4
SMIM5
SMIM6
H3F3B
EXOC7
UBALD2
UBE2O
PRPSAP1
RNF157
QRICH2
GALR2
RHBDF2
ZACN
AANAT
FOXJ1
SPHK1
TK1
TNRC6C
BIRC5
C17orf99
TMC8
AFMID
TMC6
SYNGR2
PGS1
DNAH17
CYTH1
SOCS3
FLJ45079
TMEM235
TYMS
YES1
THOC1
USP14
COLEC12
ENOSF1
CLUL1
CETN1
ADCYAP1
SMCHD1
NDC80
METTL4
LPIN2
MYL12A
MYL12B
MYOM1
EMILIN2
RBBP8
GATA6
CTAGE1
SETBP1
MBD2
POLI
C18orf54
STARD6
RAB27B
DYNAP
CCDC68
WDR7
TXNL1
ZNF236
ZNF516
MBP
GALR1
CTDP1
NFATC1
TXNL4A
PARD6G
ADNP2
POLC1
RBFA
ATP9B
HSBP1L1
KCNG2

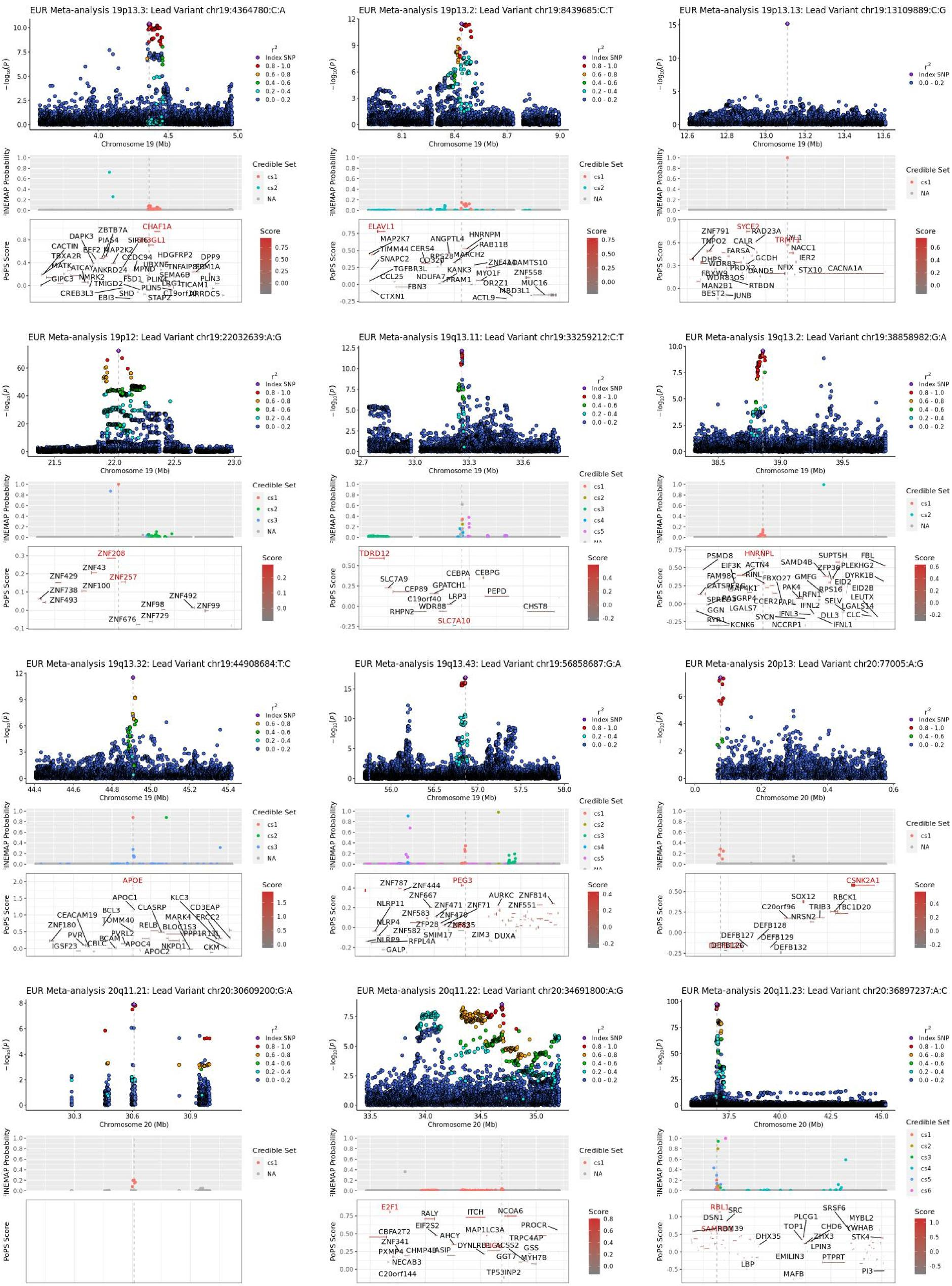

EUR Meta-analysis 19p13.3: Lead Variant chr19:4364780:C:A
EUR Meta-analysis 19p13.2: Lead Variant chr19:8439685:C:T
EUR Meta-analysis 19p13.13: Lead Variant chr19:13109889:C:G
EUR Meta-analysis 19p12: Lead Variant chr19:22032639:A:G
EUR Meta-analysis 19q13.11: Lead Variant chr19:33259212:C:T
EUR Meta-analysis 19q13.2: Lead Variant chr19:38858982:G:A
EUR Meta-analysis 19q13.32: Lead Variant chr19:44908684:T:C
EUR Meta-analysis 19q13.43: Lead Variant chr19:56858687:G:A
EUR Meta-analysis 20p13: Lead Variant chr20:77005:A:G
EUR Meta-analysis 20q11.21: Lead Variant chr20:30609200:G:A
EUR Meta-analysis 20q11.22: Lead Variant chr20:34691800:A:G
EUR Meta-analysis 20q11.23: Lead Variant chr20:36897237:A:C
Chromosome 19 (Mb)
Chromosome 20 (Mb)
FINEMAP Probability
PoPS Score
Credible Set
Score
Index SNP

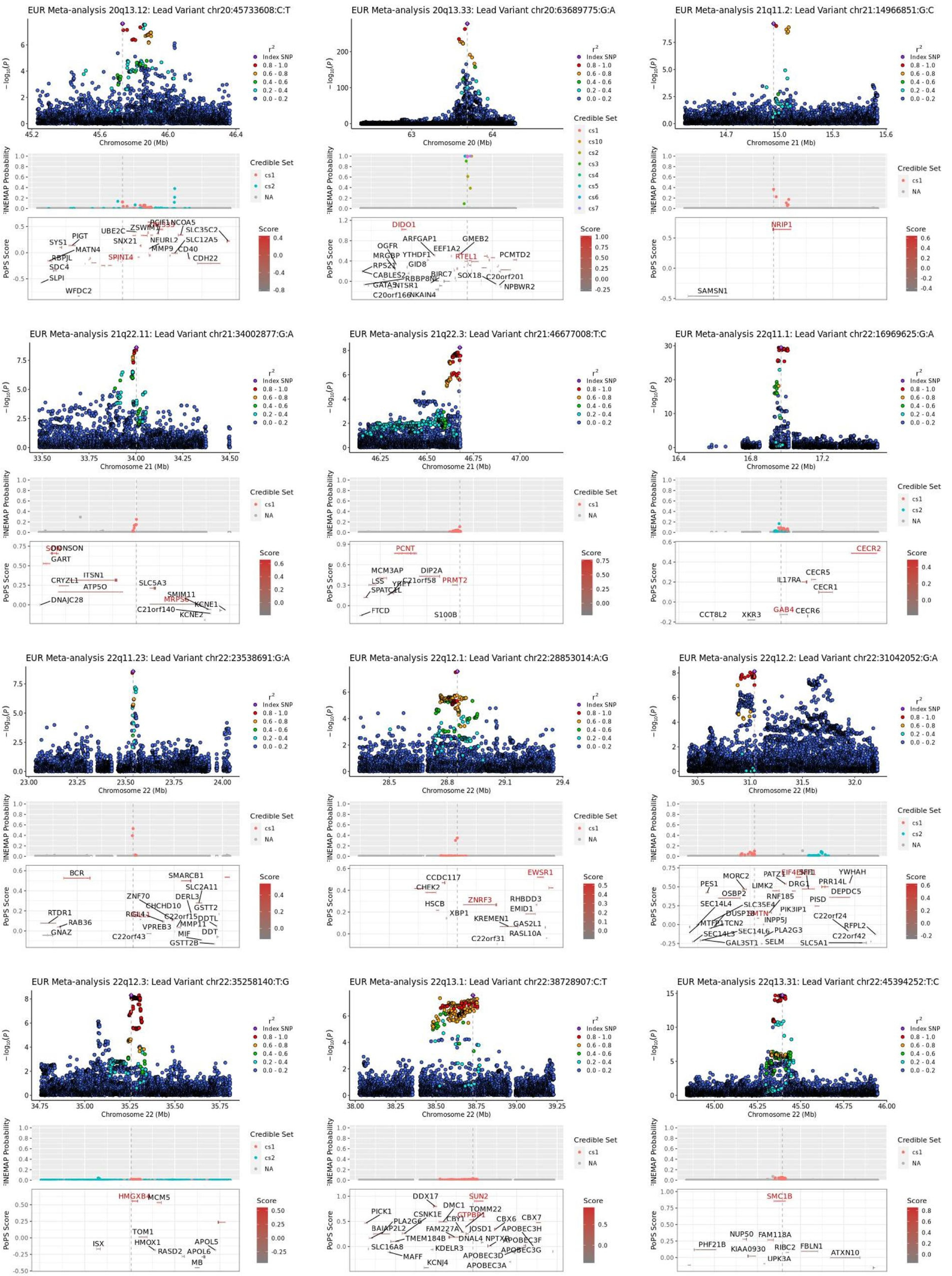
EUR Meta-analysis 20q13.12: Lead Variant chr20:45733608:C:T
EUR Meta-analysis 20q13.33: Lead Variant chr20:63689775:G:A
EUR Meta-analysis 21q11.2: Lead Variant chr21:14966851:G:C
EUR Meta-analysis 21q22.11: Lead Variant chr21:34002877:G:A
EUR Meta-analysis 21q22.3: Lead Variant chr21:46677008:T:C
EUR Meta-analysis 22q11.1: Lead Variant chr22:16969625:G:A
EUR Meta-analysis 22q11.23: Lead Variant chr22:23538691:G:A
EUR Meta-analysis 22q12.1: Lead Variant chr22:28853014:A:G
EUR Meta-analysis 22q12.2: Lead Variant chr22:31042052:G:A
EUR Meta-analysis 22q12.3: Lead Variant chr22:35258140:T:G
EUR Meta-analysis 22q13.1: Lead Variant chr22:38728907:C:T
EUR Meta-analysis 22q13.31: Lead Variant chr22:45394252:T:C
$-\log_{10}(p)$
Chromosome 20 (Mb)
Chromosome 21 (Mb)
Chromosome 22 (Mb)
$r^2$
Index SNP
0.8 - 1.0
0.6 - 0.8
0.4 - 0.6
0.2 - 0.4
0.0 - 0.2
FINEMAP Probability
Credible Set
cs1
cs2
NA
PoPS Score
Score
SPINT4
DIDO1
RTEL1
NRIP1
PCNT
PRMT2
CECR2
GAB4
BCR
EWSR1
ZNRF3
HMGXB4
SUN2
SMC1B

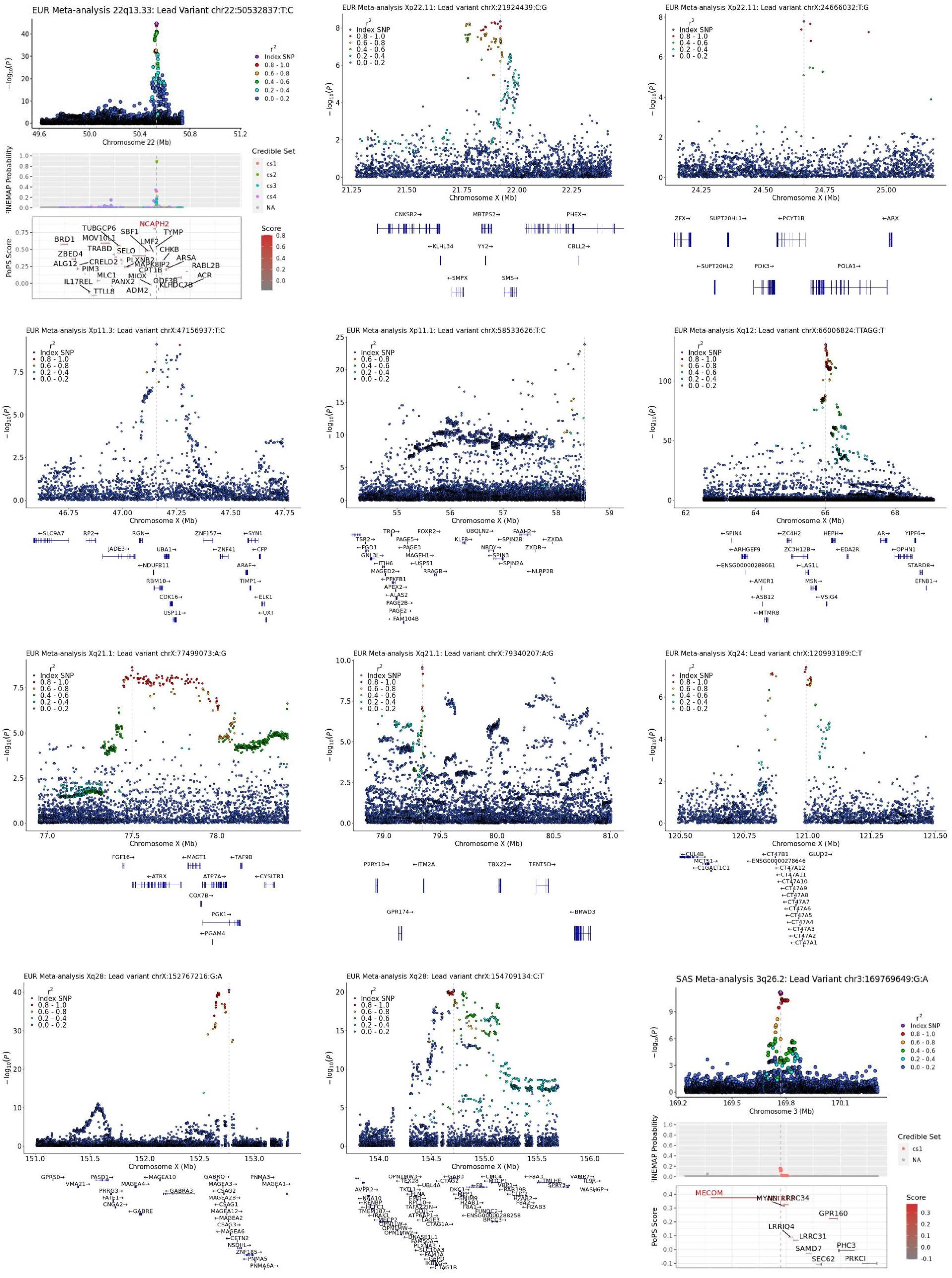

EUR Meta-analysis 22q13.33: Lead Variant chr22:50532837:T:C
EUR Meta-analysis Xp22.11: Lead variant chrX:21924439:C:G
EUR Meta-analysis Xp22.11: Lead variant chrX:24666032:T:G
EUR Meta-analysis Xp11.3: Lead variant chrX:47156937:T:C
EUR Meta-analysis Xp11.1: Lead variant chrX:58533626:T:C
EUR Meta-analysis Xq12: Lead variant chrX:66006824:TTAGG:T
EUR Meta-analysis Xq21.1: Lead variant chrX:77499073:A:G
EUR Meta-analysis Xq21.1: Lead variant chrX:79340207:A:G
EUR Meta-analysis Xq24: Lead variant chrX:120993189:C:T
EUR Meta-analysis Xq28: Lead variant chrX:152767216:G:A
EUR Meta-analysis Xq28: Lead variant chrX:154709134:C:T
SAS Meta-analysis 3q26.2: Lead Variant chr3:169769649:G:A
Chromosome 22 (Mb)
Chromosome X (Mb)
Chromosome 3 (Mb)
Index SNP
0.8 - 1.0
0.6 - 0.8
0.4 - 0.6
0.2 - 0.4
0.0 - 0.2
Credible Set
PoPS Score
Score
NCAPH2
MECOM

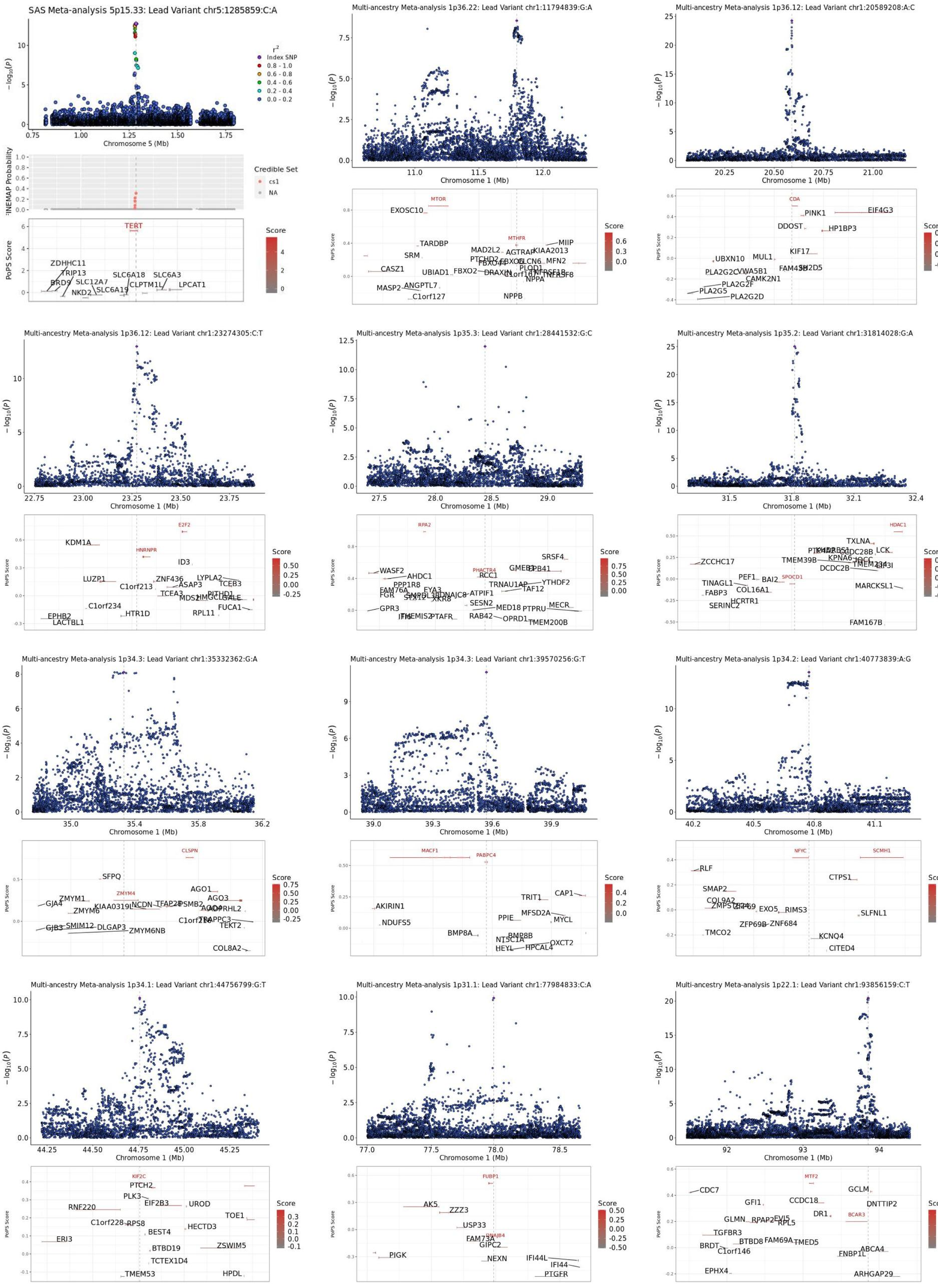

SAS Meta-analysis 5p15.33: Lead Variant chr5:1285859:C:A
Chromosome 5 (Mb)
r2
Index SNP
0.8 - 1.0
0.6 - 0.8
0.4 - 0.6
0.2 - 0.4
0.0 - 0.2
FINEMAP Probability
Credible Set
cs1
NA
PoPS Score
Score
TERT
Multi-ancestry Meta-analysis 1p36.22: Lead Variant chr1:11794839:G:A
Chromosome 1 (Mb)
MTOR
MTHFR
Multi-ancestry Meta-analysis 1p36.12: Lead Variant chr1:20589208:A:C
CDA
Multi-ancestry Meta-analysis 1p36.12: Lead Variant chr1:23274305:C:T
E2F2
HNRNPR
Multi-ancestry Meta-analysis 1p35.3: Lead Variant chr1:28441532:G:C
RPA2
PHACTR4
Multi-ancestry Meta-analysis 1p35.2: Lead Variant chr1:31814028:G:A
HDAC1
SPOCD1
Multi-ancestry Meta-analysis 1p34.3: Lead Variant chr1:35332362:G:A
CLSPN
ZMYM4
Multi-ancestry Meta-analysis 1p34.3: Lead Variant chr1:39570256:G:T
MACF1
PABPC4
Multi-ancestry Meta-analysis 1p34.2: Lead Variant chr1:40773839:A:G
NFYC
SCMH1
Multi-ancestry Meta-analysis 1p34.1: Lead Variant chr1:44756799:G:T
KIF2C
Multi-ancestry Meta-analysis 1p31.1: Lead Variant chr1:77984833:C:A
FUBP1
DNAJB4
Multi-ancestry Meta-analysis 1p22.1: Lead Variant chr1:93856159:C:T
MTF2
BCAR3

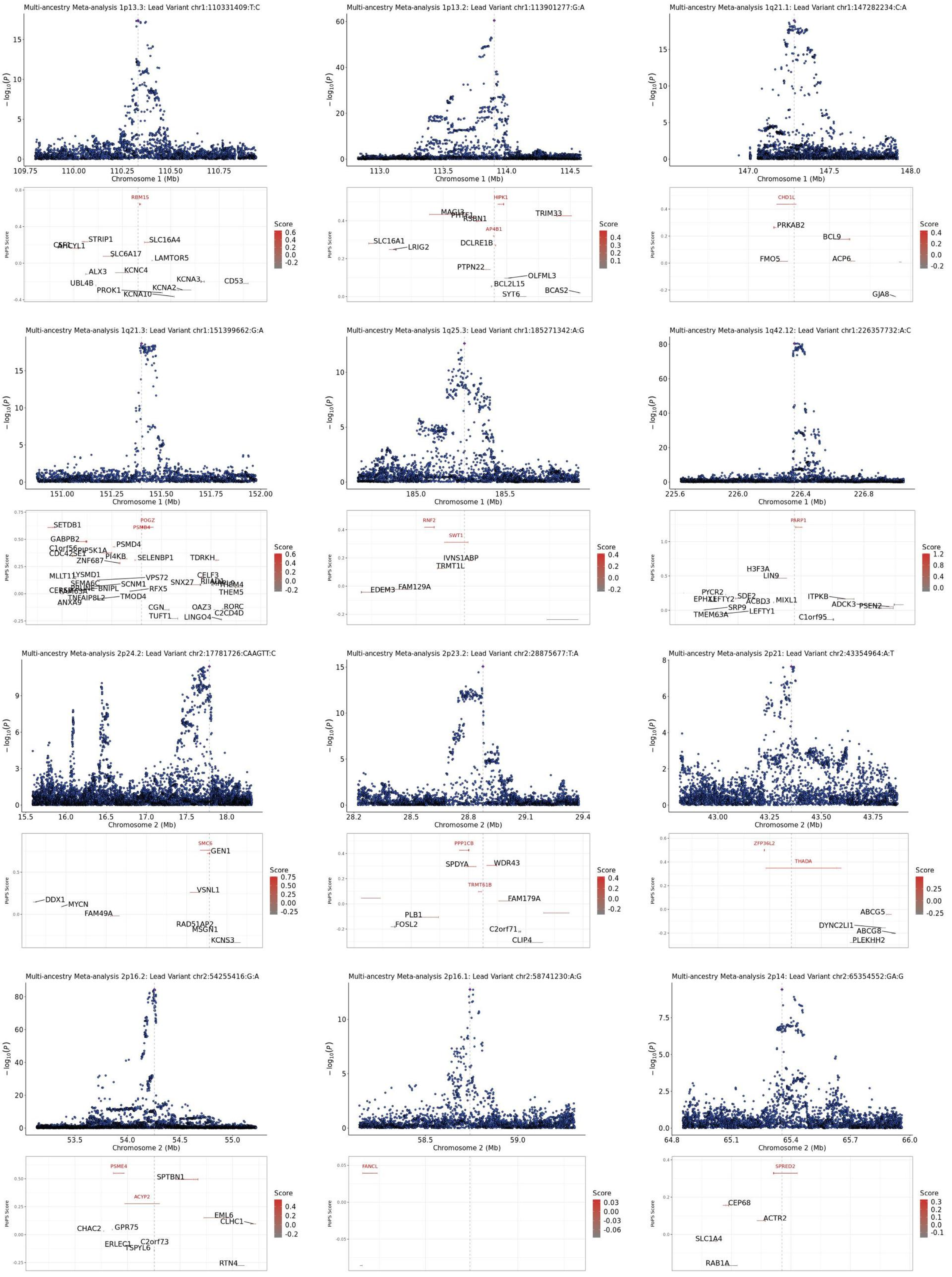

Multi-ancestry Meta-analysis 1p13.3: Lead Variant chr1:110331409:T:C
Chromosome 1 (Mb)
RBM15
STRIP1
SLC16A4
SLC6A17
LAMTOR5
ALX3
KCNC4
UBL4B
KCNA3
CD53
PROK1
KCNA2
KCNA10
Multi-ancestry Meta-analysis 1p13.2: Lead Variant chr1:113901277:G:A
Chromosome 1 (Mb)
HIPK1
MAGI3
RSBN1
TRIM33
AP4B1
SLC16A1
LRIG2
DCLRE1B
PTPN22
OLFML3
BCL2L15
BCAS2
SYT6
Multi-ancestry Meta-analysis 1q21.1: Lead Variant chr1:147282234:C:A
Chromosome 1 (Mb)
CHD1L
PRKAB2
BCL9
FMO5
ACP6
GJA8
Multi-ancestry Meta-analysis 1q21.3: Lead Variant chr1:151399662:G:A
Chromosome 1 (Mb)
POGZ
PSMB4
SETDB1
GABPB2
PSMD4
PIP5K1A
ZNF687
PI4KB
SELENBP1
TDRKH
MLLT11
LYSMD1
VPS72
CELF3
SEMA6C
SCNM1
SNX27
THEM4
THEM5
BNIPL
RFX5
TNFAIP8L2
TMOD4
ANXA9
CGN
OAZ3
RORC
TUFT1
LINGO4
C2CD4D
Multi-ancestry Meta-analysis 1q25.3: Lead Variant chr1:185271342:A:G
Chromosome 1 (Mb)
RNF2
SWT1
IVNS1ABP
TRMT1L
EDEM3
FAM129A
Multi-ancestry Meta-analysis 1q42.12: Lead Variant chr1:226357732:A:C
Chromosome 1 (Mb)
PARP1
H3F3A
LIN9
PYCR2
SDE2
ACBD3
MIXL1
ITPKB
ADCK3
PSEN2
SRP9
LEFTY1
TMEM63A
C1orf95
Multi-ancestry Meta-analysis 2p24.2: Lead Variant chr2:17781726:CAAGTT:C
Chromosome 2 (Mb)
SMC6
GEN1
VSNL1
DDX1
MYCN
FAM49A
RAD51AP2
MSGN1
KCNS3
Multi-ancestry Meta-analysis 2p23.2: Lead Variant chr2:28875677:T:A
Chromosome 2 (Mb)
PPP1CB
SPDYA
WDR43
TRMT61B
FAM179A
PLB1
FOSL2
C2orf71
CLIP4
Multi-ancestry Meta-analysis 2p21: Lead Variant chr2:43354964:A:T
Chromosome 2 (Mb)
ZFP36L2
THADA
ABCG5
DYNC2LI1
ABCG8
PLEKHH2
Multi-ancestry Meta-analysis 2p16.2: Lead Variant chr2:54255416:G:A
Chromosome 2 (Mb)
PSME4
SPTBN1
ACYP2
EML6
CLHC1
CHAC2
GPR75
ERLEC1
C2orf73
TSPYL6
RTN4
Multi-ancestry Meta-analysis 2p16.1: Lead Variant chr2:58741230:A:G
Chromosome 2 (Mb)
FANCL
Multi-ancestry Meta-analysis 2p14: Lead Variant chr2:65354552:GA:G
Chromosome 2 (Mb)
SPRED2
CEP68
ACTR2
SLC1A4
RAB1A

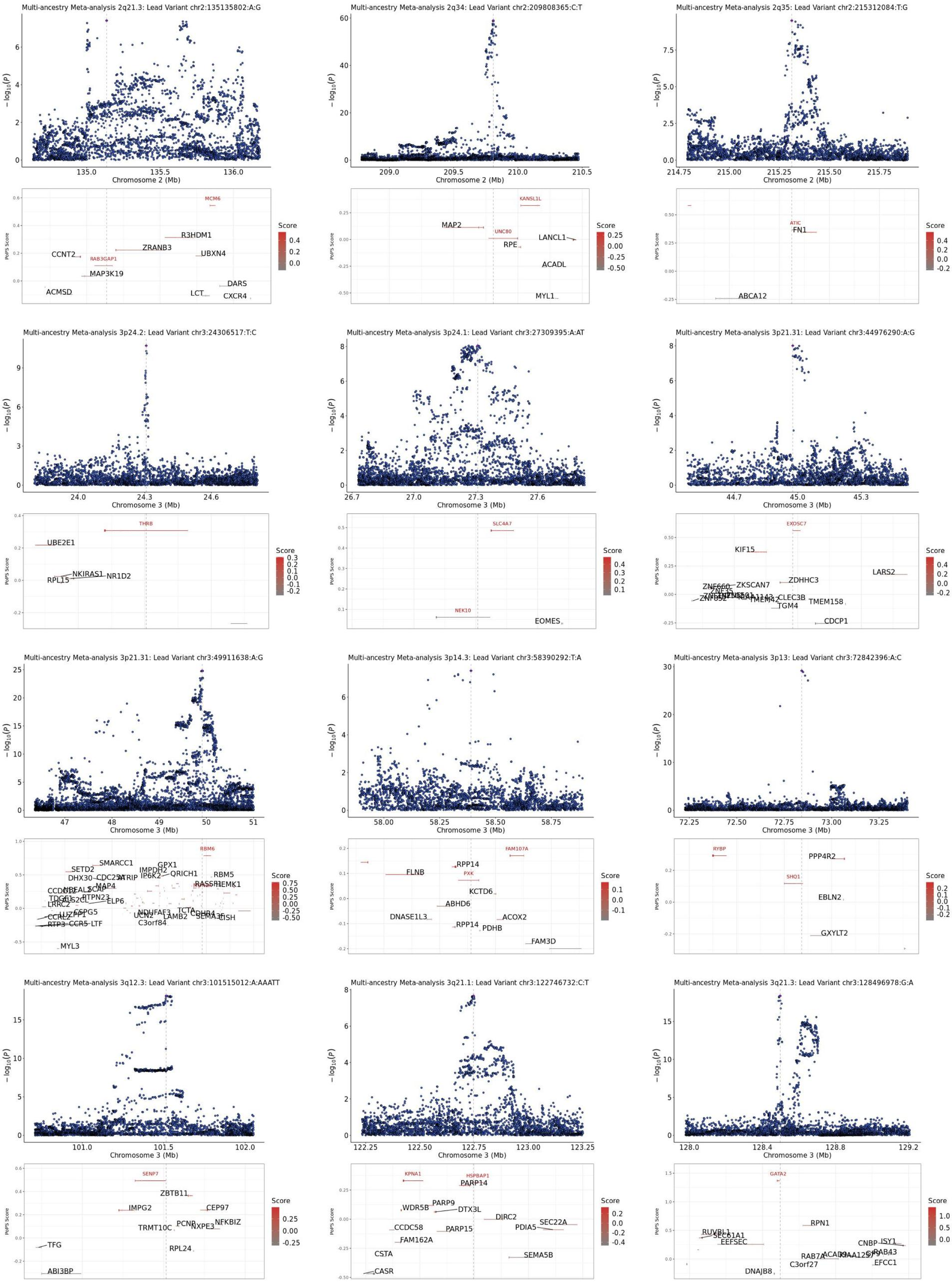

Multi-ancestry Meta-analysis 2q21.3: Lead Variant chr2:135135802:A:G
Chromosome 2 (Mb)
MCM6
R3HDM1
ZRANB3
UBXN4
CCNT2
RAB3GAP1
MAP3K19
DARS
ACMSD
LCT
CXCR4
Multi-ancestry Meta-analysis 2q34: Lead Variant chr2:209808365:C:T
Chromosome 2 (Mb)
KANSL1L
MAP2
UNC80
LANCL1
RPE
ACADL
MYL1
Multi-ancestry Meta-analysis 2q35: Lead Variant chr2:215312084:T:G
Chromosome 2 (Mb)
ATIC
FN1
ABCA12
Multi-ancestry Meta-analysis 3p24.2: Lead Variant chr3:24306517:T:C
Chromosome 3 (Mb)
THRB
UBE2E1
NKIRAS1
NR1D2
RPL15
Multi-ancestry Meta-analysis 3p24.1: Lead Variant chr3:27309395:A:AT
Chromosome 3 (Mb)
SLC4A7
NEK10
EOMES
Multi-ancestry Meta-analysis 3p21.31: Lead Variant chr3:44976290:A:G
Chromosome 3 (Mb)
EXOSC7
KIF15
LARS2
ZDHHC3
ZKSCAN7
CLEC3B
TMEM158
TMEM42
TGM4
CDCP1
Multi-ancestry Meta-analysis 3p21.31: Lead Variant chr3:49911638:A:G
Chromosome 3 (Mb)
RBM6
SMARCC1
SETD2
GPX1
IMPDH2
DHX30
CDC25A
ATRIP
IP6K2
QRICH1
RBM5
MAP4
RASSF1
HEMK1
PTPN23
ELP6
LRRC2
CSPG5
NDUFAF3
TCTA
CCRL2
UCN2
LAMB2
C3orf84
RTP3
CCR5
LTF
MYL3
Multi-ancestry Meta-analysis 3p14.3: Lead Variant chr3:58390292:T:A
Chromosome 3 (Mb)
FAM107A
RPP14
FLNB
PXK
KCTD6
ABHD6
DNASE1L3
ACOX2
PDHB
FAM3D
Multi-ancestry Meta-analysis 3p13: Lead Variant chr3:72842396:A:C
Chromosome 3 (Mb)
RYBP
PPP4R2
SHQ1
EBLN2
GXYLT2
Multi-ancestry Meta-analysis 3q12.3: Lead Variant chr3:101515012:A:AAATT
Chromosome 3 (Mb)
SENP7
ZBTB11
IMPG2
CEP97
NFKBIZ
PCNP
NXPE3
TRMT10C
TFG
RPL24
ABI3BP
Multi-ancestry Meta-analysis 3q21.1: Lead Variant chr3:122746732:C:T
Chromosome 3 (Mb)
KPNA1
HSPBAP1
PARP14
PARP9
WDR5B
DTX3L
DIRC2
SEC22A
CCDC58
PDIA5
PARP15
FAM162A
CSTA
SEMA5B
CASR
Multi-ancestry Meta-analysis 3q21.3: Lead Variant chr3:128496978:G:A
Chromosome 3 (Mb)
GATA2
RPN1
RUVBL1
SEC61A1
EEFSEC
CNBP
ISY1
RAB43
RAB7A
ACAD9
C3orf27
EFCC1
DNAJB8
PoPS Score
Score

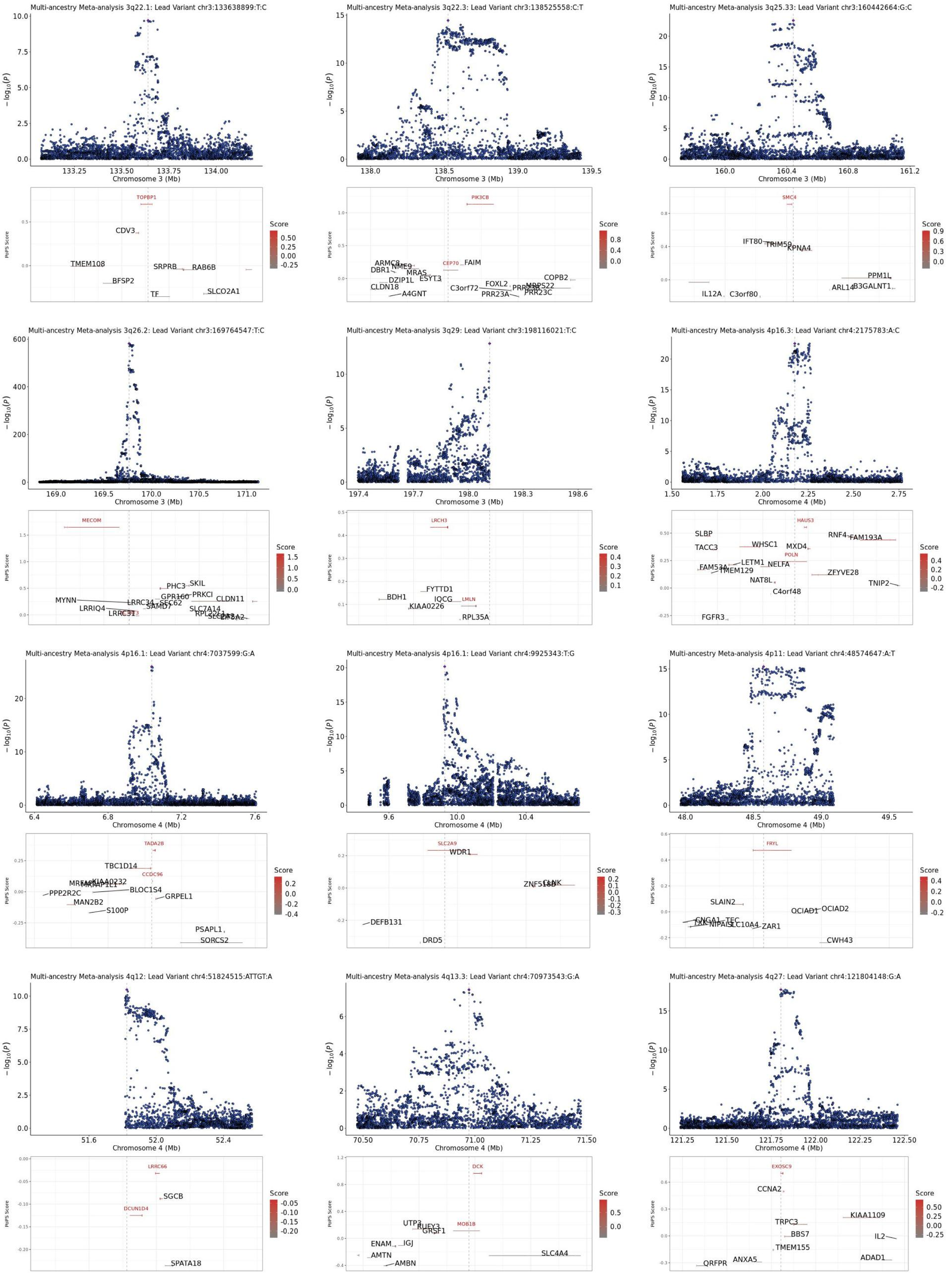

Multi-ancestry Meta-analysis 3q22.1: Lead Variant chr3:133638899:T:C
Chromosome 3 (Mb)
TOPBP1
CDV3
TMEM108
BFSP2
SRPRB
RAB6B
TF
SLCO2A1
Multi-ancestry Meta-analysis 3q22.3: Lead Variant chr3:138525558:C:T
PIK3CB
ARMC8
NME9
DBR1
MRAS
ESYT3
DZIP1L
CLDN18
A4GNT
CEP70
FAIM
FOXL2
C3orf72
PRR23A
PRR23C
MRPS22
COPB2
Multi-ancestry Meta-analysis 3q25.33: Lead Variant chr3:160442664:G:C
SMC4
IFT80
TRIM59
KPNA4
IL12A
C3orf80
PPM1L
ARL14
B3GALNT1
Multi-ancestry Meta-analysis 3q26.2: Lead Variant chr3:169764547:T:C
MECOM
PHC3
SKIL
GPR160
PRKCI
CLDN11
MYNN
LRRC34
SEC62
SAMD7
LRRIQ4
LRRC31
SLC7A14
RPL22L1
Multi-ancestry Meta-analysis 3q29: Lead Variant chr3:198116021:T:C
LRCH3
FYTTD1
BDH1
IQCG
LMLN
KIAA0226
RPL35A
Multi-ancestry Meta-analysis 4p16.3: Lead Variant chr4:2175783:A:C
Chromosome 4 (Mb)
HAUS3
SLBP
RNF4
FAM193A
TACC3
WHSC1
MXD4
POLN
LETM1
NELFA
FAM53A
TMEM129
ZFYVE28
NAT8L
TNIP2
C4orf48
FGFR3
Multi-ancestry Meta-analysis 4p16.1: Lead Variant chr4:7037599:G:A
TADA2B
TBC1D14
CCDC96
KIAA0232
PPP2R2C
BLOC1S4
GRPEL1
MAN2B2
S100P
PSAPL1
SORCS2
Multi-ancestry Meta-analysis 4p16.1: Lead Variant chr4:9925343:T:G
SLC2A9
WDR1
DEFB131
DRD5
Multi-ancestry Meta-analysis 4p11: Lead Variant chr4:48574647:A:T
FRYL
SLAIN2
OCIAD1
OCIAD2
CNGA1
TEC
NIPAL1
SLC10A4
ZAR1
CWH43
Multi-ancestry Meta-analysis 4q12: Lead Variant chr4:51824515:ATTGT:A
LRRC66
SGCB
DCUN1D4
SPATA18
Multi-ancestry Meta-analysis 4q13.3: Lead Variant chr4:70973543:G:A
DCK
UTP3
RUFY3
GRSF1
MOB1B
ENAM
IGJ
AMTN
AMBN
SLC4A4
Multi-ancestry Meta-analysis 4q27: Lead Variant chr4:121804148:G:A
EXOSC9
CCNA2
KIAA1109
TRPC3
BBS7
IL2
TMEM155
QRFPR
ANXA5
ADAD1
PoPS Score
Score

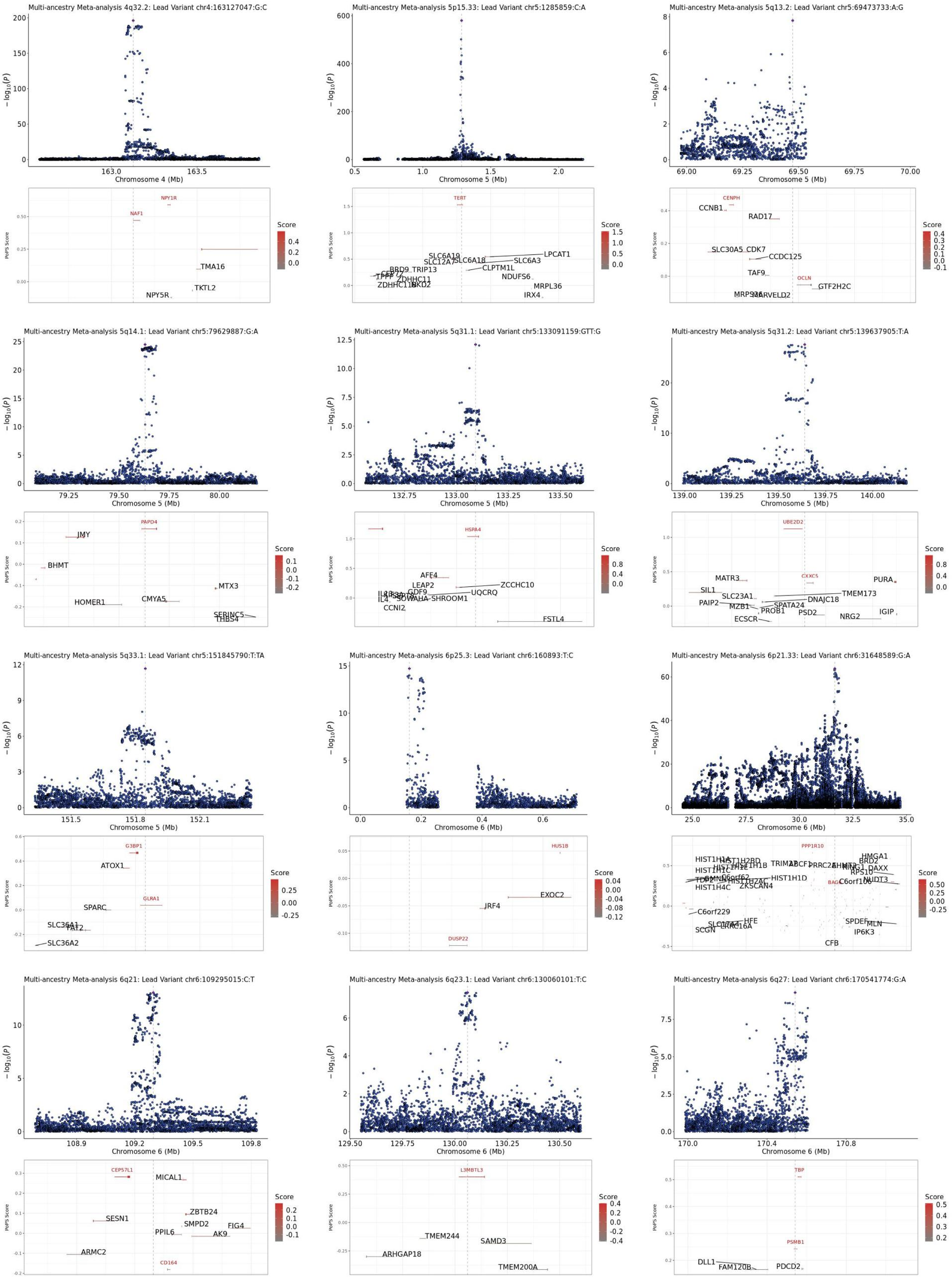

Multi-ancestry Meta-analysis 4q32.2: Lead Variant chr4:163127047:G:C
Chromosome 4 (Mb)
NPY1R
NAF1
TMA16
TKTL2
NPY5R
Multi-ancestry Meta-analysis 5p15.33: Lead Variant chr5:1285859:C:A
Chromosome 5 (Mb)
TERT
SLC6A19
SLC12A7
SLC6A18
LPCAT1
SLC6A3
CLPTM1L
NDUFS6
MRPL36
IRX4
BRD9
TRIP13
ZDHHC11
ZDHHC11B
Multi-ancestry Meta-analysis 5q13.2: Lead Variant chr5:69473733:A:G
Chromosome 5 (Mb)
CENPH
CCNB1
RAD17
SLC30A5
CDK7
CCDC125
TAF9
OCLN
GTF2H2C
MARVELD2
Multi-ancestry Meta-analysis 5q14.1: Lead Variant chr5:79629887:G:A
Chromosome 5 (Mb)
PAPD4
JMY
BHMT
MTX3
HOMER1
CMYA5
SERINC5
THBS4
Multi-ancestry Meta-analysis 5q31.1: Lead Variant chr5:133091159:GTT:G
Chromosome 5 (Mb)
HSPA4
AFF4
LEAP2
ZCCHC10
GDF9
UQCRQ
SHROOM1
CCNI2
FSTL4
Multi-ancestry Meta-analysis 5q31.2: Lead Variant chr5:139637905:T:A
Chromosome 5 (Mb)
UBE2D2
CXXC5
MATR3
PURA
SIL1
SLC23A1
TMEM173
DNAJC18
PAIP2
MZB1
SPATA24
PROB1
PSD2
IGIP
NRG2
ECSCR
Multi-ancestry Meta-analysis 5q33.1: Lead Variant chr5:151845790:T:TA
Chromosome 5 (Mb)
G3BP1
ATOX1
GLRA1
SPARC
SLC36A1
FAT2
SLC36A2
Multi-ancestry Meta-analysis 6p25.3: Lead Variant chr6:160893:T:C
Chromosome 6 (Mb)
HUS1B
EXOC2
IRF4
DUSP22
Multi-ancestry Meta-analysis 6p21.33: Lead Variant chr6:31648589:G:A
Chromosome 6 (Mb)
PPP1R10
HMGA1
BRD2
DAXX
RPS10
NUDT3
C6orf10
HIST1H1D
ZKSCAN4
C6orf229
SPDEF
MLN
IP6K3
HFE
SCGN
CFB
Multi-ancestry Meta-analysis 6q21: Lead Variant chr6:109295015:C:T
Chromosome 6 (Mb)
CEP57L1
MICAL1
SESN1
ZBTB24
SMPD2
FIG4
PPIL6
AK9
ARMC2
CD164
Multi-ancestry Meta-analysis 6q23.1: Lead Variant chr6:130060101:T:C
Chromosome 6 (Mb)
L3MBTL3
TMEM244
SAMD3
ARHGAP18
TMEM200A
Multi-ancestry Meta-analysis 6q27: Lead Variant chr6:170541774:G:A
Chromosome 6 (Mb)
TBP
PSMB1
DLL1
FAM120B
PDCD2

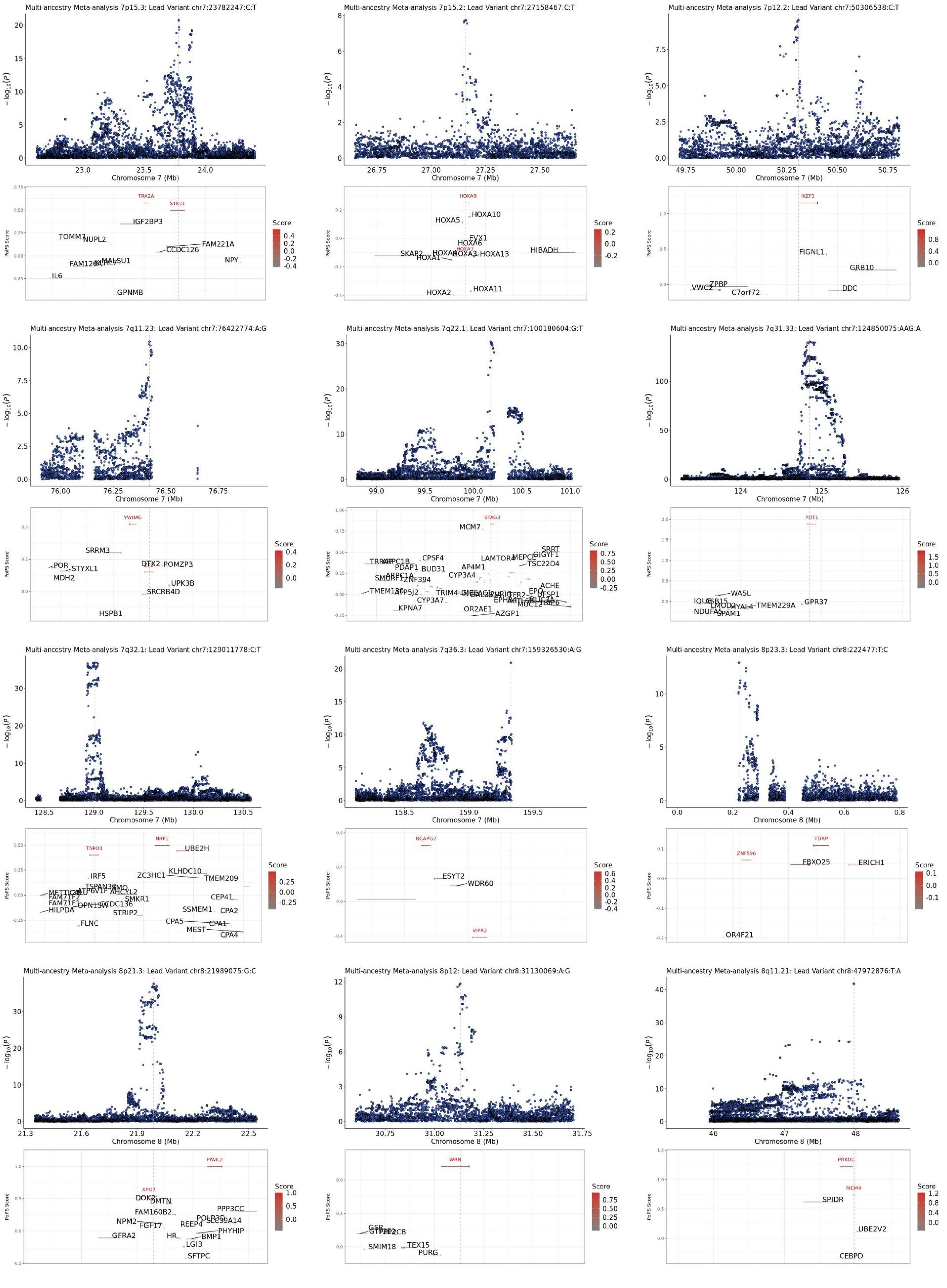

Multi-ancestry Meta-analysis 7p15.3: Lead Variant chr7:23782247:C:T
Multi-ancestry Meta-analysis 7p15.2: Lead Variant chr7:27158467:C:T
Multi-ancestry Meta-analysis 7p12.2: Lead Variant chr7:50306538:C:T
Multi-ancestry Meta-analysis 7q11.23: Lead Variant chr7:76422774:A:G
Multi-ancestry Meta-analysis 7q22.1: Lead Variant chr7:100180604:G:T
Multi-ancestry Meta-analysis 7q31.33: Lead Variant chr7:124850075:AAG:A
Multi-ancestry Meta-analysis 7q32.1: Lead Variant chr7:129011778:C:T
Multi-ancestry Meta-analysis 7q36.3: Lead Variant chr7:159326530:A:G
Multi-ancestry Meta-analysis 8p23.3: Lead Variant chr8:222477:T:C
Multi-ancestry Meta-analysis 8p21.3: Lead Variant chr8:21989075:G:C
Multi-ancestry Meta-analysis 8p12: Lead Variant chr8:31130069:A:G
Multi-ancestry Meta-analysis 8q11.21: Lead Variant chr8:47972876:T:A
$-\log_{10}(P)$
Chromosome 7 (Mb)
Chromosome 8 (Mb)
PoPS Score
Score
TRA2A
STK31
IGF2BP3
TOMM7
NUPL2
FAM221A
CCDC126
NPY
IL6
GPNMB
HOXA9
HOXA10
HOXA5
EVX1
HOXA6
HIBADH
SKAP2
HOXA1
HOXA13
HOXA2
HOXA11
IKZF1
FIGNL1
GRB10
ZPBP
VWC2
C7orf72
DDC
YWHAG
SRRM3
POR
STYXL1
MDH2
DTX2
POMZP3
UPK3B
SRCRB4D
HSPB1
STAG3
MCM7
CPSF4
LAMTOR4
MEPCE
TSC22D4
PDAP1
BUD31
AP4M1
CYP3A4
ZNF394
TMEM130
CYP3A7
KPNA7
OR2AE1
AZGP1
ACHE
POT1
WASL
TMEM229A
GPR37
SPAM1
TNPO3
NRF1
UBE2H
IRF5
ZC3HC1
KLHDC10
TMEM209
TSPAN33
SMKR1
CEP41
HILPDA
FLNC
STRIP2
SSMEM1
CPA2
CPA5
CPA1
MEST
CPA4
NCAPG2
ESYT2
WDR60
VIPR2
TDRP
ZNF596
FBXO25
ERICH1
OR4F21
PIWIL2
XPO7
DOK2
DMTN
FAM160B2
PPP3CC
NPM2
FGF17
REEP4
PHYHIP
GFRA2
BMP1
LGI3
SFTPC
WRN
GSR
SMIM18
TEX15
PURG
PRKDC
MCM4
SPIDR
UBE2V2
CEBPD

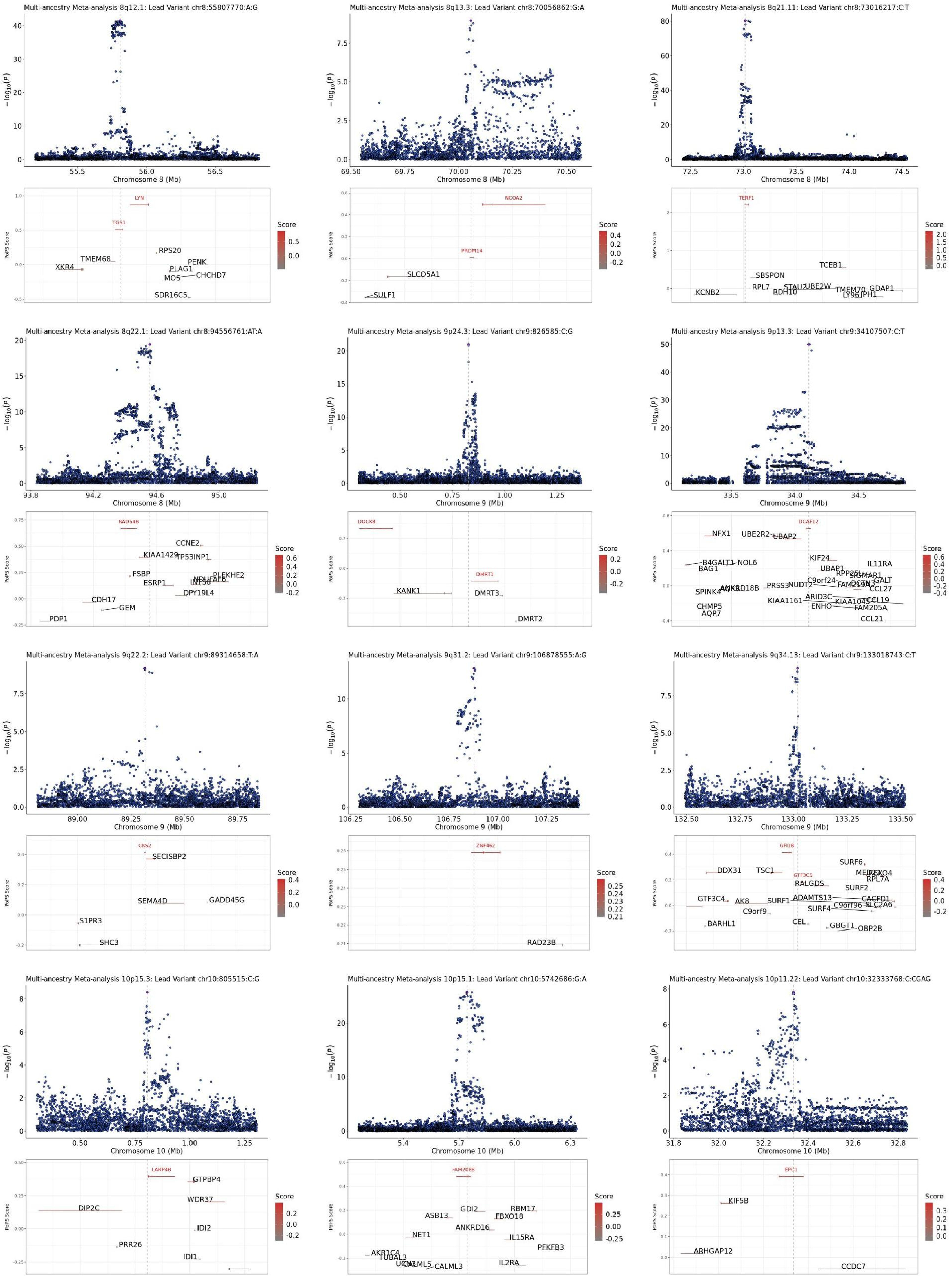

Multi-ancestry Meta-analysis 8q12.1: Lead Variant chr8:55807770:A:G
Chromosome 8 (Mb)
LYN
TGS1
RPS20
TMEM68
PENK
XKR4
PLAG1
MOS
CHCHD7
SDR16C5
Multi-ancestry Meta-analysis 8q13.3: Lead Variant chr8:70056862:G:A
Chromosome 8 (Mb)
NCOA2
PRDM14
SLCO5A1
SULF1
Multi-ancestry Meta-analysis 8q21.11: Lead Variant chr8:73016217:C:T
Chromosome 8 (Mb)
TERF1
TCEB1
SBSPON
RPL7
RDH10
UBE2W
KCNB2
TMEM70
GDAP1
Multi-ancestry Meta-analysis 8q22.1: Lead Variant chr8:94556761:AT:A
Chromosome 8 (Mb)
RAD54B
CCNE2
KIAA1429
TP53INP1
FSBP
ESRP1
PLEKHF2
DPY19L4
CDH17
GEM
PDP1
Multi-ancestry Meta-analysis 9p24.3: Lead Variant chr9:826585:C:G
Chromosome 9 (Mb)
DOCK8
DMRT1
KANK1
DMRT3
DMRT2
Multi-ancestry Meta-analysis 9p13.3: Lead Variant chr9:34107507:C:T
Chromosome 9 (Mb)
DCAF12
NFX1
UBE2R2
UBAP2
KIF24
B4GALT1
NOL6
IL11RA
BAG1
UBAP1
C9orf24
CCL27
SPINK4
PRSS3
NUDT2
ARID3C
CCL19
KIAA1161
ENHO
FAM205A
CHMP5
AQP7
CCL21
Multi-ancestry Meta-analysis 9q22.2: Lead Variant chr9:89314658:T:A
Chromosome 9 (Mb)
CKS2
SECISBP2
SEMA4D
GADD45G
S1PR3
SHC3
Multi-ancestry Meta-analysis 9q31.2: Lead Variant chr9:106878555:A:G
Chromosome 9 (Mb)
ZNF462
RAD23B
Multi-ancestry Meta-analysis 9q34.13: Lead Variant chr9:133018743:C:T
Chromosome 9 (Mb)
GFI1B
SURF6
DDX31
TSC1
GTF3C5
RPL7A
RALGDS
SURF2
GTF3C4
AK8
SURF1
ADAMTS13
C9orf96
SLC2A6
C9orf9
SURF4
CEL
BARHL1
GBGT1
OBP2B
Multi-ancestry Meta-analysis 10p15.3: Lead Variant chr10:805515:C:G
Chromosome 10 (Mb)
LARP4B
GTPBP4
WDR37
DIP2C
IDI2
PRR26
IDI1
Multi-ancestry Meta-analysis 10p15.1: Lead Variant chr10:5742686:G:A
Chromosome 10 (Mb)
FAM208B
GDI2
RBM17
ASB13
FBXO18
ANKRD16
NET1
IL15RA
PFKFB3
AKR1C4
CALML3
IL2RA
Multi-ancestry Meta-analysis 10p11.22: Lead Variant chr10:32333768:C:CGAG
Chromosome 10 (Mb)
EPC1
KIF5B
ARHGAP12
CCDC7
Score
PoPS Score

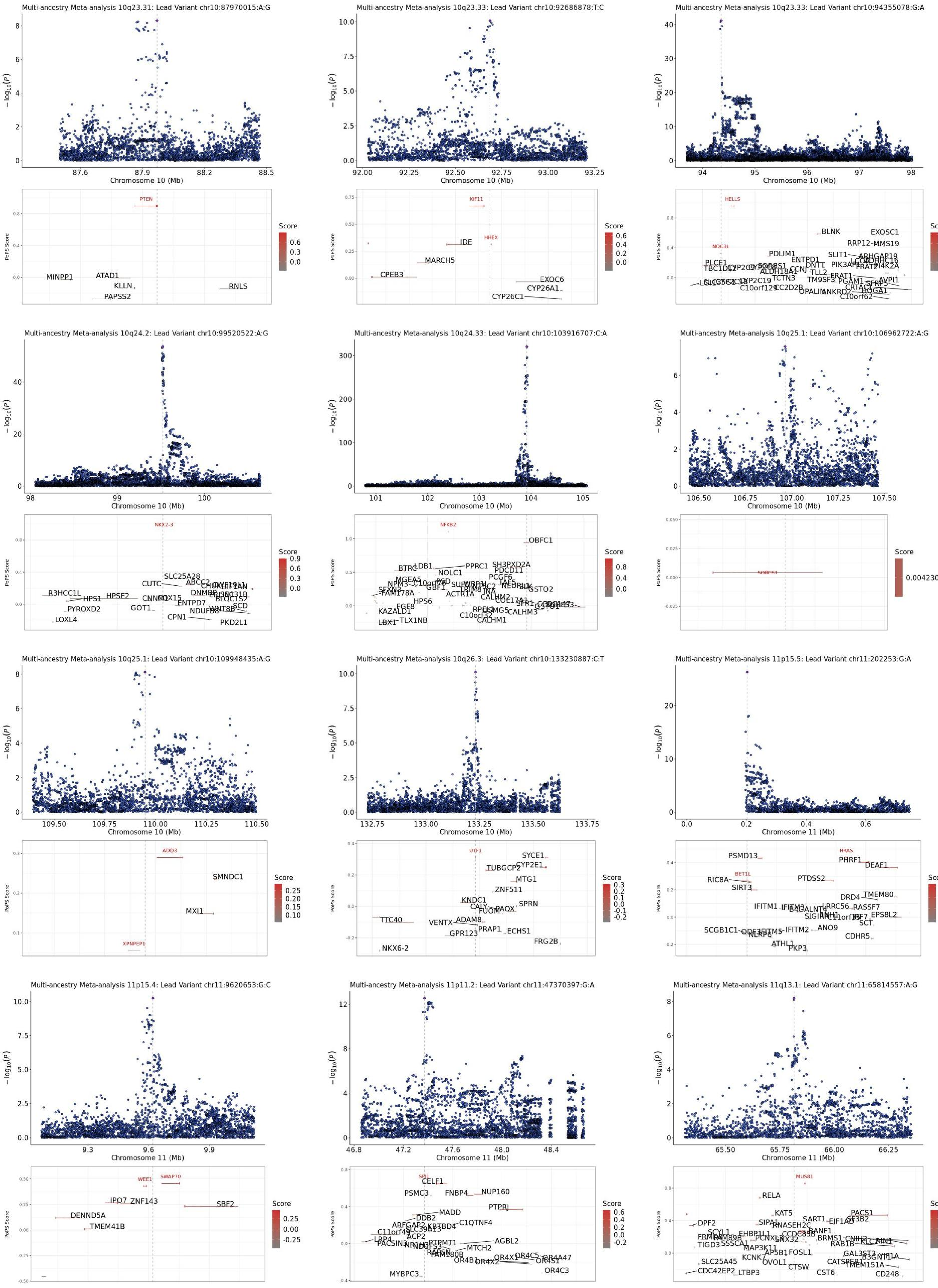

Multi-ancestry Meta-analysis 10q23.31: Lead Variant chr10:87970015:A:G
Multi-ancestry Meta-analysis 10q23.33: Lead Variant chr10:92686878:T:C
Multi-ancestry Meta-analysis 10q23.33: Lead Variant chr10:94355078:G:A
Multi-ancestry Meta-analysis 10q24.2: Lead Variant chr10:99520522:A:G
Multi-ancestry Meta-analysis 10q24.33: Lead Variant chr10:103916707:C:A
Multi-ancestry Meta-analysis 10q25.1: Lead Variant chr10:106962722:A:G
Multi-ancestry Meta-analysis 10q25.1: Lead Variant chr10:109948435:A:G
Multi-ancestry Meta-analysis 10q26.3: Lead Variant chr10:133230887:C:T
Multi-ancestry Meta-analysis 11p15.5: Lead Variant chr11:202253:G:A
Multi-ancestry Meta-analysis 11p15.4: Lead Variant chr11:9620653:G:C
Multi-ancestry Meta-analysis 11p11.2: Lead Variant chr11:47370397:G:A
Multi-ancestry Meta-analysis 11q13.1: Lead Variant chr11:65814557:A:G
$-\log_{10}(P)$
Chromosome 10 (Mb)
Chromosome 11 (Mb)
PoPS Score
Score
PTEN
MINPP1
ATAD1
KLLN
PAPSS2
RNLS
KIF11
HHEX
IDE
MARCH5
CPEB3
EXOC6
CYP26A1
CYP26C1
HELLS
NOC3L
BLNK
EXOSC1
NKX2-3
SLC25A28
R3HCC1L
HPS1
HPSE2
PYROXD2
GOT1
LOXL4
CPN1
PKD2L1
NFKB2
OBFC1
SORCS1
0.004230036
ADD3
SMNDC1
MXI1
XPNPEP1
UTF1
SYCE1
CYP2E1
TUBGCP2
MTG1
ZNF511
TTC40
NKX6-2
FRG2B
HRAS
PSMD13
PHRF1
DEAF1
BET1L
RIC8A
SIRT3
PTDSS2
WEE1
SWAP70
IPO7
ZNF143
SBF2
DENND5A
TMEM41B
SPI1
CELF1
PSMC3
FNBP4
NUP160
PTPRJ
MADD
MYBPC3
MUS81
RELA
KAT5
PACS1
CD248

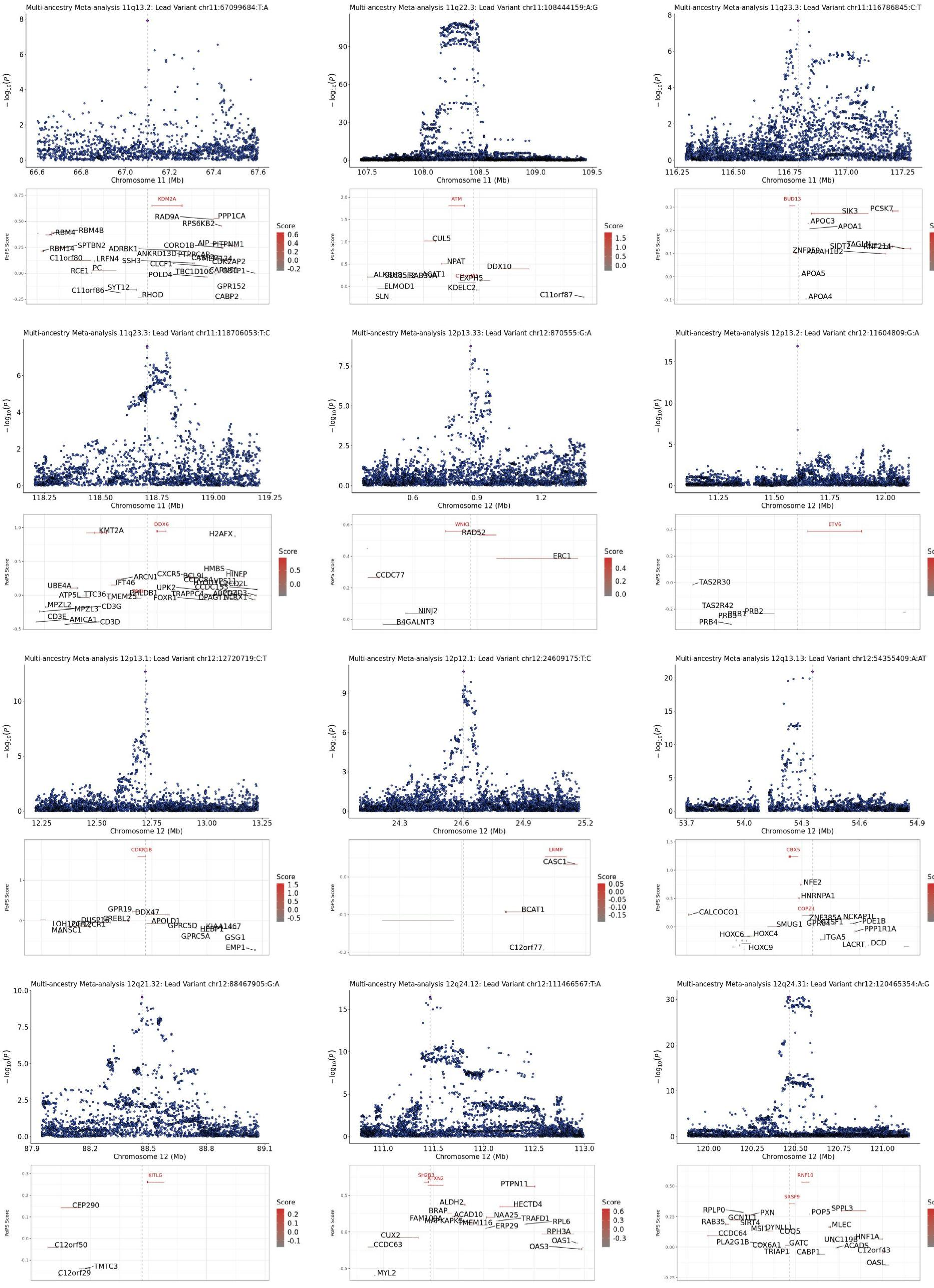

Multi-ancestry Meta-analysis 11q13.2: Lead Variant chr11:67099684:T:A
Multi-ancestry Meta-analysis 11q22.3: Lead Variant chr11:108444159:A:G
Multi-ancestry Meta-analysis 11q23.3: Lead Variant chr11:116786845:C:T
Multi-ancestry Meta-analysis 11q23.3: Lead Variant chr11:118706053:T:C
Multi-ancestry Meta-analysis 12p13.33: Lead Variant chr12:870555:G:A
Multi-ancestry Meta-analysis 12p13.2: Lead Variant chr12:11604809:G:A
Multi-ancestry Meta-analysis 12p13.1: Lead Variant chr12:12720719:C:T
Multi-ancestry Meta-analysis 12p12.1: Lead Variant chr12:24609175:T:C
Multi-ancestry Meta-analysis 12q13.13: Lead Variant chr12:54355409:A:AT
Multi-ancestry Meta-analysis 12q21.32: Lead Variant chr12:88467905:G:A
Multi-ancestry Meta-analysis 12q24.12: Lead Variant chr12:111466567:T:A
Multi-ancestry Meta-analysis 12q24.31: Lead Variant chr12:120465354:A:G
$-\log_{10}(P)$
Chromosome 11 (Mb)
Chromosome 12 (Mb)
PoPS Score
Score

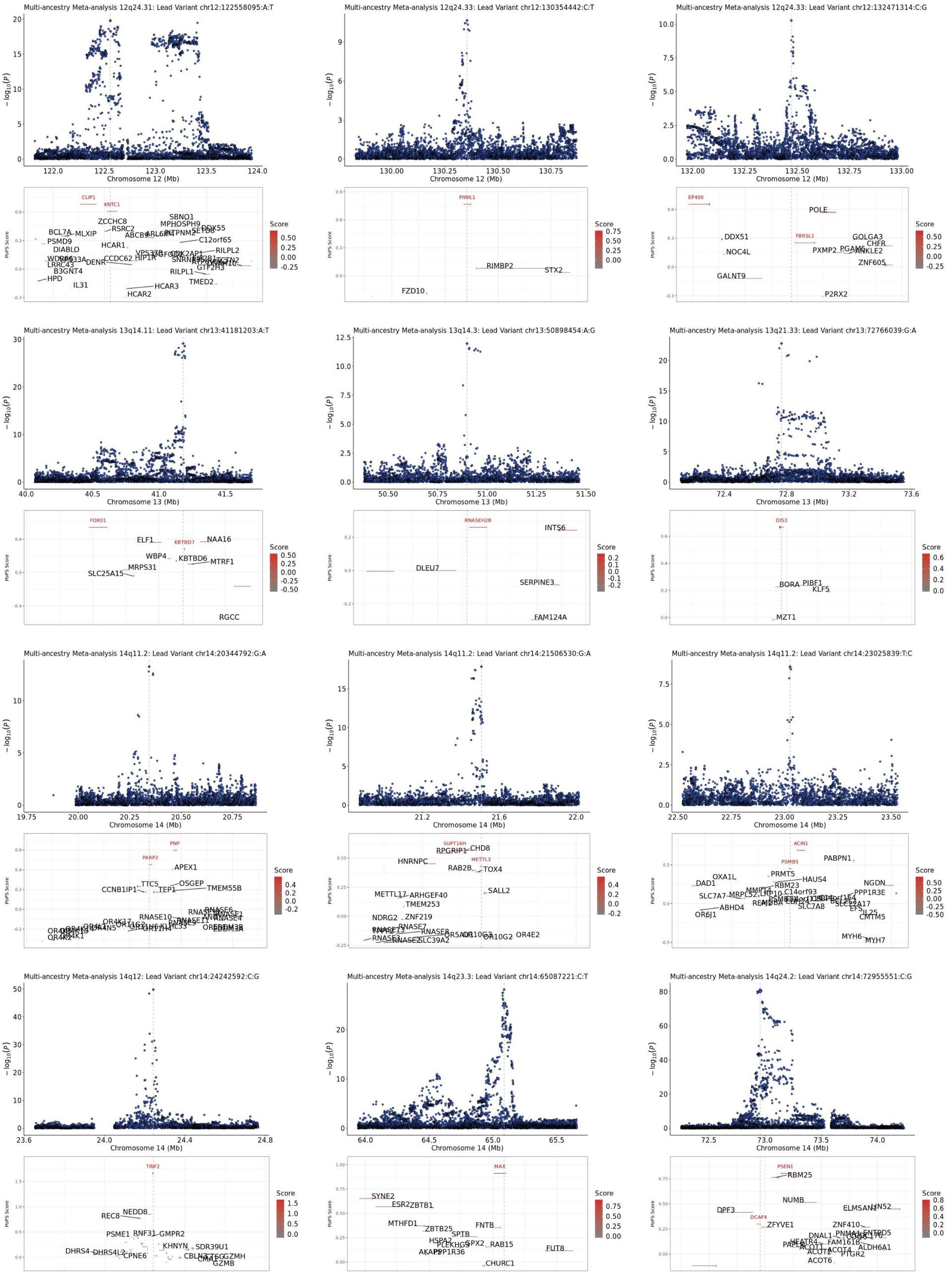

Multi-ancestry Meta-analysis 12q24.31: Lead Variant chr12:122558095:A:T
Chromosome 12 (Mb)
Multi-ancestry Meta-analysis 12q24.33: Lead Variant chr12:130354442:C:T
Chromosome 12 (Mb)
Multi-ancestry Meta-analysis 12q24.33: Lead Variant chr12:132471314:C:G
Chromosome 12 (Mb)
Multi-ancestry Meta-analysis 13q14.11: Lead Variant chr13:41181203:A:T
Chromosome 13 (Mb)
Multi-ancestry Meta-analysis 13q14.3: Lead Variant chr13:50898454:A:G
Chromosome 13 (Mb)
Multi-ancestry Meta-analysis 13q21.33: Lead Variant chr13:72766039:G:A
Chromosome 13 (Mb)
Multi-ancestry Meta-analysis 14q11.2: Lead Variant chr14:20344792:G:A
Chromosome 14 (Mb)
Multi-ancestry Meta-analysis 14q11.2: Lead Variant chr14:21506530:G:A
Chromosome 14 (Mb)
Multi-ancestry Meta-analysis 14q11.2: Lead Variant chr14:23025839:T:C
Chromosome 14 (Mb)
Multi-ancestry Meta-analysis 14q12: Lead Variant chr14:24242592:C:G
Chromosome 14 (Mb)
Multi-ancestry Meta-analysis 14q23.3: Lead Variant chr14:65087221:C:T
Chromosome 14 (Mb)
Multi-ancestry Meta-analysis 14q24.2: Lead Variant chr14:72955551:C:G
Chromosome 14 (Mb)
PoPS Score
Score

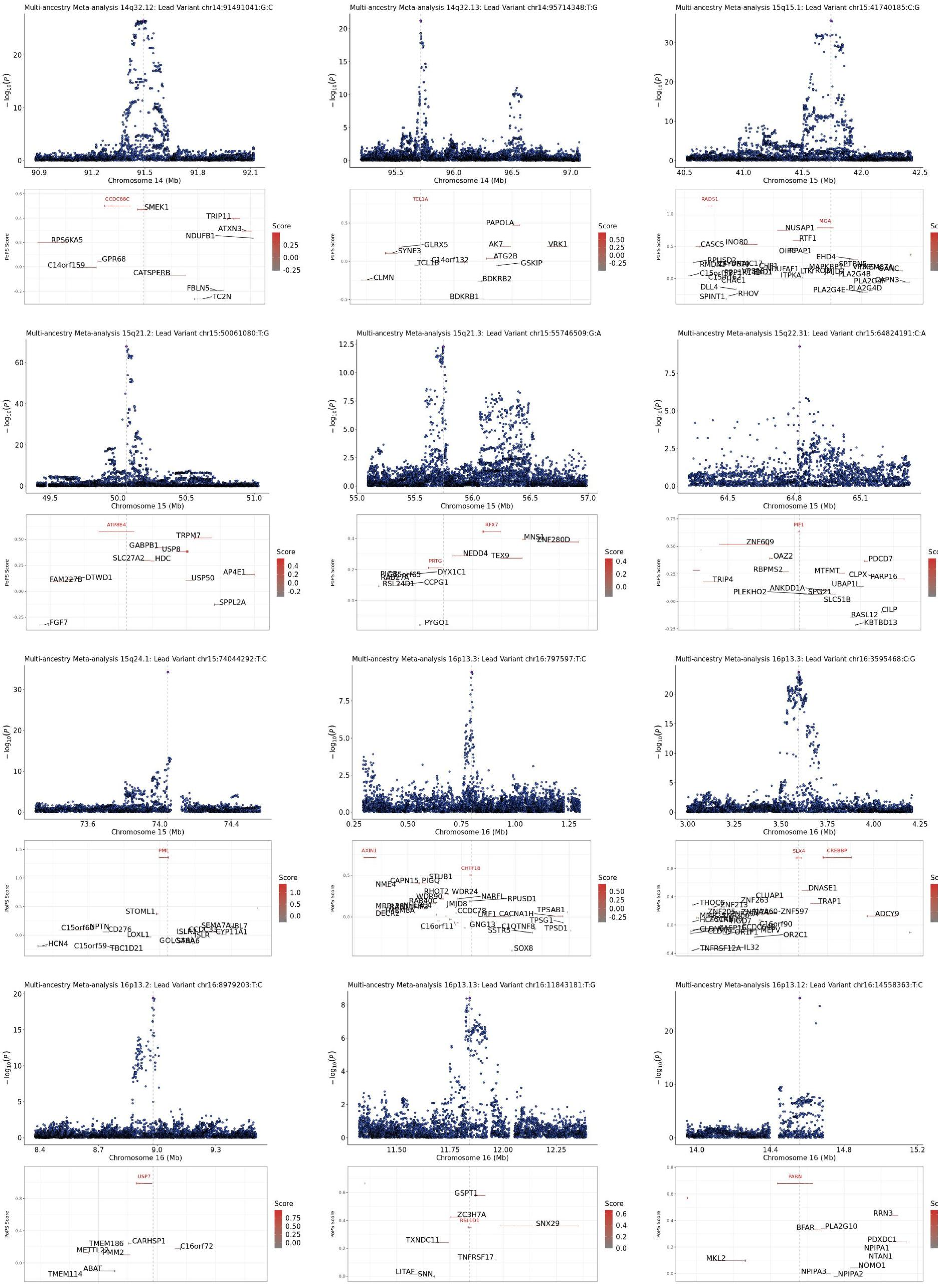

Multi-ancestry Meta-analysis 14q32.12: Lead Variant chr14:91491041:G:C
Chromosome 14 (Mb)
CCDC88C
SMEK1
TRIP11
ATXN3
NDUFB1
RPS6KA5
GPR68
C14orf159
CATSPERB
FBLN5
TC2N
Multi-ancestry Meta-analysis 14q32.13: Lead Variant chr14:95714348:T:G
Chromosome 14 (Mb)
TCL1A
PAPOLA
GLRX5
SYNE3
AK7
VRK1
ATG2B
C14orf132
TCL1B
GSKIP
CLMN
BDKRB2
BDKRB1
Multi-ancestry Meta-analysis 15q15.1: Lead Variant chr15:41740185:C:G
Chromosome 15 (Mb)
RAD51
MGA
NUSAP1
RTF1
CASC5
INO80
EHD4
DLL4
RHOV
SPINT1
PLA2G4E
PLA2G4D
PLA2G4B
Multi-ancestry Meta-analysis 15q21.2: Lead Variant chr15:50061080:T:G
Chromosome 15 (Mb)
ATP8B4
TRPM7
GABPB1
USP8
SLC27A2
HDC
FAM227B
DTWD1
AP4E1
USP50
SPPL2A
FGF7
Multi-ancestry Meta-analysis 15q21.3: Lead Variant chr15:55746509:G:A
Chromosome 15 (Mb)
RFX7
MNS1
ZNF280D
NEDD4
TEX9
PRTG
DYX1C1
RSL24D1
CCPG1
PYGO1
Multi-ancestry Meta-analysis 15q22.31: Lead Variant chr15:64824191:C:A
Chromosome 15 (Mb)
PIF1
ZNF609
OAZ2
PDCD7
RBPMS2
MTFMT
CLPX
PARP16
TRIP4
UBAP1L
ANKDD1A
PLEKHO2
SPG21
SLC51B
CILP
RASL12
KBTBD13
Multi-ancestry Meta-analysis 15q24.1: Lead Variant chr15:74044292:T:C
Chromosome 15 (Mb)
PML
STOML1
C15orf60
NPTN
CD276
LOXL1
HCN4
C15orf59
TBC1D21
GOLGA6A
SEMA7A
UBL7
CYP11A1
Multi-ancestry Meta-analysis 16p13.3: Lead Variant chr16:797597:T:C
Chromosome 16 (Mb)
AXIN1
CHTF18
STUB1
CAPN15
PIGQ
NME4
RHOT2
WDR24
NARFL
RPUSD1
JMJD8
CCDC78
TPSAB1
LMF1
CACNA1H
TPSG1
GNG13
C1QTNF8
TPSD1
SSTR5
C16orf11
DECR2
SOX8
Multi-ancestry Meta-analysis 16p13.3: Lead Variant chr16:3595468:C:G
Chromosome 16 (Mb)
SLX4
CREBBP
DNASE1
CLUAP1
TRAP1
THOC6
ZNF263
ZNF597
ADCY9
OR2C1
TNFRSF12A
IL32
Multi-ancestry Meta-analysis 16p13.2: Lead Variant chr16:8979203:T:C
Chromosome 16 (Mb)
USP7
TMEM186
CARHSP1
C16orf72
PMM2
ABAT
TMEM114
Multi-ancestry Meta-analysis 16p13.13: Lead Variant chr16:11843181:T:G
Chromosome 16 (Mb)
GSPT1
ZC3H7A
RSL1D1
SNX29
TXNDC11
TNFRSF17
LITAF
SNN
Multi-ancestry Meta-analysis 16p13.12: Lead Variant chr16:14558363:T:C
Chromosome 16 (Mb)
PARN
RRN3
BFAR
PLA2G10
PDXDC1
NPIPA1
NTAN1
MKL2
NOMO1
NPIPA3
NPIPA2
$-\log_{10}(P)$
PoPS Score
Score

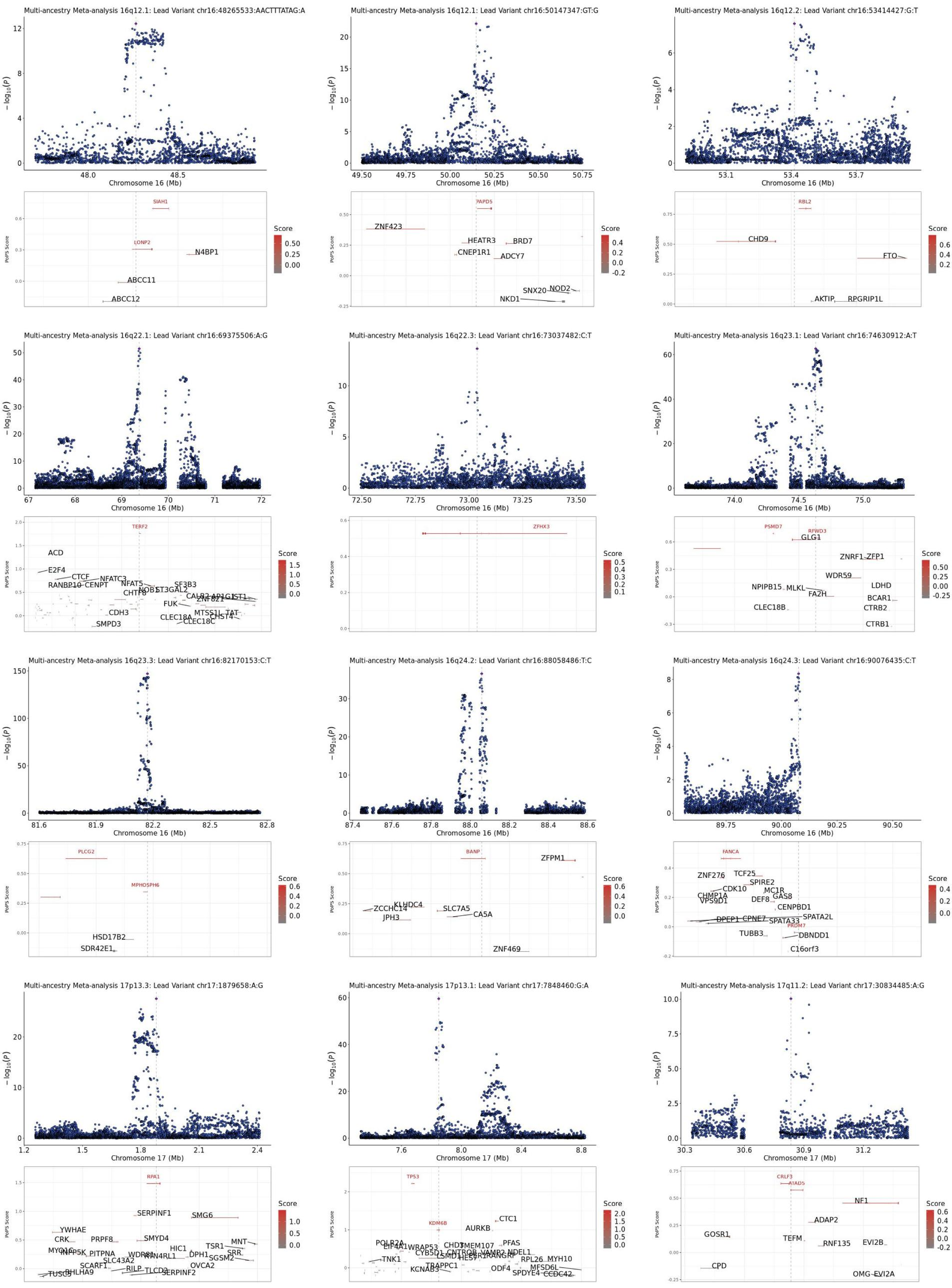

Multi-ancestry Meta-analysis 16q12.1: Lead Variant chr16:48265533:AACTTTATAG:A
Multi-ancestry Meta-analysis 16q12.1: Lead Variant chr16:50147347:GT:G
Multi-ancestry Meta-analysis 16q12.2: Lead Variant chr16:53414427:G:T
Multi-ancestry Meta-analysis 16q22.1: Lead Variant chr16:69375506:A:G
Multi-ancestry Meta-analysis 16q22.3: Lead Variant chr16:73037482:C:T
Multi-ancestry Meta-analysis 16q23.1: Lead Variant chr16:74630912:A:T
Multi-ancestry Meta-analysis 16q23.3: Lead Variant chr16:82170153:C:T
Multi-ancestry Meta-analysis 16q24.2: Lead Variant chr16:88058486:T:C
Multi-ancestry Meta-analysis 16q24.3: Lead Variant chr16:90076435:C:T
Multi-ancestry Meta-analysis 17p13.3: Lead Variant chr17:1879658:A:G
Multi-ancestry Meta-analysis 17p13.1: Lead Variant chr17:7848460:G:A
Multi-ancestry Meta-analysis 17q11.2: Lead Variant chr17:30834485:A:G
$-\log_{10}(P)$
Chromosome 16 (Mb)
Chromosome 17 (Mb)
PoPS Score
Score
SIAH1
LONP2
N4BP1
ABCC11
ABCC12
PAPD5
ZNF423
HEATR3
BRD7
CNEP1R1
ADCY7
SNX20
NOD2
NKD1
RBL2
CHD9
FTO
AKTIP
RPGRIP1L
TERF2
ACD
E2F4
CTCF
NFATC3
RANBP10
CENPT
NOB1
SF3B3
CHTF8
FUK
CDH3
SMPD3
CLEC18A
CLEC18C
MTSS1L
TAT
ZFHX3
PSMD7
RFWD3
GLG1
ZNRF1
ZFP1
WDR59
NPIPB15
MLKL
FA2H
LDHD
BCAR1
CTRB2
CLEC18B
CTRB1
PLCG2
MPHOSPH6
HSD17B2
SDR42E1
BANP
ZFPM1
KLHDC4
ZCCHC14
SLC7A5
JPH3
CA5A
ZNF469
FANCA
ZNF276
TCF25
SPIRE2
CDK10
MC1R
CHMP1A
VPS9D1
DEF8
GAS8
CENPBD1
SPATA2L
DPEP1
CPNE7
SPATA33
PRDM7
TUBB3
DBNDD1
C16orf3
RPA1
SERPINF1
SMG6
YWHAE
CRK
PRPF8
SMYD4
HIC1
TSR1
MNT
SGSM2
SRR
DPH1
OVCA2
SCARF1
RILP
TLCD2
SERPINF2
TUSC5
BHLHA9
TP53
KDM6B
CTC1
AURKB
PFAS
TNK1
NDEL1
RPL26
MYH10
ODF4
MFSD6L
SPDYE4
CCDC42
KCNAB3
TRAPPC1
CRLF3
ATAD5
NF1
ADAP2
GOSR1
TEFM
RNF135
EVI2B
CPD
OMG
EVI2A

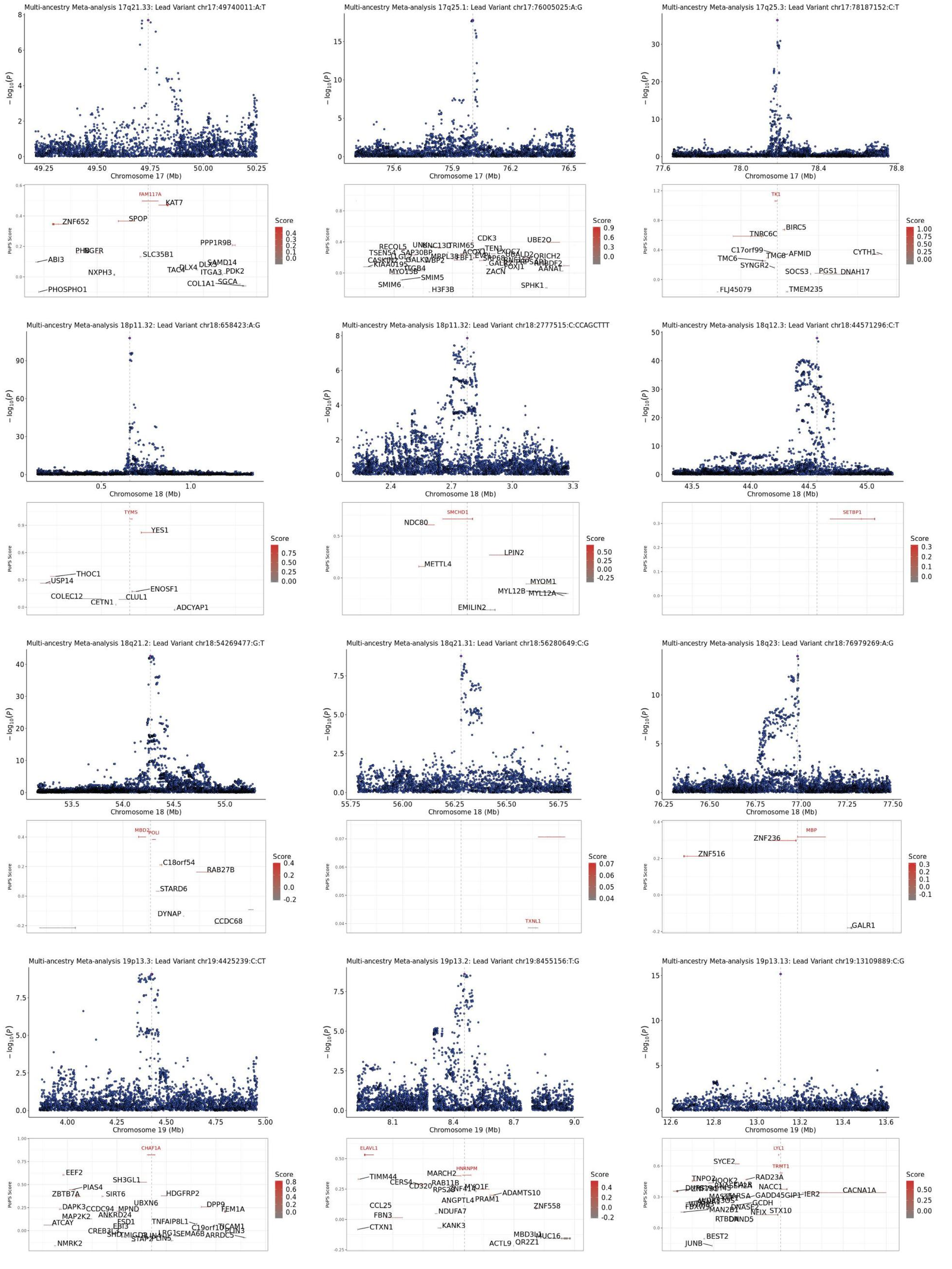

Multi-ancestry Meta-analysis 17q21.33: Lead Variant chr17:49740011:A:T
Chromosome 17 (Mb)
FAM117A
KAT7
ZNF652
SPOP
PPP1R9B
ABI3
SLC35B1
NXPH3
TAC4
DLX4
SAMD14
ITGA3
PDK2
COL1A1
SGCA
PHOSPHO1
Score
PoPS Score
Multi-ancestry Meta-analysis 17q25.1: Lead Variant chr17:76005025:A:G
Chromosome 17 (Mb)
CDK3
UBE2O
TSEN54
SAP30BP
MRPL38
CASKIN2
KIAA0195
MYO15B
ITGB4
SMIM5
SMIM6
H3F3B
ZACN
SPHK1
AANAT
Multi-ancestry Meta-analysis 17q25.3: Lead Variant chr17:78187152:C:T
Chromosome 17 (Mb)
TK1
BIRC5
TNRC6C
C17orf99
TMC8
AFMID
TMC6
CYTH1
SYNGR2
SOCS3
PGS1
DNAH17
FLJ45079
TMEM235
Multi-ancestry Meta-analysis 18p11.32: Lead Variant chr18:658423:A:G
Chromosome 18 (Mb)
TYMS
YES1
THOC1
USP14
ENOSF1
COLEC12
CLUL1
CETN1
ADCYAP1
Multi-ancestry Meta-analysis 18p11.32: Lead Variant chr18:2777515:C:CCAGCTTT
Chromosome 18 (Mb)
SMCHD1
NDC80
LPIN2
METTL4
MYOM1
MYL12B
MYL12A
EMILIN2
Multi-ancestry Meta-analysis 18q12.3: Lead Variant chr18:44571296:C:T
Chromosome 18 (Mb)
SETBP1
Multi-ancestry Meta-analysis 18q21.2: Lead Variant chr18:54269477:G:T
Chromosome 18 (Mb)
MBD2
POLI
C18orf54
RAB27B
STARD6
DYNAP
CCDC68
Multi-ancestry Meta-analysis 18q21.31: Lead Variant chr18:56280649:C:G
Chromosome 18 (Mb)
TXNL1
Multi-ancestry Meta-analysis 18q23: Lead Variant chr18:76979269:A:G
Chromosome 18 (Mb)
MBP
ZNF236
ZNF516
GALR1
Multi-ancestry Meta-analysis 19p13.3: Lead Variant chr19:4425239:C:CT
Chromosome 19 (Mb)
CHAF1A
EEF2
SH3GL1
PIAS4
ZBTB7A
SIRT6
HDGFRP2
UBXN6
DAPK3
CCDC94
MPND
DPP9
FEM1A
MAP2K2
ANKRD24
ATCAY
EBI3
TNFAIP8L1
C19orf10
CREB3L3
LRG1
SEMA6B
ARRDC5
NMRK2
Multi-ancestry Meta-analysis 19p13.2: Lead Variant chr19:8455156:T:G
Chromosome 19 (Mb)
ELAVL1
HNRNPM
TIMM44
MARCH2
CERS4
RAB11B
MYO1F
ADAMTS10
PRAM1
ANGPTL4
ZNF558
CCL25
NDUFA7
FBN3
CTXN1
KANK3
MBD3L1
MUC16
ACTL9
OR2Z1
Multi-ancestry Meta-analysis 19p13.13: Lead Variant chr19:13109889:C:G
Chromosome 19 (Mb)
LYL1
TRMT1
SYCE2
RAD23A
TNPO2
NACC1
CACNA1A
GADD45GIP1
IER2
MAN2B1
NFIX
STX10
RTBDN
BEST2
JUNB

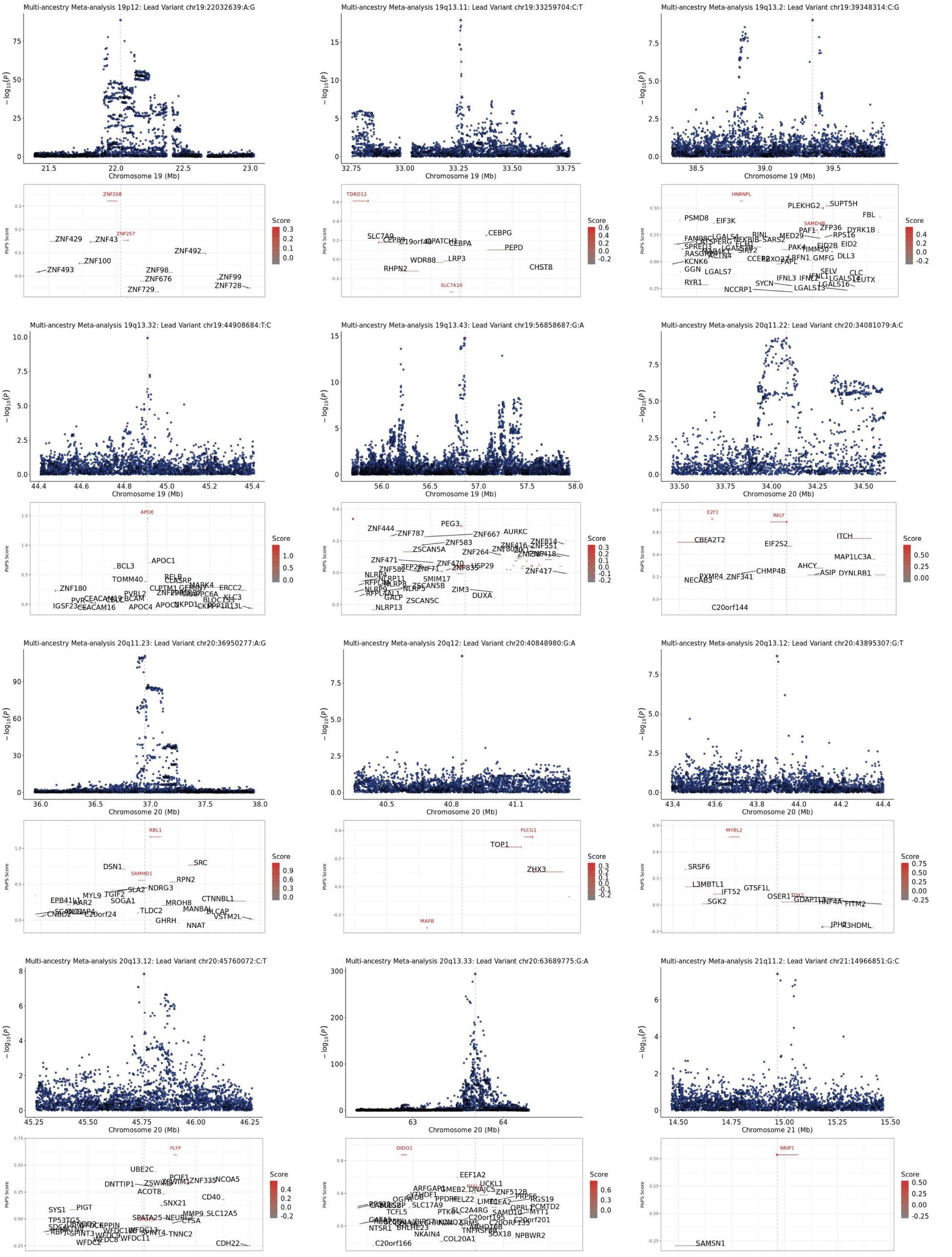

Multi-ancestry Meta-analysis 19p12: Lead Variant chr19:22032639:A:G
Multi-ancestry Meta-analysis 19q13.11: Lead Variant chr19:33259704:C:T
Multi-ancestry Meta-analysis 19q13.2: Lead Variant chr19:39348314:C:G
Multi-ancestry Meta-analysis 19q13.32: Lead Variant chr19:44908684:T:C
Multi-ancestry Meta-analysis 19q13.43: Lead Variant chr19:56858687:G:A
Multi-ancestry Meta-analysis 20q11.22: Lead Variant chr20:34081079:A:C
Multi-ancestry Meta-analysis 20q11.23: Lead Variant chr20:36950277:A:G
Multi-ancestry Meta-analysis 20q12: Lead Variant chr20:40848980:G:A
Multi-ancestry Meta-analysis 20q13.12: Lead Variant chr20:43895307:G:T
Multi-ancestry Meta-analysis 20q13.12: Lead Variant chr20:45760072:C:T
Multi-ancestry Meta-analysis 20q13.33: Lead Variant chr20:63689775:G:A
Multi-ancestry Meta-analysis 21q11.2: Lead Variant chr21:14966851:G:C
$-\log_{10}(P)$
Chromosome 19 (Mb)
Chromosome 20 (Mb)
Chromosome 21 (Mb)
PoPS Score
Score
ZNF208
ZNF257
ZNF429
ZNF43
ZNF100
ZNF493
ZNF492
ZNF98
ZNF676
ZNF729
ZNF99
ZNF728
TDRD12
SLC7A9
CEBPG
CEBPA
PEPD
LRP3
WDR88
RHPN2
CHST8
SLC7A10
HNRNPL
PLEKHG2
SUPT5H
PSMD8
EIF3K
FBL
SAMD4B
ZFP36
DYRK1B
RPS16
EID2
DLL3
KCNK6
GGN
LGALS7
RYR1
SELV
CLC
LEUTX
IFNL3
LGALS16
LGALS13
SYCN
NCCRP1
APOE
APOC1
BCL3
TOMM40
ZNF180
IGSF23
CEACAM16
APOC4
ERCC2
PEG3
ZNF444
ZNF787
ZNF667
AURKC
ZNF583
ZSCAN5A
ZNF264
ZNF471
ZNF417
USP29
SMIM17
ZIM3
DUXA
ZSCAN5C
NLRP13
E2F1
RALY
CBFA2T2
EIF2S2
ITCH
MAP1LC3A
AHCY
ASIP
DYNLRB1
CHMP4B
ZNF341
NECAB3
C20orf144
RBL1
DSN1
SAMHD1
SRC
RPN2
NDRG3
SLA2
TGIF2
MYL9
SOGA1
TLDC2
MROH8
CTNNBL1
MANBAL
BLCAP
VSTM2L
GHRH
NNAT
C20orf24
PLCG1
TOP1
ZHX3
MAFB
MYBL2
SRSF6
L3MBTL1
IFT52
GTSF1L
SGK2
OSER1
TOX2
GDAP1L1
HNF4A
FITM2
JPH2
R3HDML
PLTP
UBE2C
PCIF1
ZNF335
NCOA5
DNTTIP1
ACOT8
CD40
SNX21
SYS1
PIGT
SPATA25
MMP9
SLC12A5
CTSA
TNNC2
WFDC2
WFDC11
CDH22
DIDO1
EEF1A2
UCKL1
ZNF512B
PRPF6
RGS19
PCMTD2
MYT1
SLC17A9
TCFL5
PTK6
SAMD10
NTSR1
BHLHE23
NKAIN4
COL20A1
SOX18
NPBWR2
C20orf166
NRIP1
SAMSN1

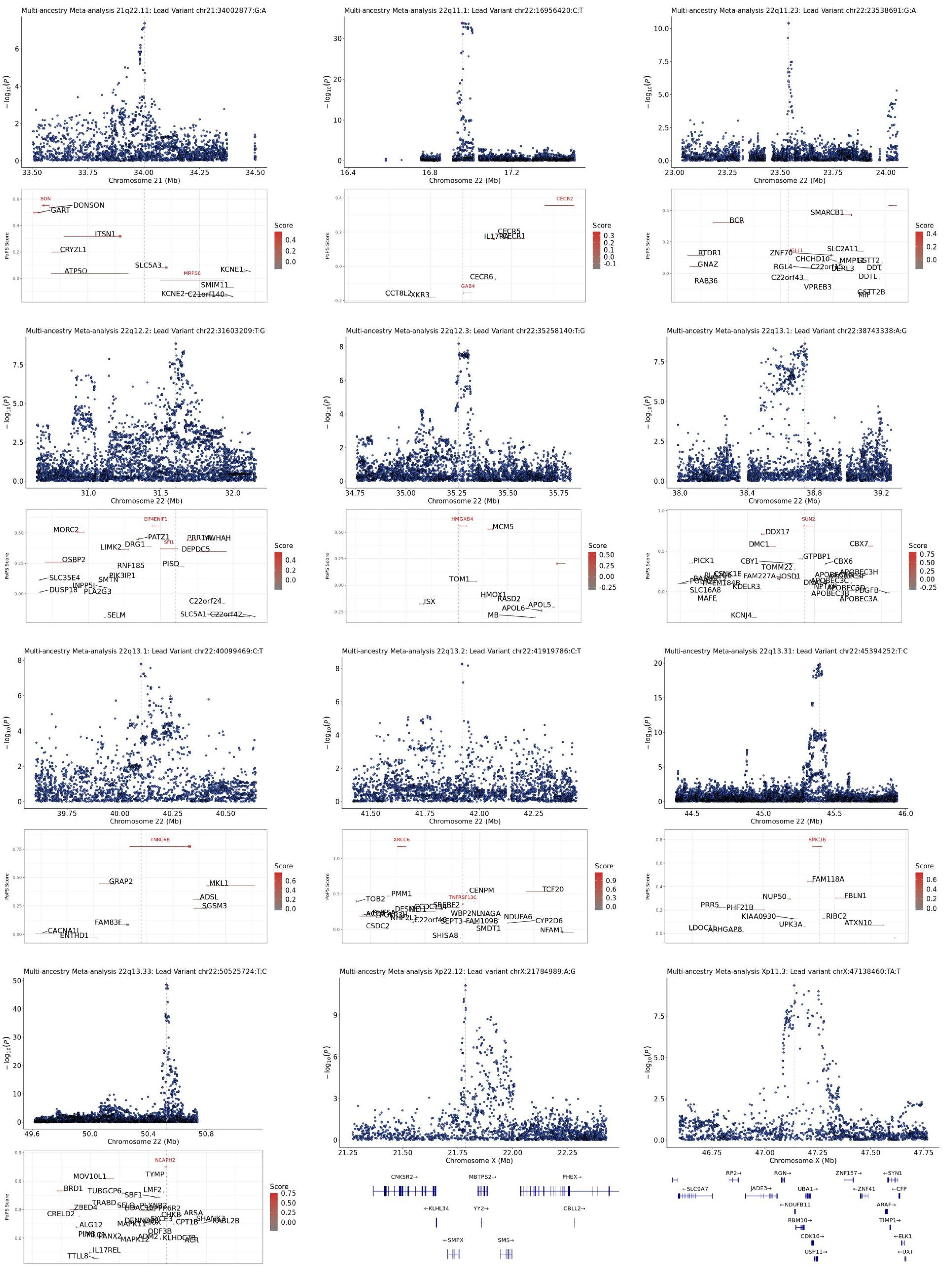

Multi-ancestry Meta-analysis 21q22.11: Lead Variant chr21:34002877:G:A
Chromosome 21 (Mb)
Multi-ancestry Meta-analysis 22q11.1: Lead Variant chr22:16956420:C:T
Chromosome 22 (Mb)
Multi-ancestry Meta-analysis 22q11.23: Lead Variant chr22:23538691:G:A
Chromosome 22 (Mb)
Multi-ancestry Meta-analysis 22q12.2: Lead Variant chr22:31603209:T:G
Chromosome 22 (Mb)
Multi-ancestry Meta-analysis 22q12.3: Lead Variant chr22:35258140:T:G
Chromosome 22 (Mb)
Multi-ancestry Meta-analysis 22q13.1: Lead Variant chr22:38743338:A:G
Chromosome 22 (Mb)
Multi-ancestry Meta-analysis 22q13.1: Lead Variant chr22:40099469:C:T
Chromosome 22 (Mb)
Multi-ancestry Meta-analysis 22q13.2: Lead Variant chr22:41919786:C:T
Chromosome 22 (Mb)
Multi-ancestry Meta-analysis 22q13.31: Lead Variant chr22:45394252:T:C
Chromosome 22 (Mb)
Multi-ancestry Meta-analysis 22q13.33: Lead Variant chr22:50525724:T:C
Chromosome 22 (Mb)
Multi-ancestry Meta-analysis Xp22.12: Lead variant chrX:21784989:A:G
Chromosome X (Mb)
Multi-ancestry Meta-analysis Xp11.3: Lead variant chrX:47138460:TA:T
Chromosome X (Mb)
−log10(P)
PoPS Score
Score

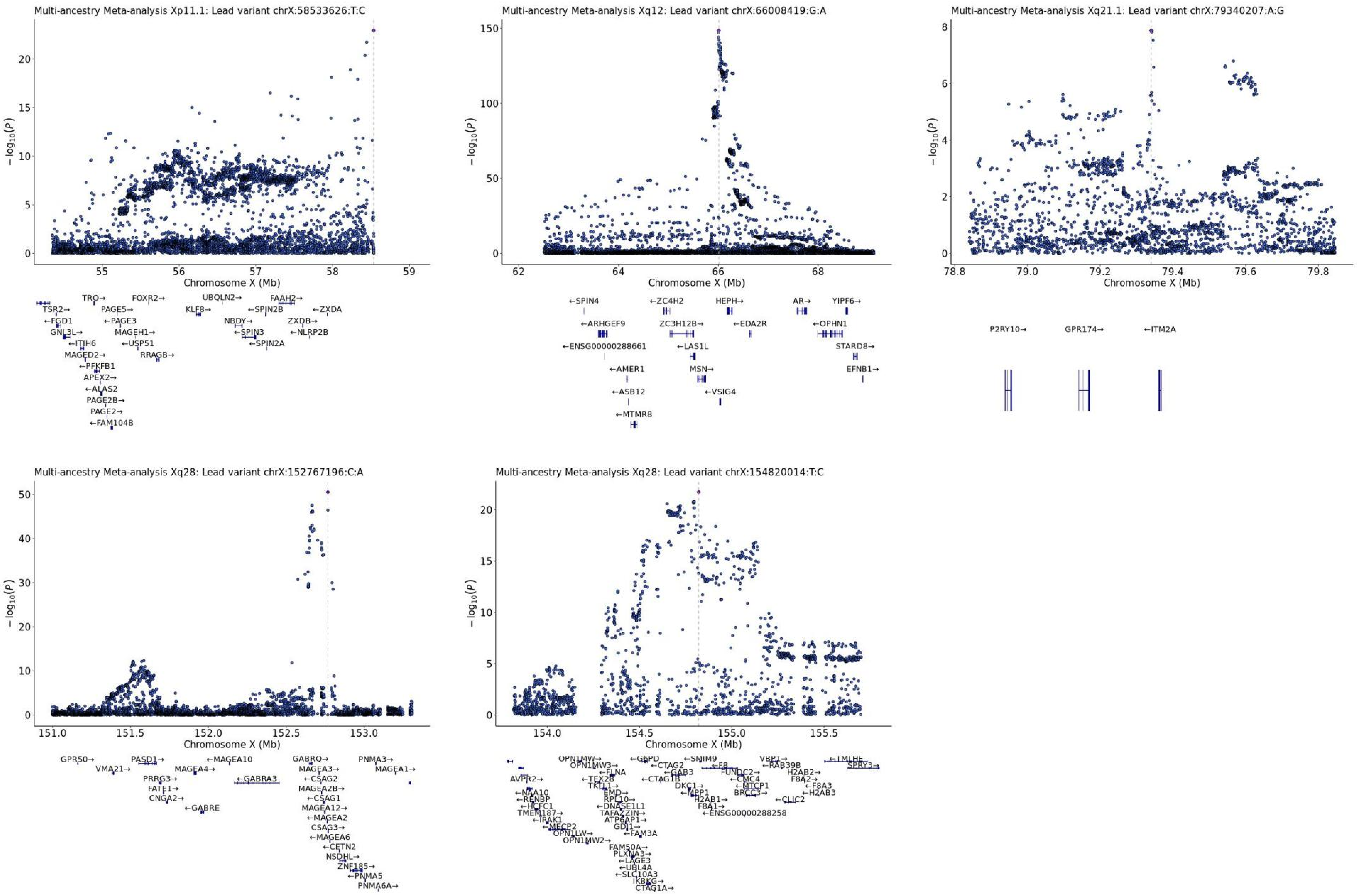
Multi-ancestry Meta-analysis Xp11.1: Lead variant chrX:58533626:T:C
Multi-ancestry Meta-analysis Xq12: Lead variant chrX:66008419:G:A
Multi-ancestry Meta-analysis Xq21.1: Lead variant chrX:79340207:A:G
Multi-ancestry Meta-analysis Xq28: Lead variant chrX:152767196:C:A
Multi-ancestry Meta-analysis Xq28: Lead variant chrX:154820014:T:C
$-\log_{10}(P)$
Chromosome X (Mb)
TRO→
FOXR2→
UBQLN2→
FAAH2→
TSR2→
PAGE5→
KLF8→
←SPIN2B
←ZXDA
←FGD1
←PAGE3
NBDY→
ZXDB→
GNL3L→
MAGEH1→
←SPIN3
←NLRP2B
←ITIH6
←USP51
←SPIN2A
MAGED2→
RRAGB→
←PFKFB1
APEX2→
←ALAS2
PAGE2B→
PAGE2→
←FAM104B
←SPIN4
←ZC4H2
HEPH→
AR→
YIPF6→
←ARHGEF9
ZC3H12B→
←EDA2R
←OPHN1
←ENSG00000288661
←LAS1L
STARD8→
←AMER1
MSN→
EFNB1→
←ASB12
←VSIG4
←MTMR8
P2RY10→
GPR174→
←ITM2A
GPR50→
PASD1→
←MAGEA10
GABRQ→
PNMA3→
VMA21→
MAGEA4→
MAGEA3→
MAGEA1→
PRRG3→
←GABRA3
←CSAG2
FATE1→
MAGEA2B→
CNGA2→
←CSAG1
←GABRE
MAGEA12→
←MAGEA2
CSAG3→
←MAGEA6
←CETN2
NSDHL→
ZNF185→
←PNMA5
PNMA6A→
OPN1MW→
←G6PD
←SMIM9
VBP1→
←TMLHE
OPN1MW3→
←CTAG2
←F8
←RAB39B
SPRY3→
←FLNA
←GAB3
FUNDC2→
H2AB2→
AVPR2→
←TEX28
←CTAG1B
←CMC4
F8A2→
TKTL1→
DKC1→
←MTCP1
←F8A3
←NAA10
EMD→
←MPP1
BRCC3→
←H2AB3
←RENBP
RPL10→
H2AB1→
←CLIC2
←HCFC1
←DNASE1L1
F8A1→
TMEM187→
TAFAZZIN→
←ENSG00000288258
←IRAK1
ATP6AP1→
←MECP2
GDI1→
OPN1LW→
←FAM3A
OPN1MW2→
FAM50A→
PLXNA3→
←LAGE3
←UBL4A
←SLC10A3
IKBKG→
CTAG1A→
